# Community health workers and adolescent health and well-being across sub-Saharan Africa: A systematic review and meta-analysis

**DOI:** 10.1101/2025.09.26.25336574

**Authors:** Yangxu Lin, Oluwatoni Adesina, Broline Sagini Asuma, Jenny Chen-Charles, Satvika Chalasani, Lucie Cluver, Rosalie Aduayi Diop, William E. Rudgard

**Affiliations:** Department of Social Policy and Intervention, University of Oxford, Oxford, United Kingdom; Programme Group, UNICEF, New York, NY 10003, USA; Department of Psychiatry and Mental Health, University of Cape Town, Cape Town, South Africa; Principal Researcher, Lecturer at the Institute of Population, Development and Reproductive Health, Cheikh Anta Diop University, Dakar IPDSR/UCAD; Centre for Social Science Research, University of Cape Town, Cape Town, South Africa

## Abstract

**Introduction:** Adolescence represents a pivotal stage of development. Community health workers (CHWs) may offer a valuable approach to addressing adolescent health challenges in sub-Saharan Africa (SSA). This review systematically synthesised evidence on the effectiveness of CHW-led interventions to improve adolescent health and well-being in SSA.

**Methods:** We searched 14 databases and 17 grey literature sources for studies published between January 2014 and October 2024. The protocol was registered in PROSPERO (CRD42024618546). Interventions were categorised by setting (community or school) and provider (lay or paraprofessional; peer- or adult-led). Vote-counting and meta-analyses were conducted, alongside risk of bias using Joanna Briggs Institute tools and evidence quality assessments using the GRADE approach.

**Results:** Thirty-five studies were included spanning across 12 countries and 12 outcomes. All outcomes were summarised narratively. Meta-analyses showed very low-certainty strong evidence for increased modern contraceptive use (RR: 1.50, 95% confidence interval [CI] 1.13-1.99), low-certainty moderate evidence for reduced depression (SMD: −0.31, 95% CI −0.61-0.00), and low-certainty weak evidence for improved human immunodeficiency virus viral suppression (RR: 1.16, 95% CI 0.99-1.36). No clear effects were observed for sexual health knowledge, unmet need for contraception, or adolescent pregnancy, all with very-low certainty.

**Discussion:** CHW-led interventions show promise for improving adolescent contraceptive use and mental health in SSA. However, substantial heterogeneity warrants cautious interpretation, and more rigorous evidence is needed across all health domains.

**Implications and Contribution:** Drawing on 35 studies, this systematic review consolidates evidence on community health workers’ role in supporting adolescent health across Africa. It shows potential to improve modern contraceptive use, mental health, and HIV viral load suppression, while exposing evidence gaps in other domains and setting priorities for future investment and research.

## Introduction

Adolescence, particularly early adolescence, is a formative period of growth and development [1]. It offers a critical window to shape lifelong health behaviour and values, offering an ideal opportunity to promote healthy habits and informed decision-making [2,3]. The stakes are particularly high in Africa, which is home to nearly 258 million youth in 2020, making it the world’s youngest region [4].

Despite progress in many areas, adolescents in sub-Saharan Africa (SSA) continue to face substantial and interconnected health and well-being challenges [5]. Undernutrition remains widespread, while overweight and obesity are rising [6]. Antiretroviral treatment (ART) coverage remains below target, particularly in Eastern and Southern Africa [7]. Mental health conditions, such as post-traumatic stress (PTS) and depression, are increasingly recognised but remain inadequately addressed [8,9]. Educational outcomes also lag, with low completion of lower secondary school and persistent literacy gaps [10]. Adolescent girls face additional risks, including sexual coercion, gender-based violence, child marriage, unmet need for modern contraception, and early pregnancy [11,12].

These challenges are compounded by persistent barriers to accessing high-quality, youth-friendly services, including financial constraints, long distances to facilities, low health literacy, judgemental provider attitudes, lack of privacy, inconvenient service hours, fragmented care, and restricted social norms [13–15]. Community health workers (CHWs) may offer a promising approach to mitigating some of these challenges. CHWs comprise a broad and heterogeneous group of providers. They are generally defined as lay or paraprofessional individuals who deliver health-related interventions without formal clinical qualifications, including roles such as peer educators, lay counsellors, outreach workers, and trained community volunteers, and excluding licensed healthcare professionals [16]. Widely deployed across Africa, CHWs deliver culturally responsive health promotion and education, extend facility-based services to households, and facilitate linkages to formal healthcare systems [17,18].

While CHWs have demonstrated effectiveness in maternal and child health, immunisation, and communicable diseases like human immunodeficiency virus (HIV), tuberculosis (TB), and malaria, their role specifically in adolescent health remains underexplored. A 2013 review on CHWs and adolescent health services in Africa identified only one relevant study, with unclear attribution of CHW effects within a multi-component intervention, highlighting a significant evidence gap in the literature [19]. Since then, CHW-based approaches have expanded, yet the evidence on their effectiveness remains fragmented and has not been comprehensively synthesised in recent years.

This review revisited this topic and extended the evidence base by synthesising evidence on CHW-led interventions targeting adolescent health and well-being outcomes published between 2014 and 2024. Given the diversity of outcomes reported in the literature and the cross-cutting nature of CHW interventions, we assessed whether interventions improved outcomes across multiple adolescent health and well-being domains [20] rather than focusing on a single outcome type. Specifically, the review aimed to (i) map and synthesis the characteristics of CHW-delivered interventions targeting adolescent health and well-being in SSA, (ii) evaluate intervention effectiveness, and (iii) examine gender differences in outcomes.

## Methods

We conducted a Preferred Reporting Items for Systematic reviews and Meta-Analyses (PRISMA)-compliant review of the role of CHWs in improving adolescent health and wellbeing across SSA [21,22]. The review protocol was registered in PROSPERO (CRD42024618546). This analysis focuses exclusively on quantitative studies, as integrating qualitative evidence was beyond the scope of reporting in a single journal article.

### Search and Information Sources

The following electronic bibliographic databases were searched: PubMed, Embase, Global Health, Medline, PsychINFO, Scopus, CINAHL, ERIC, Web of Science, Proquest Social Science, EconLit, Premium, Cochrane Library, and Africa-Wide Information.

We supplemented our bibliographic database searches with grey literature searches on Google Scholar and 16 websites of relevant implementing agencies and international organisations (see Appendix A.1). Searches on Google Scholar and other websites were limited to the first 100 results.

The search string was composed of three elements: 1) adolescents, 2) CHWs (-delivered interventions), and 3) Africa^1^. We did not include a specific outcome element in our search string due to the broad nature of our focus on adolescent health and well-being. We consulted colleagues at UNICEF and a Bodleian librarian at the University of Oxford to refine our information sources and search terms. Search terms used for all bibliographic and grey literature databases were provided in Appendix A.2 and A.4 respectively.

We also conducted backward citation searches of all included studies, and two highly relevant systematic reviews published in 2023 [23,24]. The reviews each focused on a single outcome domain, and examined peer support or peer-led interventions, which may partially overlap with peer CHW-delivered interventions. This step aimed to minimise the risk of omitting eligible studies.

### Eligibility criteria

Quantitative studies and quantitative sections from mixed-methods studies published from January 2014 to October 2024 were eligible for inclusion. This predefined timeframe was selected to synthesise evidence from the most recent decade of published literature available at the commencement of the review project, while building on the existing evidence base established by the previous systematic review [19].

Eligible studies focused on interventions delivered by CHWs and their effect on adolescent health and well-being outcomes in Eastern, Southern, Western and Central Africa [25]. In this review, CHWs were defined according to the World Health Organisation (WHO) definition as health workers based in communities, conducting outreach beyond primary healthcare facilities, or working at peripheral health posts not staffed by doctors or nurses^2^.

They typically receive less than two years of formal training but at least some training if only for a few hours and are either paid or volunteer. We further categorised CHWs as adolescent peers if they were aged <24, and adults if they were aged 24 years and older, and as either lay workers or paraprofessionals largely based on Olaniran et al. (2021) [16] (for detailed definitions we used, see Appendix B).

Our target population for this review was adolescents aged 10-19. We also included studies with broader age ranges if they met one of the following criteria: (1) they reported age-disaggregated results for a subgroup within the 10–19 age range, or (2) the median (or mean if the median was unavailable) age of participants fell between 10 and 19 years, and the overall age range lay between five and twenty-four years old. Where populations were referred to as senior primary, secondary or first-year high school students, we assumed that their age range fitted the criteria above. This decision was made to capture as much relevant insight as possible in the literature while acknowledging that adolescence is a period defined by puberty and rapid growth.

Comparators included studies where the intervention was compared to no service, placebo, or usual or standard care as defined by authors. Studies with no valid comparator were excluded, such as uncontrolled pre-post studies or uncontrolled pre-post with repeated cross-sectional design studies^3^. Eligible studies evaluated intervention effectiveness using one of the following designs: (i) randomised control trials, (ii) controlled before-after designs, (iii) cohort studies with comparison groups assessed under similar settings, and (v) non-equivalent groups, post-test only design studies with a clearly defined comparison group. Additionally, studies were required to have a minimum sample size of 50.

Outcomes for this review were assessed across two broad dimensions. First, adolescent health knowledge and perceptions as characterised by the Health Belief Model [31], and second, health and well-being using the five broad domains in the WHO framework, including (i) good health and optimum nutrition, (ii) safety and a supportive environment, (iii) connectedness, positive values and contribution to society, (iv) learning, competence, education, skills and employability, and (v) agency and resilience, supplemented with sub-domains in the Global Action for Measurement of Adolescent Health (GAMA) framework [20,32].

### Study records

#### Data management

References returned from bibliographic databases and grey literature were downloaded into EndNote by J.C. for organisation and deduplication. Titles and abstracts were then uploaded to the screening software Rayyan to facilitate efficient and blinded screening by reviewers.

#### Selection of studies

Titles and abstracts were screened by two independent reviewers (Y.L. and J.C.), working in a blinded manner, to identify potentially relevant studies [33]. For studies without full abstracts, they examined the full text to determine inclusion. Any discrepancies were resolved through discussion between Y.L. and J.C.. When consensus could not be reached, a third reviewer W.R. was consulted [33]. The second stage involved full-text review of studies identified as relevant in the initial screening. Y.L. and J.C. examined each full-text study to ensure adherence to the eligibility criteria. Any studies excluded at this stage were accompanied by documented justifications for exclusion. For the grey literature search, O.A. reviewed the titles, abstracts, or executive summaries of each study downloaded from grey literature sources. If studies lacked abstracts or executive summaries, the introduction and conclusion sections were examined to determine inclusion. The second stage of the grey literature review followed a similar process as the review of bibliographic studies described above. Finally, Y.L. conducted backward searches and assessed inclusion/exclusion for all studies. W.R. reviewed each decision at full-text screening stage, followed by discussion to reach agreement.

#### Data items and extraction

Y.L. and B.S. split the included studies for systematic data extraction using pre-prepared and piloted data extraction forms in Microsoft Excel. Since no study met our inclusion criteria from the grey literature, no data extraction was conducted. The data items we collected included study identification details, study characteristics, participant information, programme details, outcomes, effects, and gender related analysis. Details about the data items are reported in Appendix C. Reciprocal random checks of ten percent of studies extracted by Y.L. and B.S. identified no substantial discrepancy.

#### Quality Appraisal

The relevant Joanna Briggs Institute (JBI) tools were used to assess the quality of included randomised control trials, non-randomised studies of intervention (NRSIs), and cross-sectional studies [35] independently by Y.L. and B.S. Any discrepancies were resolved during discussion among the two assessors and W.R. Grading of Recommendations Assessment, Development and Evaluation (GRADE) was used to assess the certainty of evidence for each primary outcome [36–42] by Y.L. with revision and advice from B.S. and W.R.

#### Data synthesis and analysis

We first synthesised the programmatic design features of identified interventions and the number of studies by outcome. This included *a priori* categorisation by provider characteristics (lay vs. paraprofessional; peer- vs. adult-led) and setting (community vs. school), based on their relevance to effectiveness and to capture heterogeneity in the literature. Provider characteristics reflect differences in training, expertise, and social proximity, while setting captures delivery conditions. Qualitative evidence suggests that these dimensions influence engagement and implementation. Peer CHWs may improve engagement and access through relatability and trust [43–48], whereas adult CHWs may provide greater credibility and expertise but may be less acceptable due to privacy concerns [46,49,50]. Delivery context further shapes implementation. For example, home visits may support contextual understanding but raise privacy concerns, while logistical or safety constraints (e.g., travel to remote areas) may affect feasibility and effectiveness [49–51].

When at least three studies were comparable in terms of interventions, outcome definitions, and reported effect measures, we conducted univariate random-effect meta-analyses to obtain pooled effect estimates. Effect size conversions were performed independently by W.R. and Y.L., with discrepancies resolved through discussion. Odds ratios were converted to risk ratios using the *MetaConvert* R package [52], and adjusted incidence rate ratios were converted to risk ratios following Hanley (2018) [53]. Main analyses used restricted maximum likelihood (REML) to estimate between-study variance, and the Hartung-Knapp-Sidik-Jonkman (HKSJ) method [54,55] to derive confidence intervals, thereby accounting for uncertainty in heterogeneity estimation and the small number of studies. Heterogeneity was assessed using the I² statistic, τ², and the Cochran Q-test. Robustness checks were conducted using alternative methods, including DerSimonian and Laird (DL) estimator for τ², and conventional Wald-type confidence intervals^4^ (Appendix J).

Dependency was addressed in accordance with Cochrane guidelines [33] and the accompanying Cochrane tutorial on handling multi-arm studies [57]. First, for the two studies with multiple intervention arms, the intervention groups were combined in one study [58]. In the other, the effect estimate from the CHW arm was selected because there was insufficient information to combine the intervention groups or appropriately split the control group, and the CHW arm most closely aligned with our prespecified definition of a CHW intervention [59]. Second, for the one study that reported separate effect estimates for urban and rural settings across two countries [60], country-specific estimates were derived by combining the urban and rural estimates using fixed-effect inverse-variance weighting. Sensitivity analyses examining alternative approaches to handling dependency were conducted for each outcome where relevant. Full details of the methods applied to each source of dependency and results are provided in Appendix J.

Where effect measures could not be harmonised, findings were synthesised narratively using vote-counting. Subgroup analyses by gender and socioeconomic status were prespecified but not feasible due to the limited number of studies per outcome. Relevant findings by gender are reported narratively.

## Results

### Literature search

The search yielded 5,003 records after duplicate removal (see Appendix D. PRISMA flowchart). Two independent reviewers screened all records, with a disagreement rate of 2% at the title and abstract screening stage. Following this, 122 articles were assessed for full-text eligibility. Of these, 87 were excluded, with reasons detailed in Appendix E, resulting in 35 included studies (34 quantitative and 1 mixed methods).

### Study characteristics

Among the 35 studies included, twenty-four studies were randomised controlled trials (RCTs). Eight studies used non-equivalent groups, post-test only design, of which two had propensity score matching (PSM)-related statistical adjustments. Three studies used controlled before-after design [61–63], of which two used panel data and difference-in-differences (DiD) methods [61,62] while one used repeated cross-sectional pre-post [63].

All were published in English. Five studies were in Western Africa, 13 in Eastern Africa and 15 in Southern Africa. Two studies spanned across multiple countries. Appendix F provided detailed information on included studies’ design, interventions, sample size, target populations and outcome addressed.

### Target populations

Thirteen studies examined adolescents within the age range of 10-19 years [64–74,62,63] with the minimum age no less than seven years old. Nineteen studies examined a wider age range (i) *either* including both adolescents and young adults with median age falling within the 10-19 age range and maximum age no greater than 24 [75–81] (ii) *or* including both children and adolescents with median age falling between 10-19 and minimum age no less than five years old [82,83,59,61,84,85,60], or (iii) *otherwise* providing age-disaggregated results for the adolescent group [43,86–89]. The remaining three studies referred to senior primary and secondary school students [58,90,91].

### CHW interventions and Comparators

We identified 15 peer-, 20 adult- and two peer plus adult CHW-delivered programmes/interventions, with 26 taking place in the community settings, 11 in school settings^5^. Among the evaluated interventions/programmes, peer CHWs were active in both rural and urban areas, with support base from non-governmental organisations (NGOs). They typically received only a few days of training. In contrast, adult CHWs primarily operated in rural areas, often receiving over a week of training, and were supported by both government and NGOs. Key design features of evaluated programmes are summarised in Figure 1.

**Figure 1.**
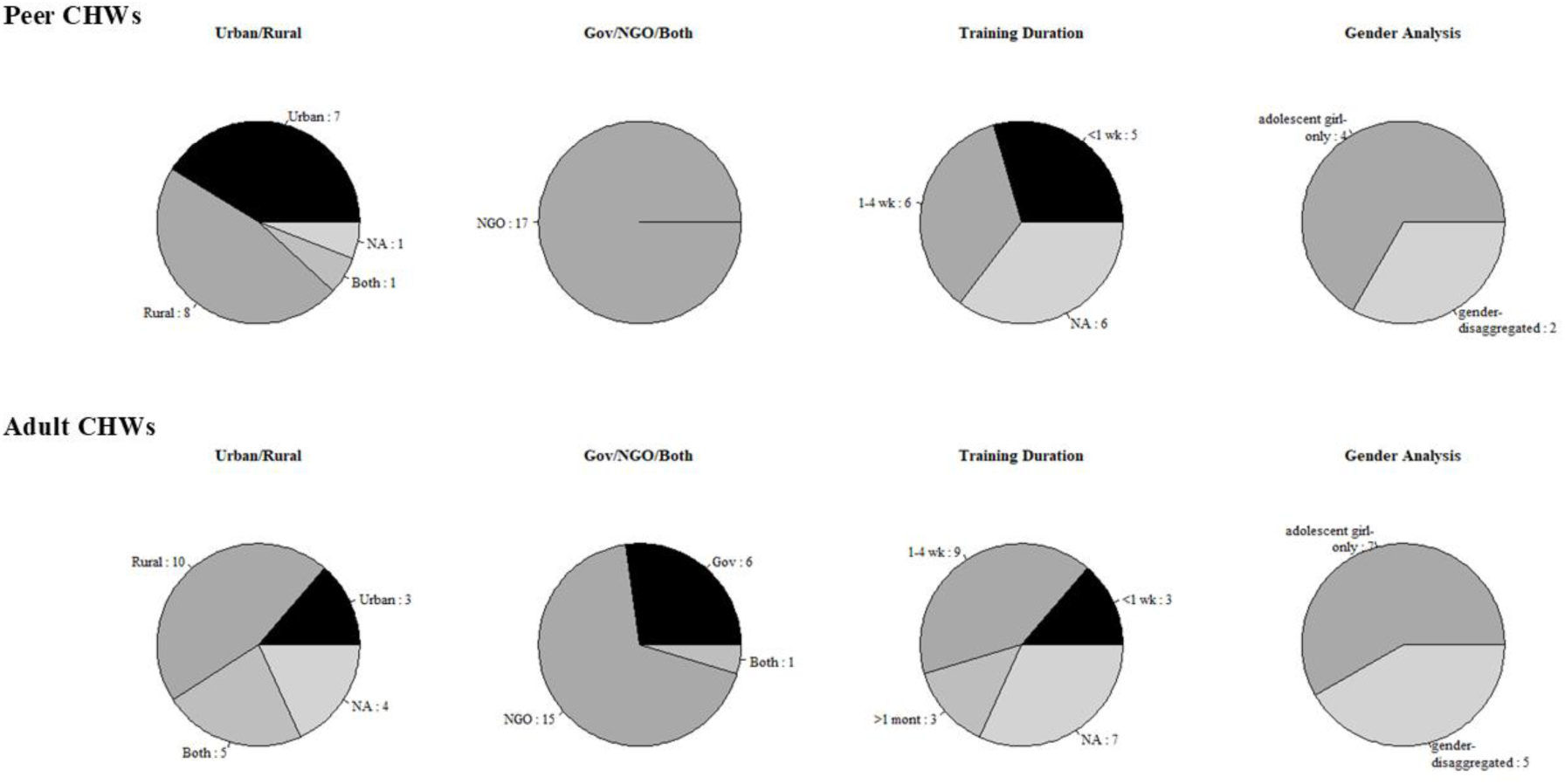
Programmatic design features of the evaluated CHW interventions.

The primary focus areas of evaluated programmes are summarised in Figure G.1 in Appendix G. Peer CHW interventions in community settings commonly centred on health education and awareness building, such as *Mzake* peer group intervention in Malawi [87]. Psychosocial support was exemplified by the Zvandiri programme’s peer-led counselling sessions in Zimbabwe [73,74,80]. They also involved service delivery, engagement, and linkages like the Yathu Yathu strategy, which featured community-based youth hubs staffed by peer support workers in Zambia [78], and Peer Educator-Based Refill of ART (PEBRA) model in Lesotho, which provided health-based differentiated service delivery using an ehealth application [79]. In school settings, peer CHWs typically delivered structured education sessions designed to improve knowledge and promote behavioural change. Examples included a behavioural change and health promotion programme focused on dietary diversity in Ethiopia [70].

Adult CHW interventions in community settings also covered a broad range of activities. These included the delivery of health education, promotion, and prevention messages, and psychosocial and mental health support such as cognitive behavioural therapy (CBT)-based interventions in Kenya, Sierra Leone, Tanzania and Zambia [60,75,76,85]. Adult CHWs also played a role in facilitating access to care, as seen in the *relais communautaires* model in Niger [64], and provided direct services and social support such as HEWs in Ethiopia [61,68]. Six studies examined adult CHW programmes implemented in school settings including groups of adolescents and their caregivers to address behavioural problems and mental health issues in Uganda [59,83], psycho-education group sessions focused on mental well-being [82], group education sessions focused on HIV and sexual and reproductive health and rights (SRHR)-related topics in Malawi [72], Nigeria [43] and Kenya [58], and sanitary pad distribution in Kenya [58].

Two programmes integrated both peer and adult CHWs, including participatory group education sessions on HIV prevention and sexual health in Nigeria [43], and support sessions to mitigate social isolation and stigma of adolescent motherhood in Zimbabwe [62].

### Outcomes

The identified evidence spanned 12 outcome domains, highlighting the broad potential of CHW-led interventions to support diverse aspects of adolescent health and wellbeing. Within these domains, studies focused on three key subdomains, which were (i) health knowledge, attitude and perceptions, (ii) health behaviours and outcomes, and (iii) socioeconomic well-being, summarised in Figure 2.

**Figure 2.**
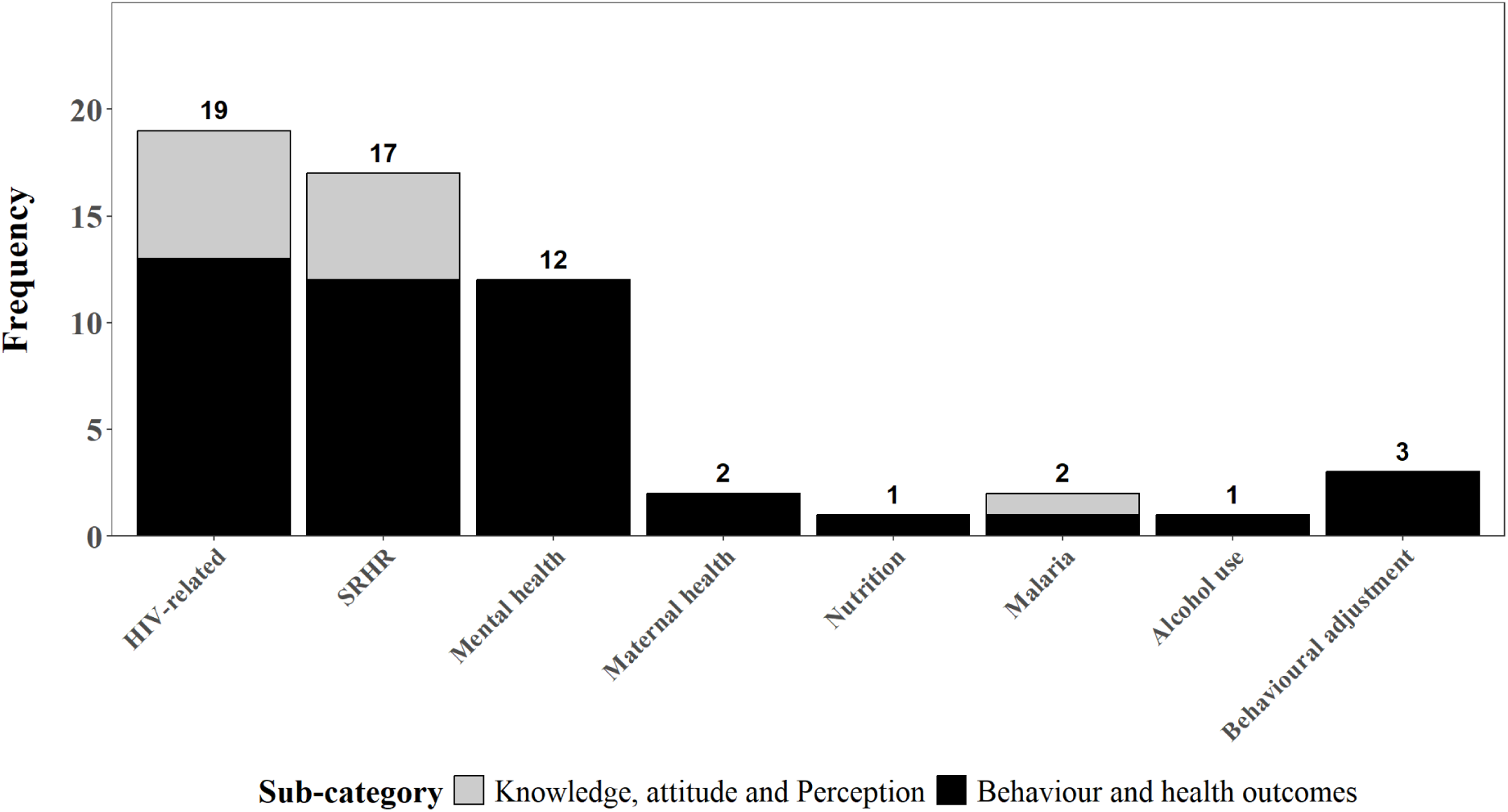
Outcome domains across CHW-led adolescent health interventions, by type. **Note:** The count numbers are at the intervention level rather than at study level. This review also summarised evidence for the role of CHWs in adolescent well-being in Appendix K. The number of the sub-categories for well-being outcomes were as follow. Safe and supportive environments included intimate partner violence (IPV) (n= 2), child labour (n=1), child marriage (n=1), (perceived) social support from family, peers and community (n=5), gender norms (n=1), and family functioning (n=1). Connectedness included healthy relationships (n=3). Learning, education and employment included school attendance, completion and retention (n=3), literacy, numeracy and academic performance (n=4), and labour/employment and earnings (n=1). Agency and resilience included self-efficacy (n=3), self-esteem (n=1), and hope (n=1).

### Risk of bias and Confidence in cumulative evidence

Of the 24 RCTs, 23 were rated as having a moderate risk of bias, and one was rated as having a high risk of bias, based on the JBI checklist for RCTs [92]. Among the 10 NRSIs, one was judged to have a high risk of bias, six a moderate risk, and three a low risk, according to the JBI checklist for NRSIs [93]. One cross-sectional study was evaluated as low risk using the JBI checklist for analytical cross-sectional studies [94].

We conducted a GRADE assessment for the seven meta-analysed outcomes. Certainty of evidence was rated as low for HIV viral load suppression, PTS, and depression, and very low for knowledge and use of modern contraceptives, unmet need for contraception, and adolescent pregnancy. We identified some concerns regarding publication bias for modern contraceptive use, but no serious concerns for depression, while the remaining outcomes were considered uncertain. Given the small number of effect estimates, funnel plots should be interpreted with caution. Full details of the risk-of-bias assessments, and GRADE justifications were provided in Appendix H.

### Data analysis

We presented vote-counting results for each outcome (see Appendix I for a summary) and reported pooled analyses in forest plots based on harmonised risk ratios or standardised mean difference when applicable. The focus of results in our review was on (i) knowledge, attitude and perception, (ii) health behaviours and outcomes and (iii) well-being, with results for well-being presented in Appendix K.

### (i) Knowledge, attitude and perception

#### HIV-related

Seven studies assessed HIV-related knowledge, attitude, perception, HIV-status awareness, and service-related attitudes. Evidence was mixed for HIV knowledge (5/7 positive) [43,72,87], and attitude towards HIV (2/3 positive) [43,76]. A positive effect was observed for knowledge of HIV status [78] but evidence is scant.

#### SRHR-related

Five studies evaluated CHW-led SRHR-related interventions, mainly group-based education delivered by female or gender-matched CHWs, covering contraception, sexually transmitted infections (STIs), fertility, pregnancy, menstruation, and SRHR-service awareness. A meta-analysis found no evidence of an effect on knowledge of modern contraceptives or condom use (RR: 0.99, 95% CI: 0.76-1.30, I^2^=88.16%, Figure M.1 in Appendix M [58,63]). Evidence was also consistently null for fertility knowledge [43,68] and SRHR-related services awareness^6^ [63]. Findings on attitudes were mixed: attitude towards modern contraceptives and condoms showed inconsistent effects (2/6 positive, 1/6 negative, and 3/6 null) [63,67], while attitude towards menstrual health was more favourable but with limited evidence (1/1 positive) [58].

#### Malaria-related

Evidence was limited to a single study, which reported improvements in knowledge of and attitude towards malaria [90]. The intervention also increased perceived vulnerability but reduced perceived disease severity^7^ [90].

### (ii) Health behaviour and outcomes

#### HIV-related

Thirteen studies examined CHW roles in HIV-related behaviour and health outcomes, with interventions often focused on treatment support and service delivery through home visits, reminders and referrals. CHWs showed promise in HIV testing services (4/5 positive effects [72,81,88,89]). There was also weak evidence of improved viral suppression (meta-analysis of six effects [73,74,76,79,80,84]: RR: 1.16; 95% CI: 0.99, 1.36; I² = 45.86%; Figure 3). In contrast, effects on ART adherence were weaker (2/7 positive [71,76]), and no improvements were observed for linkage to or retention in care (0/6 positive [74,79,84]). Evidence on other outcomes, such as voluntary medical circumcision (VMMC) uptake [78,81] and patient satisfaction [79], was limited and inconsistent.

**Figure 3.**
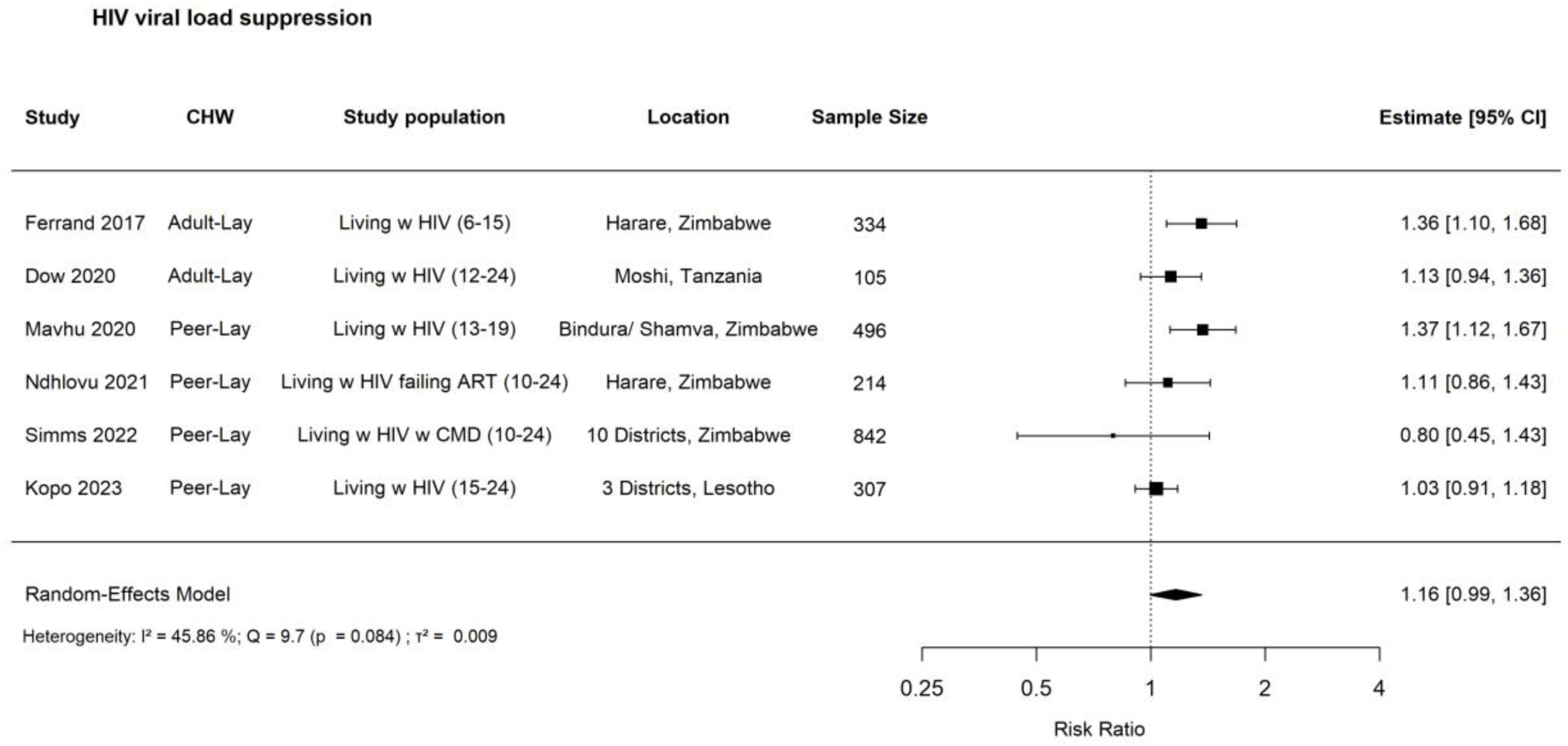
Forest plot: risk ratio for the effects of CHW-led interventions on HIV viral load suppression among adolescents in SSA **Note:** Six of the eight identified effect sizes were included. In both Ferrand et al. (2017) [84] and Kopo et al. (2023) [79], two effect sizes were reported, but only one from each study was retained. The excluded effect size in Ferrand et al. (2017) [84] was based on a composite outcome that extended beyond viral load suppression, while the excluded effect size in Kopo et al. (2023) [79] used a cutoff of <20 copies/mL, which differed substantially from the 1,000 or 400 copies/mL thresholds applied in our analysis. Visual inspection of the funnel plot (see Appendix H.2) suggests possible asymmetry as reflected by the lack of small-sample studies that reported negative results on HIV viral load suppression. While this may be an indication of publication bias, the small number of included studies (n=6) means this evidence may not be conclusive. Details about publication bias, heterogeneity, and imprecision for this outcome can be found in Appendix H.

#### SRHR-related

Ten studies assessed SRHR behaviour and health outcomes. Interventions were diverse, including education sessions (primarily peer-led), household visits (exclusively adult-led) and service delivery. Meta-analyses showed CHWs improved modern contraceptive or consistent condom use among adolescent girls (RR: 1.50, 95% CI: 1.13-1.99, I^2^ = 87.83%; Figure 4 [63–65,69,72,91]), but had null effects on unmet need for contraception (RR: 0.94, 95% CI: 0.68–1.29, I^2^ = 0.01%; Figure M.2 in Appendix M [63,69,78]) or adolescent pregnancy (RR: 0.92, 95% CI: 0.23–3.77, I^2^ = 90.64%; Figure M.3 in Appendix M [63,68,78]). Vote-counting suggested more consistent benefits for menstrual product use and menstruation management [58,77], and SRH service coverage [81], whereas no effects emerged for met material need of menstrual product [77] and the number of sexual partners [72]. Mixed effects were observed on sexual self-efficacy [63,72].

**Figure 4.**
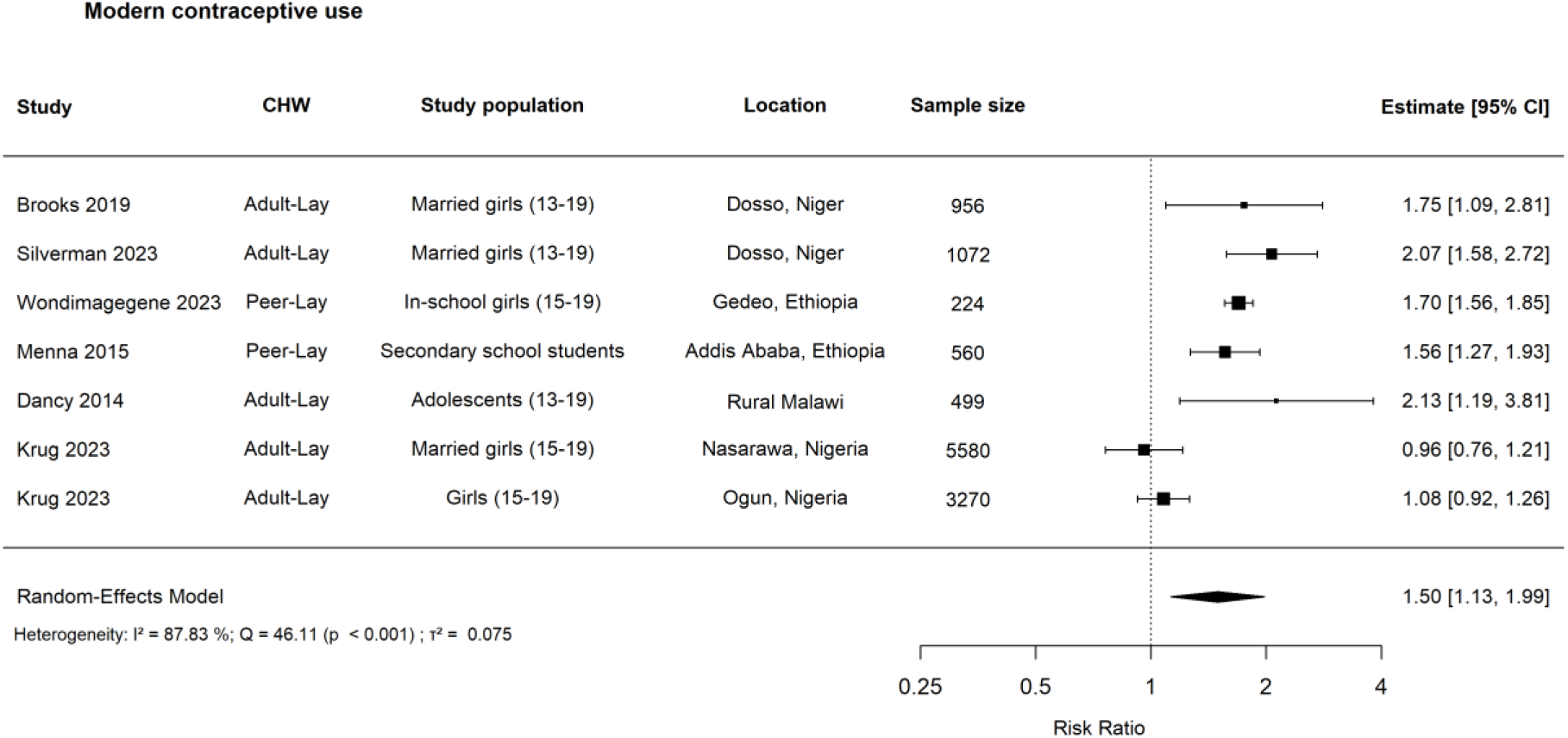
Forest plot: risk ratio for the effects of CHW-led interventions on modern contraceptive use among adolescents in SSA. **Note:** Seven of 15 effect sizes were included. The other effect sizes were excluded because they only focused on a specific modern contraceptive [63], or they only counted “ever use” rather than “current use” or “consistent use” [63,72,78]. The effect sizes included in the meta-analysis above reflected current use of modern contraceptives or consistent condom use in the last 2 or 12 months. Visual inspection of the funnel plot (see Appendix H.2) suggests some asymmetry, with a greater concentration of studies on one side of the pooled estimate and a relative absence of small-sample studies reporting null or negative results. While this may indicate potential publication bias, the limited number of studies and possible heterogeneity across settings and interventions mean that this evidence may not be robust. We still downgraded by one level to reflect potential risk. Details about publication bias, heterogeneity, and imprecision for this outcome can be found in Appendix H.

#### Mental health

Twelve studies, mostly adapted CBT using group modality, examined CHWs’ roles in adolescents’ mental health outcomes. Meta-analyses showed no evidence of protective effects in reducing trauma/stress-related symptoms (SMD: −0.58, 95% CI: −1.54, 0.38, I^2^ = 97.55%; Figure M.4 in Appendix M [60,75,76,82,85]), but moderate evidence of reducing adolescent depression (SMD −0.31, 95% CI: −0.61, 0.00, I^2^ = 84.94%; Figure 5 [59,60,62,66,73,76,82]). Mixed evidence was reported for grief (1/2 [60]), and functional impairment (3/5 [59,75,83]). No evidence was found for emotional/behavioural symptoms (0/5 [60,75]).

**Figure 5.**
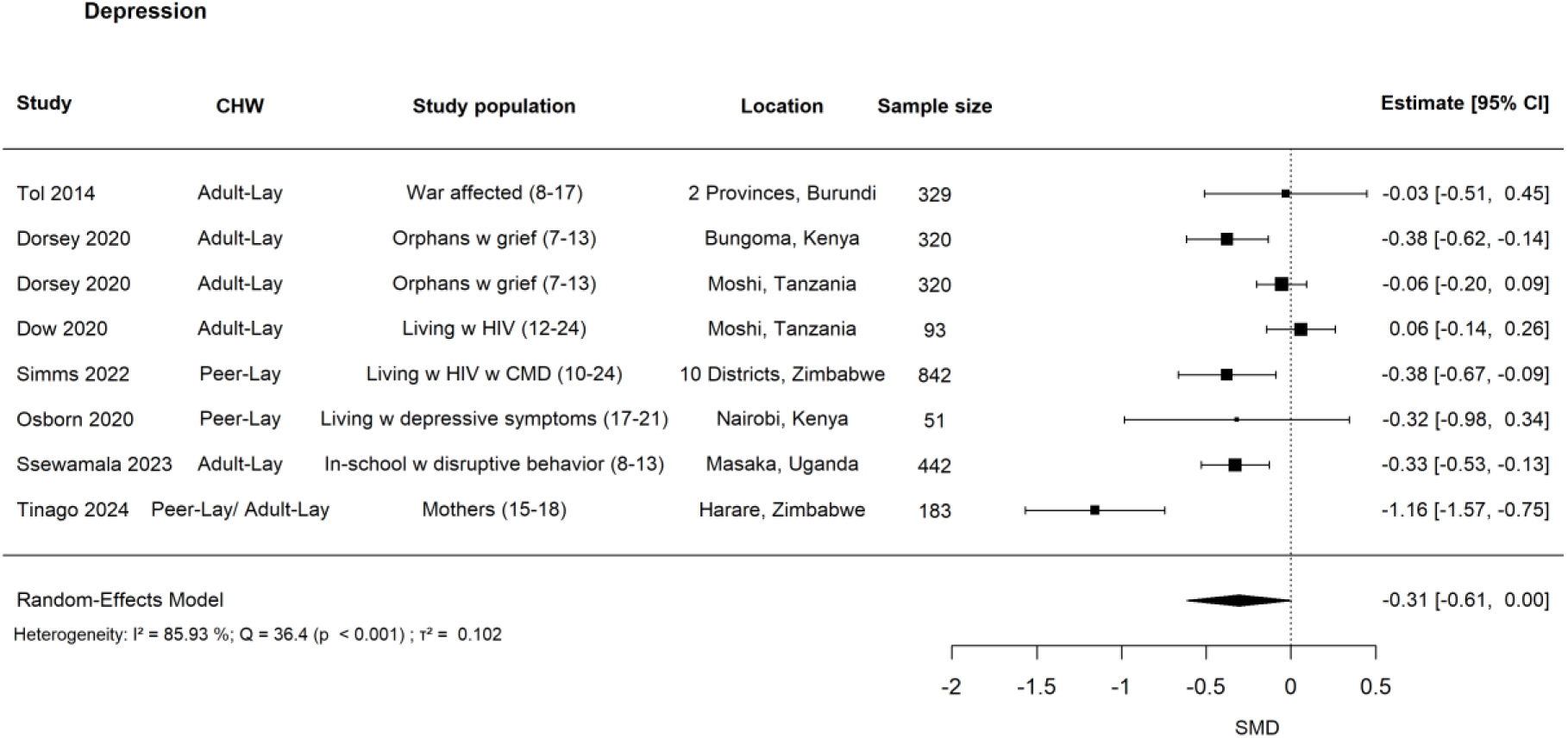
Forest plot: standardised mean difference (SMD) for the effects of CHW-led interventions on depression among adolescents in SSA. **Note:** Eight out of 15 effect sizes were included in the meta-analysis presented above. In the main analysis here, consistent with the approach for PTS, we combined urban and rural estimates within Kenya and Tanzania in Dorsey et al. (2020) [60] using fixed-effect inverse-variance weighting. Similarly, we prioritised child self-reports over parent reports, resulting in the exclusion of four guardian-reported measures from Dorsey et al. (2020) [60]. Additionally, one adjusted prevalence ratio based on a binary depression outcome was excluded [74]. Finally, to avoid dependence and double counting, we retained the effect estimate from the CHW-led intervention group in Ssewamala et al. (2023) and excluded the parent peer intervention group [59]. We are aware of the limitation of this approach, that is, it lost valuable information. However, we could neither combine treatment arms nor partition the shared control group as recommended following the Cochrane tutorial [57] due to insufficient information available. Nevertheless, we provided alternative results when including both effect estimates and using multilevel meta-analysis to test for robustness in Table J.7 in Appendix J. Visual inspection of the funnel plot for depression (see Appendix H.2) shows that although some variation appeared to exist in the effect estimates from relatively larger studies, the effect estimates still quite consistently pointed to a beneficial effect of the task-shifting strategy to CHWs. The consistency of positive effects in large trials, combined with the symmetry of the funnel plot around the pooled effect, tended to suggest a lower degree of publication bias. Details about publication bias, heterogeneity, and imprecision for this outcome can be found in Appendix H.

#### Other areas

In Uganda, two studies of the multifamily group programme for oppositional defiant disorder reported improvements immediately post-treatment but no sustained effects at week 8 follow-up [59,83]. The remaining outcomes were examined in one single study only. Evidence suggests positive effect on pro-social behaviour in Sierra Leone [75], improved dietary diversity [70], and insecticide-treated bed net use [90] in Ethiopia. No effect was observed on low/no alcohol use among Ethiopian adolescent boys [68].

### Gender analysis

Eight studies reported sex-disaggregated outcomes, revealing notable patterns, particularly in HIV-related outcomes. Across contexts, adolescent boys tend to show greater improvements than girls. In Zambia, boys experienced larger gains in knowledge of their HIV status following peer CHW-led youth hubs that offered community-based HIV testing [78]. Similarly, in Malawi, a 12-month HIV self-testing campaign increased lifetime HIV testing for both sexes, with stronger effects among boys [89]. Evidence from another Malawian study further suggests heterogeneity among girls: adult CHW-led, school-based education reduced HIV risk among boys of all ages and older adolescent girls (aged 16-19), but not among younger girls (aged 13-15), highlighting the need for more tailored interventions for younger female adolescents [72]. In Zimbabwe, a community adolescent treatment supporters (CATS)-led HIV intervention, comprising enhanced home visits and reminders, reduced viral load among male adolescents but not females [74].

Nine unique studies focused exclusively on adolescent girls, covering SRHR [58,63–65,67,69], maternal care [86], menstruation [58,77] and adolescent motherhood [62]. Although findings were sometimes inconclusive, the overall direction of effects was generally positive, particularly for gendered outcomes such as modern contraceptive use, maternal health, and menstrual practices. No study focused solely on adolescent boys.

Three studies examined gender-matched group interventions^8^, including those targeting contraceptive knowledge and use [63,65] and support for war-affected youth [75]. In addition, six studies employed female CHWs^9^ [58,61–63,67,68], primarily targeting girls across domains like menstruation [58], child marriage [68], modern contraceptive [67], and adolescent mothers [62]. These interventions produced mixed, but generally promising, outcomes.

### Sensitivity analysis

To assess the robustness of our findings, we conducted a series of sensitivity analyses, including the use of alternative estimators for between-study variance, alternative methods for confidence interval estimation, and the inclusion of uncontrolled pre–post studies that met all eligibility criteria except for the comparator requirement. We further examined robustness by applying alternative approaches to handle dependency. Full details are provided in the summary tables in Appendix J^10^.

Overall, the results were highly stable across tests for knowledge and use of modern contraceptives, unmet need, adolescent pregnancy, PTS, and HIV viral load suppression. In contrast, the estimated effect on depression showed some sensitivity to model specification when multilevel meta-analysis was used to account for multiple treatment arms.

## Discussion

### Findings and Contributions

This review provided a comprehensive synthesis of CHW-led interventions for adolescent health and well-being between 2014 and 2024, covering diverse outcomes. Findings were promising but varied by type, intensity and context of interventions, underscoring the need for more rigorous evaluations.

From a good number of RCTs evaluating task-shifting trauma-informed CBT to lay CHWs, we found moderate evidence that CHWs can reduce adolescent depressive symptoms, largely confirming and extending earlier reviews in low- and middle-income countries (LMICs) [96]. Our adolescent-specific focus reinforces CHWs’ potential to bridge mental health service gaps. There was weak evidence on CHWs-led programmes on HIV viral load suppression particularly Zimbabwe’s Zvandiri, although the certainty of evidence is low and replication in other contexts is needed. This extends prior reviews by focusing specifically on adolescents [97].

Evidence suggests a strong and robust positive trend in modern contraceptive and condom use among adolescent girls, particularly through education-based home visits [64,65] and peer-led school education sessions [69], contrasting findings from prior reviews [98]. However, unmet need for contraception, typically defined as adolescent girls who wish to delay or avoid pregnancy but are not using contraceptives, and adolescent pregnancy rates remained unchanged, potentially highlighting gaps between demand and supply. Many interventions focused on demand creation by improving knowledge and awareness with largely consistent positive effects, such as [65,69,72,91], whereas fewer examined CHWs’ direct provision and delivery of SRHR services [78]. Strengthening CHW provision of SRH services, alongside demand-side support, may better align contraceptive uptake with fertility preferences, thereby reducing unmet need and unwanted pregnancy.

Structural and cultural barriers may also shape these outcomes. Biases against premarital sexual activity discourage unmarried girls from accessing contraceptives [98], and persistently high child marriage alongside pronatalist norms^11^ limit progress in reducing unmet need and pregnancy particularly among the newly married, nulliparous adolescents [15]^12^. These contrasting effects might be because the pooled findings for contraceptive use versus unmet need and pregnancy derived from distinct sets of studies and contexts. Further research is needed to explain the heterogeneous impacts of CHW-led interventions across sociocultural settings, and to understand the influence of these contextual factors on the effectiveness of CHWs’ activities and engagement with adolescents.

Another possible explanation is the gendered power imbalances in sexual relationships and limited self-efficacy, which can prevent girls from using contraceptives or condoms even when desired. Although some evidence suggests CHWs can improve sexual self-efficacy, findings were mixed^13^. Moreover, only one study examined CHW-led effects on power dynamics and gender norms, finding that reproductive health education addressing gender and power improved some aspects [58], consistent with a prior review [99], but not all [58], highlighting the difficulty of shifting entrenched dynamics and the potential value of broader community-level change combined with gender- and power-focused education.

Taken together, while these structural, cultural, and relational mechanisms provide plausible explanations for the observed effects of CHWs on modern contraceptive use, the findings should be interpreted in light of the generally low quality of the existing evidence, highlighting the need for more rigorous research in this area.

No conclusive evidence emerged that CHWs improved adolescents’ contraceptive knowledge. Most studies were in context with high baseline knowledge limiting measurable gains, with studies of moderate risk of bias and inconsistent results^14^. Baseline knowledge, ranging from 0.44 to 0.82, again suggest possible ceiling effects. While the lack of evidence on CHW’s roles in improving SRHR knowledge is consistent with a prior evidence review [98], we suggest that more rigorous research is needed to assess CHWs’ role in achieving further improvements in adolescents’ HIV and modern contraceptive knowledge across SSA.

CHWs also appear promising in areas, such as menstruation management, nutrition, behavioural adjustment, although evidence is limited for robust conclusions. Moreover, research on adolescent maternal health has declined since 2014 and the focus has shifted to behaviours rather than health outcomes, but promising findings remain consistent with broader literature [100]. Safe abortion access remains unaddressed [101].

Girl-focused interventions and female CHWs often yielded positive results, especially in gendered outcomes such as menstruation management. Evidence on how CHW age affects intervention effectiveness in SSA remains mixed and inclusive following previous literature [102,103]. Our review could not compare peer versus adult CHW-led interventions due to limited data. Intergenerational models where CHWs of different ages work together (e.g., peer and adult CHWs in our review), seen in Ethiopia [63], Nigeria [43] and Zimbabwe [62], showed potential but lacked detailed insights into the collaboration dynamics. As such, we could not expand on the sparse literature exploring how generational factors like hierarchy and tension, noted in studies from South Africa, shape CHW delivery and outcomes [104].

Finally, CHW effectiveness depends not only on CHW delivery but also on intervention design^15^, and the availability and quality of facility/hospital-based care as interventions often involved referrals and community-facility linkages (see Appendix F and G). In seven multi-component interventions where CHWs are one element [58,63,77,78,81,86,90], attributing effects specifically to CHWs is less clear, and findings should therefore be interpreted with some caution^16^.

### Future policy and research recommendations

Moderate evidence supported low-intensity psychological interventions delivered by CHWs for adolescents experiencing trauma, isolation or stigma. Scaling such models, especially for war-affected youth, adolescent mothers and adolescents living with HIV, should be a priority. Sustaining impact requires integration into national systems, clear role definitions, workload assessments, and support for CHWs’ own mental health. Notably, no intervention targeted boys or LGBTQ+ adolescents was evaluated, highlighting gaps for future research and programming.

Further efforts are also needed to ascertain CHWs’ roles in improving adolescents’ HIV viral load suppression and to develop better measures for HIV treatment adherence. There is also a need to identify programme designs that can effectively meet adolescents’ need for modern contraceptives and reduce adolescent pregnancy. In addition, research should clarify how CHWs influence knowledge, attitudes, and perceptions among hard-to-reach adolescents, and whether these mediate health behaviours and outcomes. As more studies become available, meta-regression analyses should examine programme design elements (e.g., CHW age/gender, training duration, incentives, urban/rural context) and recipient characteristics (e.g., gender, socioeconomic status) to understand how best to motivate CHWs and enhance effectiveness. Finally, efforts are needed to equip CHWs to deliver inclusive, trust-based services, especially for marginalised adolescents.

Finally, greater attention is needed on under-researched outcomes such as adolescent nutrition, non-communicable diseases and injury prevention, which are critical for shaping lifelong health [3].

### Limitations

Despite its significant contributions, this review has several limitations across four main areas. First, regarding the search strategy, although comprehensive, our approach prioritised worker-related terms (e.g., peer educators, peer counsellors, peer supporters) over intervention mode-related terms (e.g., peer-led), which may have led to the omission of relevant studies that did not mention worker-specific terminology in the abstract. Second, in terms of data analysis, evidence quality posed significant challenges. Vote-counting provides limited inferential value, and meta-analyses were only feasible for seven outcomes due to inconsistent measurement and limited data. The process of transforming effect estimates to risk ratios relied on key assumptions that may introduce bias, including: using control group baseline risks to estimate risk ratios assumes similar covariate distributions across study groups; (even more strongly) unadjusted baseline risks^17^ were used for conversion despite most studies reporting adjusted effect measures, assuming similar covariate distribution across each participant; and adjusted prevalence ratios reported in two studies [74,78] were treated as equivalent to risk ratios. Substantial heterogeneity across interventions, study designs, and outcome measures further limited valid comparisons. In particular, high heterogeneity for PTS, depression, and adolescent pregnancy, combined with the limited number of studies, increases uncertainty in estimating the variance of the pooled estimates under HKSJ method. These meta-analysis findings should therefore be interpreted with caution. Third, cost and incentive data were rarely reported, constraining assessment of financial sustainability and optimal compensation models. Finally, we did not include French-language search terms, which may have led to the omission of relevant studies, particularly from Francophone West Africa.

### Conclusion

This review found evidence supporting CHWs’ potential to improve multiple areas of adolescent health and well-being in SSA. Strong evidence supports CHW-led interventions for improving contraceptive uptake, moderate evidence for CHW-led CBT for adolescent mental health, and weak evidence for HIV viral load suppression. Evidence across other domains remains limited. These findings support continued investment in CHWs and their integration into national health systems, alongside the need for more rigorously designed programmes to optimise targeting, delivery and effectiveness.

## Data Availability

All data produced in the present study are available upon reasonable request to the authors

## Acknowledgements

We had multiple group meetings with members from UNICEF. We would like to express our sincere gratitude to Alinane Kamlongera, Damilola Walker, Fatu Wurie, Joanna Lai, Lauren Galvin, Lauren Rumble, Maureen Kerubo Momanyi, Megan Christensen, Theresa Cerveau, Yi-Chen Jenny Han, Zeinab Hijazi (in alphabetical order) for joining regular research update meetings and providing detailed feedback for research findings.

## Author contributions

Yangxu Lin: Conceptualisation, data curation, methodology, formal analysis, writing – original draft, review & editing; Oluwatoni Adesina: Conceptualisation, data curation, methodology, formal analysis, writing – review & editing; Broline Sagini Asuma: Conceptualisation, data curation, methodology, formal analysis, writing – review & editing; Jenny Chen-Charles: Conceptualisation, data curation, methodology, formal analysis, writing – review & editing; Satvika Chalasani: Conceptualisation, writing – review & editing; Lucie Cluver: Conceptualisation, writing –review & editing; William E. Rudgard: Conceptualisation, funding acquisition, formal analysis, project administration, supervision, writing – original draft, review & editing.

## Abbreviations

AIDS, Acquired Immune Deficiency Syndrome; ANC, Ante-natal Care; ART, Antiretroviral Therapy; BASE, Bielefeld Academic Search Engine; CATS, Community Adolescent Treatment Supporters; CBCL, Child Behaviour Checklist; CBT, Cognitive Behavioural Therapy; CI, Confidence Interval; cRCT, Clustered Randomised Controlled Trial; CATS, Community Adolescent Treatment Supporter; CHW, Community Health Worker; CIAO, Colombia International Affairs Online; DFID, Department for International Development; DiD, Difference-in-Differences; DL, DerSimonian and Laird; GAMA, Global Action for Measurement of Adolescent Health; GRADE, Grading of Recommendations, Assessment, Development and Evaluations; HB, Hepatitis B; HEW, Health Extension Worker; HIV, Human Immunodeficiency Virus; HKSJ, Hartung-Knapp-Sidik-Jonkman; IFPRI, International Food Policy Research Institute; ITN, Insecticide-Treated Net; IPV, Intimate Partner Violence; JBI, Joanna Briggs Institute; LMICs, Low- and Middle-Income Countries; MD, Mean Difference; NGOs, Non-governmental Organisations; NRSIs: Non-randomised studies of intervention; ODD, Oppositional Defiant Disorder; OR, Odds Ratio; OIS, Optimal Information Size; OPM, Oxford Policy Management; PATH, Pathfinder International Programme for Appropriate Technology in Health; PEBRA, Peer Educator Based Refill of ART; PHQ-9, Patient Health Questionnaire-9; PICO, Population, Intervention, Comparator and Outcome; PrEP, Pre-Exposure Prophylaxis; PSI, Population Services International; PSM, Propensity Score Matching; PST, Problem-Solving Therapy; PRISMA, Preferred Reporting Items for Systematic Reviews and Meta-analyses; PTSD, Post-Traumatic Stress Disorder; PTS, Post-Traumatic Stress; R4D, Results for Development; RCT(s), Randomised Controlled Trial(s); REML, Restricted Maximum Likelihood; RR, Risk Ratio; SMD, Standardised Mean Difference; SSA, sub-Saharan Africa; SRH, Sexual and Reproductive Health; SRHR, Sexual and Reproductive Health and Right; STI, Sexually Transmitted Infection; TB, Tuberculosis; UNICEF, United Nations International Children’s Emergency Fund; VMMC, Voluntary Medical Male Circumcision; WHO, World Health Organisation.

## Appendices

## Appendix A. Search Strategy

### Appendix A.1 Databases and Platforms Searched for Grey Literature

Google Scholar, Last Mile Health, Population Council, FHI 360, Population Services International (PSI), Pathfinder International Program for Appropriate Technology in Health (PATH), Pathfinder International, John Snow Inc, Eldis, UNICEF, WHO, World Bank, International Food Policy Research Institute (IFPRI), Oxford Policy Management (OPM), Overton, Colombia International Affairs Online (CIAO), and Bielefeld Academic Search Engine (BASE).

### Appendix A.2 Search Terms for All Bibliographic Databases

#### A.2.1 Scopus (Elsevier)

1) TITLE-ABS-KEY (“adolescent” OR “youth” “young people” OR “young person” OR “teen” OR “teens” OR “teenager” OR “adolescents” OR “young women” OR “young men” OR “reproductive age” OR “15-24” OR “15-19”)

2) TITLE-ABS-KEY (“Community Health Worker*” OR “CHWs” OR “health volunteers” OR “CHV” OR “CHVs” OR “CHW” OR “Volunteer Health” OR “Village Health” OR “Lay Health” OR “Lay Counsel*” OR “Health Extension” OR “Agentes Polivalentes” OR “Village Malaria” OR “Nutrition Volunteer*” OR “Nutrition Worker” OR “Community Drug Distributor” OR “Community Reproductive Health” OR “Community Mobilizer” OR “Health Promoter” OR “Peer Counsellor” OR “Traditional Birth Attendant” OR “Health Extension Program*” OR “Community Health Volunteers” OR “Village Health Worker” OR “Ward-Based Primary Health Care Outreach Team*” OR “Health Care Outreach” OR “Paths to Primary Health Care” OR “Relais Communautaires” OR “Community health promoter” OR “Community health extension workers” OR “Village Health Teams” OR “Community health aide” OR “Community health promoters” OR “Linkage facilitators” OR “Health Extension worker” OR “Women’s development army” OR “Community health agents” OR “Trained traditional birth attendants” OR “Volunteer health promoters” OR “Community health officer” OR “Community health nurse” OR “Community health assistant” OR “Community health service supervisors” OR “Trained traditional midwives” OR “Household health promoters” OR “Community directed distributors” OR “Agents Communautaires de Nutrition” OR “Agents Communautaires” OR “Community Nutrition Workers” OR “Community Workers” OR “Community Health Nurses” OR “Midwives” OR “Medical Assistants” OR “Environmental Health Assistants” OR “Health Surveillance Assistants” OR “Agentes Polivalentes Elementares” OR “Basic Multipurpose Health Workers” OR “Agents de Santé Communautaire” OR “Relais Volunteers” OR “Relay Volunteers” OR “Volunteer Village Health Workers” OR “Community Health Aides” OR “Community Health Assistants” OR “Christian Health Association of Nigeria” OR “Animatrice de Santé Maternelle” OR “Binôme” OR “Volunteer Cell Coordinators” OR “Maternal Health Moderator” OR “Binomial” OR “Traditional birth attendants” OR “Community drug distributors” OR “Ward-Based Primary Health Care Outreach Team” OR “Environmental Health Officers” OR “Village Executive Officers” OR “Community-Based Volunteers” OR “Village health worker” OR “Raedat Refiat (village pioneer)” OR “Agents de la santé communautaire” OR “Agents de santé communautaires” OR “Matrones; bajenu gox” OR “Relais communautaires” OR “Relais Communautaire” OR “Agent Santé Communautaire” OR “Community Mobilizers” OR “Community counsellors” OR “Agent de Santé à Base Communautaire” OR “Community Health Agent” OR “Community relays” OR “Community health agents” OR “Village Health Workers” OR “Musharit (among Sudanese refugees)” OR “Cabillos” OR “Relais communautaires” OR “Relais de site” OR “Relais promotionnels” OR “Community based distributors” OR “Community national volunteers” OR “Home health promoters” OR “Community Volunteers” OR “Agents de Santé Communautaires” OR “Village Health Workers” OR “Leader – Pastoral da Criança” OR “Volunteer Community-based Maternal and Newborn Care” OR “TB/HIV Promoters” OR “Lifestyle Ambassadors” OR “Home-based Care Providers” OR “Health Education Assistants” OR “Community Health Home-based Care Volunteers” OR “Rural Health Motivators”)

3) “Africa” OR (“Africa”[MeSH]) OR Africa” OR Algeria OR Angola OR Benin OR Botswana OR “Burkina Faso” OR Burundi OR Cameroon OR “Canary Islands” OR “Cape Verde” OR “Central African Republic” OR Chad OR Comoros OR Congo OR “Democratic Republic of Congo” OR Djibouti OR Egypt OR “Equatorial Guinea” OR Eritrea OR Ethiopia OR Gabon OR Gambia OR Ghana OR Guinea OR “Guinea Bissau” OR “Ivory Coast” OR “Cote d’Ivoire” OR Jamahiriya OR Jamahiriya OR Kenya OR Lesotho OR Liberia OR Libya OR Libya OR Madagascar OR Malawi OR Mali OR Mauritania OR Mauritius OR Morocco OR Mozambique OR Mozambique OR Namibia OR Niger OR Nigeria OR Principe OR Reunion OR Rwanda OR “Sao Tome” OR Senegal OR Seychelles OR “Sierra Leone” OR Somalia OR “South Africa” OR “St Helena” OR Sudan OR Swaziland OR Tanzania OR Togo OR Tunisia OR Uganda OR “Western Sahara” OR Zaire OR Zambia OR Zimbabwe OR “Central Africa” OR “Central African” OR “West Africa” OR “West African” OR “Western Africa” OR “Western African” OR “East Africa” OR “East African” OR “Eastern Africa” OR “Eastern African” OR “North Africa” OR “North African” OR “Northern Africa” OR “Northern African” OR “South African” OR “Southern Africa” OR “Southern African” OR “sub Saharan Africa” OR “sub Saharan African” OR “sub-Saharan Africa”

#### A.2.2 PubMed

(Community Health Worker* OR CHWs OR health volunteers OR CHV OR Volunteer Health OR Village Health OR Lay Health OR Lay Counsel* OR Health Extension Worker* OR Village Malaria Worker* OR Nutrition Volunteer* OR Nutrition Worker* OR Community Drug Distributor* OR Community Reproductive Health Worker* OR Community Mobilizer* OR Health Promoter* OR Peer Counsellor* OR Peer Navigat* OR Peer Counsel* OR Peer Educat* OR Peer Support* OR Peer-to-Peer OR Traditional Birth Attendant* OR Community Health Volunteer* OR Village Health Worker* OR Ward-Based Primary Health Care Outreach Team* OR Relais Communautaire* OR Community Health Promoter* OR Community Health Extension Worker* OR Village Health Team* OR Linkage Facilitator* OR Women’s Development Army* OR Community Health Agent* OR Trained Traditional Birth Attendant* OR Volunteer Health Promoter* OR Community Health Officer* OR Community Health Nurse* OR Community Health Assistant* OR Community Health Service Supervisor* OR Trained Traditional Midwife* OR Household Health Promoter* OR Community Directed Distributor* OR Agent* Communautaire de Nutrition OR Community Nutrition Worker* OR Community Worker* OR Midwife* OR Medical Assistant* OR Environmental Health Assistant* OR Health Surveillance Assistant* OR Agentes Polivalentes Elementares OR Basic Multipurpose Health Worker* OR Agents de Sante Communautaire OR Volunteer Village Health Worker* OR Animatrice de Sante Maternelle OR Volunteer Cell Coordinator* OR Maternal Health Moderator* OR Community-Based Volunteer* OR Home Health Promoter* OR Raedat Refiat OR Musharit OR Cabillos OR Relais Promotionnel* OR Community-Based Distributor* OR Lifestyle Ambassador* OR Home-Based Care Provider* OR Health Education Assistant* OR Rural Health Motivator*)

AND

(adolescent* OR youth OR young people OR teen* OR teenager* OR young women OR young men OR reproductive age OR “15-24” OR “15-19”)

AND

(Africa OR Algeria OR Angola OR Benin OR Botswana OR Burkina Faso OR Burundi OR Cameroon OR Canary Islands OR Cape Verde OR Central African Republic OR Chad OR Comoros OR Congo OR Democratic Republic of Congo OR Djibouti OR Egypt OR Equatorial Guinea OR Eritrea OR Ethiopia OR Gabon OR Gambia OR Ghana OR Guinea OR Guinea-Bissau OR Ivory Coast OR Cote d’Ivoire OR Kenya OR Lesotho OR Liberia OR Libya OR Madagascar OR Malawi OR Mali OR Mauritania OR Mauritius OR Morocco OR Mozambique OR Namibia OR Niger OR Nigeria OR Rwanda OR Senegal OR Seychelles OR Sierra Leone OR Somalia OR South Africa OR Sudan OR Eswatini OR Tanzania OR Togo OR Tunisia OR Uganda OR Western Sahara OR Zambia OR Zimbabwe OR Central Africa OR West Africa OR East Africa OR North Africa OR Southern Africa OR sub-Saharan Africa)

#### A.2.3 Embase, Global Health, Medline, and PsycINFO (via Ovid)

(Community Health Worker* OR CHWs OR health volunteers OR CHV* OR Volunteer Health OR Village Health OR Lay Health OR Lay Counsel* OR Health Extension Worker* OR Village Malaria Worker* OR Nutrition Volunteer* OR Nutrition Worker* OR Community Drug Distributor* OR Community Reproductive Health Worker* OR Community Mobilizer* OR Health Promoter* OR Peer Counsellor* OR Peer Navigat* OR Peer Counsel* OR Peer Educat* Peer Support* OR Peer-to-Peer OR Traditional Birth Attendant* OR Community Health Volunteer* OR Village Health Worker* OR Ward-Based Primary Health Care Outreach Team* OR Relais Communautaire* OR Community Health Promoter* OR Community Health Extension Worker* OR Village Health Team* OR Linkage Facilitator* OR Women’s Development Army* OR Community Health Agent* OR Trained Traditional Birth Attendant* OR Volunteer Health Promoter* OR Community Health Officer* OR Community Health Nurse* OR Community Health Assistant* OR Community Health Service Supervisor* OR Trained Traditional Midwife* OR Household Health Promoter* OR Community Directed Distributor* OR Agent* Communautaire de Nutrition OR Community Nutrition Worker* OR Community Worker* OR Midwife* OR Medical Assistant* OR Environmental Health Assistant* OR Health Surveillance Assistant* OR Agentes Polivalentes Elementares OR Basic Multipurpose Health Worker* OR Agents de Sante Communautaire OR Volunteer Village Health Worker* OR Animatrice de Sante Maternelle OR Volunteer Cell Coordinator* OR Maternal Health Moderator* OR Community-Based Volunteer* OR Home Health Promoter* OR Raedat Refiat OR Musharit OR Cabillos OR Relais Promotionnel* OR Community-Based Distributor* OR Lifestyle Ambassador* OR Home-Based Care Provider* OR Health Education Assistant* OR Rural Health Motivator*).af.

AND

(adolescent* OR youth OR young people OR teen* OR teenager* OR young women OR young men OR reproductive age OR “15-24” OR “15-19”).af.

AND

(Africa OR Algeria OR Angola OR Benin OR Botswana OR Burkina Faso OR Burundi OR Cameroon OR Canary Islands OR Cape Verde OR Central African Republic OR Chad OR Comoros OR Congo OR Democratic Republic of Congo OR Djibouti OR Egypt OR Equatorial Guinea OR Eritrea OR Ethiopia OR Gabon OR Gambia OR Ghana OR Guinea OR Guinea-Bissau OR Ivory Coast OR Cote d’Ivoire OR Kenya OR Lesotho OR Liberia OR Libya OR Madagascar OR Malawi OR Mali OR Mauritania OR Mauritius OR Morocco OR Mozambique OR Namibia OR Niger OR Nigeria OR Rwanda OR Senegal OR Seychelles OR Sierra Leone OR Somalia OR South Africa OR Sudan OR Eswatini OR Tanzania OR Togo OR Tunisia OR Uganda OR Western Sahara OR Zambia OR Zimbabwe OR Central Africa OR West Africa OR East Africa OR North Africa OR Southern Africa OR sub-Saharan Africa).af.

#### A.2.4 CINAHL, ERIC, Africa-Wide Information (via EBSCO)

(TX(“Community Health Worker*” OR “CHWs” OR “health volunteers” OR “CHV” OR “CHVs” OR “CHW” OR “Volunteer Health” OR “Village Health” OR “Lay Health” OR “Lay Counsel*” OR “Health Extension” OR “Agentes Polivalentes” OR “Village Malaria” OR “Nutrition Volunteer*” OR “Nutrition Worker” OR “Community Drug Distributor” OR “Community Reproductive Health” OR “Community Mobilizer” OR “Health Promoter” OR “Peer Counsellor” OR “Peer Navigat*” OR “Peer Counsel*” OR “Peer Educat*” OR “Peer Support*” OR “Peer-to-Peer” OR “Traditional Birth Attendant” OR “Health Extension Program*” OR “Community Health Volunteers” OR “Village Health Worker” OR “Ward-Based Primary Health Care Outreach Team*” OR “Health Care Outreach” OR “Paths to Primary Health Care” OR “Relais Communautaires” OR “Community health promoter” OR “Community health extension workers” OR “Village Health Teams” OR “Community health aide” OR “Community health promoters” OR “Linkage facilitators” OR “Health Extension worker” OR “Women’s development army” OR “Community health agents” OR “Trained traditional birth attendants” OR “Volunteer health promoters” OR “Community health officer” OR “Community health nurse” OR “Community health assistant” OR “Community health service supervisors” OR “Trained traditional midwives” OR “Household health promoters” OR “Community directed distributors” OR “Agents Communautaires de Nutrition” OR “Agents Communautaires” OR “Community Nutrition Workers” OR “Community Workers” OR “Community Health Nurses” OR “Midwives” OR “Medical Assistants” OR “Environmental Health Assistants” OR “Health Surveillance Assistants” OR “Agentes Polivalentes Elementares” OR “Basic Multipurpose Health Workers” OR “Agents de Santé Communautaire” OR “Relais Volunteers” OR “Relay Volunteers” OR “Volunteer Village Health Workers” OR “Community Health Aides” OR “Community Health Assistants” OR “Christian Health Association of Nigeria” OR “Animatrice de Santé Maternelle” OR “Binôme” OR “Volunteer Cell Coordinators” OR “Maternal Health Moderator” OR “Binomial” OR “Traditional birth attendants” OR “Community drug distributors” OR “Ward-Based Primary Health Care Outreach Team” OR “Environmental Health Officers” OR “Village Executive Officers” OR “Community-Based Volunteers” OR “Village health worker” OR “Raedat Refiat (village pioneer)” OR “Agents de la santé communautaire” OR “Agents de santé communautaires” OR “Matrones; bajenu gox” OR “Relais communautaires” OR “Relais Communautaire” OR “Agent Santé Communautaire” OR “Community Mobilizers” OR “Community counsellors” OR “Agent de Santé à Base Communautaire” OR “Community Health Agent” OR “Community relays” OR “Community health agents” OR “Village Health Workers” OR “Musharit (among Sudanese refugees)” OR “Cabillos” OR “Relais communautaires” OR “Relais de site” OR “Relais promotionnels” OR “Community based distributors” OR “Community national volunteers” OR “Home health promoters” OR “Community Volunteers” OR “Agents de Santé Communautaires” OR “Village Health Workers” OR “Leader – Pastoral da Criança” OR “Volunteer Community-based Maternal and Newborn Care” OR “TB/HIV Promoters” OR “Lifestyle Ambassadors” OR “Home-based Care Providers” OR “Health Education Assistants” OR “Community Health Home-based Care Volunteers” OR “Rural Health Motivators”))

AND

(TX (“adolescent” OR “youth” OR “young people” OR “teen” OR “teens” OR “teenager” OR “adolescents” OR “young women” OR “young men” OR “reproductive age” OR “15-24” OR “15-19”))

AND

(TX (“Africa” OR “Africa*” OR “Algeria” OR “Angola” OR “Benin” OR “Botswana” OR “Burkina Faso” OR “Burundi” OR “Cameroon” OR “Canary Islands” OR “Cape Verde” OR “Central African Republic” OR “Chad” OR “Comoros” OR “Congo” OR “Democratic Republic of Congo” OR “Djibouti” OR “Egypt” OR “Equatorial Guinea” OR “Eritrea” OR “Ethiopia” OR “Gabon” OR “Gambia” OR “Ghana” OR “Guinea” OR “Guinea Bissau” OR “Ivory Coast” OR “Cote d’Ivoire” OR “Jamahiriya” OR “Kenya” OR “Lesotho” OR “Liberia” OR “Libya” OR “Madagascar” OR “Malawi” OR “Mali” OR “Mauritania” OR “Mauritius” OR “Morocco” OR “Mozambique” OR “Namibia” OR “Niger” OR “Nigeria” OR “Principe” OR “Reunion” OR “Rwanda” OR “Sao Tome” OR “Senegal” OR “Seychelles” OR “Sierra Leone” OR “Somalia” OR “South Africa” OR “St Helena” OR “Sudan” OR “Swaziland” OR “Tanzania” OR “Togo” OR “Tunisia” OR “Uganda” OR “Western Sahara” OR “Zaire” OR “Zambia” OR “Zimbabwe” OR “Central Africa” OR “Central African” OR “West Africa” OR “West African” OR “Western Africa” OR “Western African” OR “East Africa” OR “East African” OR “Eastern Africa” OR “Eastern African” OR “North Africa” OR “North African” OR “Northern Africa” OR “Northern African” OR “South African” OR “Southern Africa” OR “Southern African” OR “sub Saharan Africa” OR “sub Saharan African” OR “sub-Saharan Africa” OR “sub-Saharan African”))

#### A.2.5 Web of Science, Proquest Social Science, Premium, and EconLit (via SOLO)

(“Community Health Worker*” OR “CHWs” OR “health volunteers” OR “CHV” OR “CHVs” OR “CHW” OR “Volunteer Health” OR “Village Health” OR “Lay Health” OR “Lay Counsel*” OR “Health Extension” OR “Agentes Polivalentes” OR “Village Malaria” OR “Nutrition Volunteer*” OR “Nutrition Worker” OR “Community Drug Distributor” OR “Community Reproductive Health” OR “Community Mobilizer” OR “Health Promoter” OR “Peer Counsellor” OR “Peer Navigat*” OR “Peer Counsel*” OR “Peer Educat*” OR “Peer Support*” OR “Peer-to-Peer” OR “Traditional Birth Attendant” OR “Health Extension Program*” OR “Community Health Volunteers” OR “Village Health Worker” OR “Ward-Based Primary Health Care Outreach Team*” OR “Health Care Outreach” OR “Paths to Primary Health Care” OR “Relais Communautaires” OR “Community health promoter” OR “Community health extension workers” OR “Village Health Teams” OR “Community health aide” OR “Community health promoters” OR “Linkage facilitators” OR “Health Extension worker” OR “Women’s development army” OR “Community health agents” OR “Trained traditional birth attendants” OR “Volunteer health promoters” OR “Community health officer” OR “Community health nurse” OR “Community health assistant” OR “Community health service supervisors” OR “Trained traditional midwives” OR “Household health promoters” OR “Community directed distributors” OR “Agents Communautaires de Nutrition” OR “Agents Communautaires” OR “Community Nutrition Workers” OR “Community Workers” OR “Community Health Nurses” OR “Midwives” OR “Medical Assistants” OR “Environmental Health Assistants” OR “Health Surveillance Assistants” OR “Agentes Polivalentes Elementares” OR “Basic Multipurpose Health Workers” OR “Agents de Santé Communautaire” OR “Relais Volunteers” OR “Relay Volunteers” OR “Volunteer Village Health Workers” OR “Community Health Aides” OR “Community Health Assistants” OR “Christian Health Association of Nigeria” OR “Animatrice de Santé Maternelle” OR “Binôme” OR “Volunteer Cell Coordinators” OR “Maternal Health Moderator” OR “Binomial” OR “Traditional birth attendants” OR “Community drug distributors” OR “Ward-Based Primary Health Care Outreach Team” OR “Environmental Health Officers” OR “Village Executive Officers” OR “Community-Based Volunteers” OR “Village health worker” OR “Raedat Refiat (village pioneer)” OR “Agents de la santé communautaire” OR “Agents de santé communautaires” OR “Matrones; bajenu gox” OR “Relais communautaires” OR “Relais Communautaire” OR “Agent Santé Communautaire” OR “Community Mobilizers” OR “Community counsellors” OR “Agent de Santé à Base Communautaire” OR “Community Health Agent” OR “Community relays” OR “Community health agents” OR “Village Health Workers” OR “Musharit (among Sudanese refugees)” OR “Cabillos” OR “Relais communautaires” OR “Relais de site” OR “Relais promotionnels” OR “Community based distributors” OR “Community national volunteers” OR “Home health promoters” OR “Community Volunteers” OR “Agents de Santé Communautaires” OR “Village Health Workers” OR “Leader – Pastoral da Criança” OR “Volunteer Community-based Maternal and Newborn Care” OR “TB/HIV Promoters” OR “Lifestyle Ambassadors” OR “Home-based Care Providers” OR “Health Education Assistants” OR “Community Health Home-based Care Volunteers” OR “Rural Health Motivators”)

AND

(“adolescent” OR “youth” OR “young people” OR “teen” OR “teens” OR “teenager” OR “adolescents” OR “young women” OR “young men” OR “reproductive age” OR “15-24” OR “15-19”)

AND

(“Africa” OR “Africa*” OR “Algeria” OR “Angola” OR “Benin” OR “Botswana” OR “Burkina Faso” OR “Burundi” OR “Cameroon” OR “Canary Islands” OR “Cape Verde” OR “Central African Republic” OR “Chad” OR “Comoros” OR “Congo” OR “Democratic Republic of Congo” OR “Djibouti” OR “Egypt” OR “Equatorial Guinea” OR “Eritrea” OR “Ethiopia” OR “Gabon” OR “Gambia” OR “Ghana” OR “Guinea” OR “Guinea Bissau” OR “Ivory Coast” OR “Cote d’Ivoire” OR “Jamahiriya” OR “Kenya” OR “Lesotho” OR “Liberia” OR “Libya” OR “Madagascar” OR “Malawi” OR “Mali” OR “Mauritania” OR “Mauritius” OR “Morocco” OR “Mozambique” OR “Namibia” OR “Niger” OR “Nigeria” OR “Principe” OR “Reunion” OR “Rwanda” OR “Sao Tome” OR “Senegal” OR “Seychelles” OR “Sierra Leone” OR “Somalia” OR “South Africa” OR “St Helena” OR “Sudan” OR “Swaziland” OR “Tanzania” OR “Togo” OR “Tunisia” OR “Uganda” OR “Western Sahara” OR “Zaire” OR “Zambia” OR “Zimbabwe” OR “Central Africa” OR “Central African” OR “West Africa” OR “West African” OR “Western Africa” OR “Western African” OR “East Africa” OR “East African” OR “Eastern Africa” OR “Eastern African” OR “North Africa” OR “North African” OR “Northern Africa” OR “Northern African” OR “South African” OR “Southern Africa” OR “Southern African” OR “sub Saharan Africa” OR “sub Saharan African” OR “sub-Saharan Africa” OR “sub-Saharan African”)

#### A.2.6 Cochrane Library

(Community Health Worker OR CHWs OR health volunteers OR CHV OR CHVs OR CHW OR Volunteer Health OR Village Health OR Lay Health OR Lay Counsel OR Health Extension OR Agentes Polivalentes OR Village Malaria OR Nutrition Volunteer OR Nutrition Worker OR Community Drug Distributor OR Community Reproductive Health OR Community Mobilizer OR Health Promoter OR Peer Counsellor OR Peer Navigat* OR Peer Counsel* OR Peer Educat* Peer Support* OR Peer-to-Peer OR Traditional Birth Attendant OR Health Extension Program OR Community Health Volunteers OR Village Health Worker OR Ward-Based Primary Health Care Outreach Team OR Health Care Outreach OR Paths to Primary Health Care OR Relais Communautaires OR Community health promoter OR Community health extension workers OR Village Health Teams OR Community health aide OR Community health promoters OR Linkage facilitators OR Health Extension worker OR Womens development army OR Community health agents OR Trained traditional birth attendants OR Volunteer health promoters OR Community health officer OR Community health nurse OR Community health assistant OR Community health service supervisors OR Trained traditional midwives OR Household health promoters OR Community directed distributors OR Agents Communautaires de Nutrition OR Agents Communautaires OR Community Nutrition Workers OR Community Workers OR Community Health Nurses OR Midwives OR Medical Assistants OR Environmental Health Assistants OR Health Surveillance Assistants OR Agentes Polivalentes Elementares OR Basic Multipurpose Health Workers OR Agents de Santé Communautaire OR Relais Volunteers OR Relay Volunteers OR Volunteer Village Health Workers OR Community Health Aides OR Community Health Assistants OR Christian Health Association of Nigeria OR Animatrice de Santé Maternelle OR Binôme OR Volunteer Cell Coordinators OR Maternal Health Moderator OR Binomial OR Traditional birth attendants OR Community drug distributors OR Ward-Based Primary Health Care Outreach Team OR Environmental Health Officers OR Village Executive Officers OR Community-Based Volunteers OR Village health worker OR Raedat Refiat village pioneer OR Agents de la santé communautaire OR Agents de santé communautaires OR Matrones bajenu gox OR Relais communautaires OR Relais Communautaire OR Agent Santé Communautaire OR Community Mobilizers OR Community counsellors OR Agent de Santé à Base Communautaire OR Community Health Agent OR Community relays OR Community health agents OR Village Health Workers OR Musharit among Sudanese refugees OR Cabillos OR Relais communautaires OR Relais de site OR Relais promotionnels OR Community based distributors OR Community national volunteers OR Home health promoters OR Community Volunteers OR Agents de Santé Communautaires OR Village Health Workers OR Leader Pastoral da Criança OR Volunteer Community-based Maternal and Newborn Care OR TBHIV Promoters OR Lifestyle Ambassadors OR Home-based Care Providers OR Health Education Assistants OR Community Health Home-based Care Volunteers OR Rural Health Motivators):ti,ab,kw AND (“adolescent” OR “youth” OR “young NEXT people” OR “teen” OR “teens” OR “teenager” OR “adolescents” OR “young NEXT women” OR “young NEXT men” OR “reproductive NEXT age” OR “15-24” OR “15-19”):ti,ab,kw AND (Africa OR Algeria OR Angola OR Benin OR Botswana OR Burkina Faso OR Burundi OR Cameroon OR Canary Islands OR Cape Verde OR Central African Republic OR Chad OR Comoros OR Congo OR Democratic Republic of Congo OR Djibouti OR Egypt OR Equatorial Guinea OR Eritrea OR Ethiopia OR Gabon OR Gambia OR Ghana OR Guinea OR Guinea Bissau OR Ivory Coast OR Cote dIvoire OR Jamahiriya OR Kenya OR Lesotho OR Liberia OR Libya OR Madagascar OR Malawi OR Mali OR Mauritania OR Mauritius OR Morocco OR Mozambique OR Namibia OR Niger OR Nigeria OR Principe OR Reunion OR Rwanda OR Sao Tome OR Senegal OR Seychelles OR Sierra Leone OR Somalia OR South Africa OR St Helena OR Sudan OR Swaziland OR Tanzania OR Togo OR Tunisia OR Uganda OR Western Sahara OR Zaire OR Zambia OR Zimbabwe OR Central Africa OR Central African OR West Africa OR West African OR Western Africa OR Western African OR East Africa OR East African OR Eastern Africa OR Eastern African OR North Africa OR North African OR Northern Africa OR Northern African OR South African OR Southern Africa OR Southern African OR sub Saharan Africa OR sub Saharan African OR subSaharan Africa OR subSaharan African):ti,ab,kw” with Cochrane Library publication date Between Jan 2014 and Nov 2024, in Trials (Word variations have been searched)

### Appendix A.3 Rationale for Bibliographic Database Search Terms

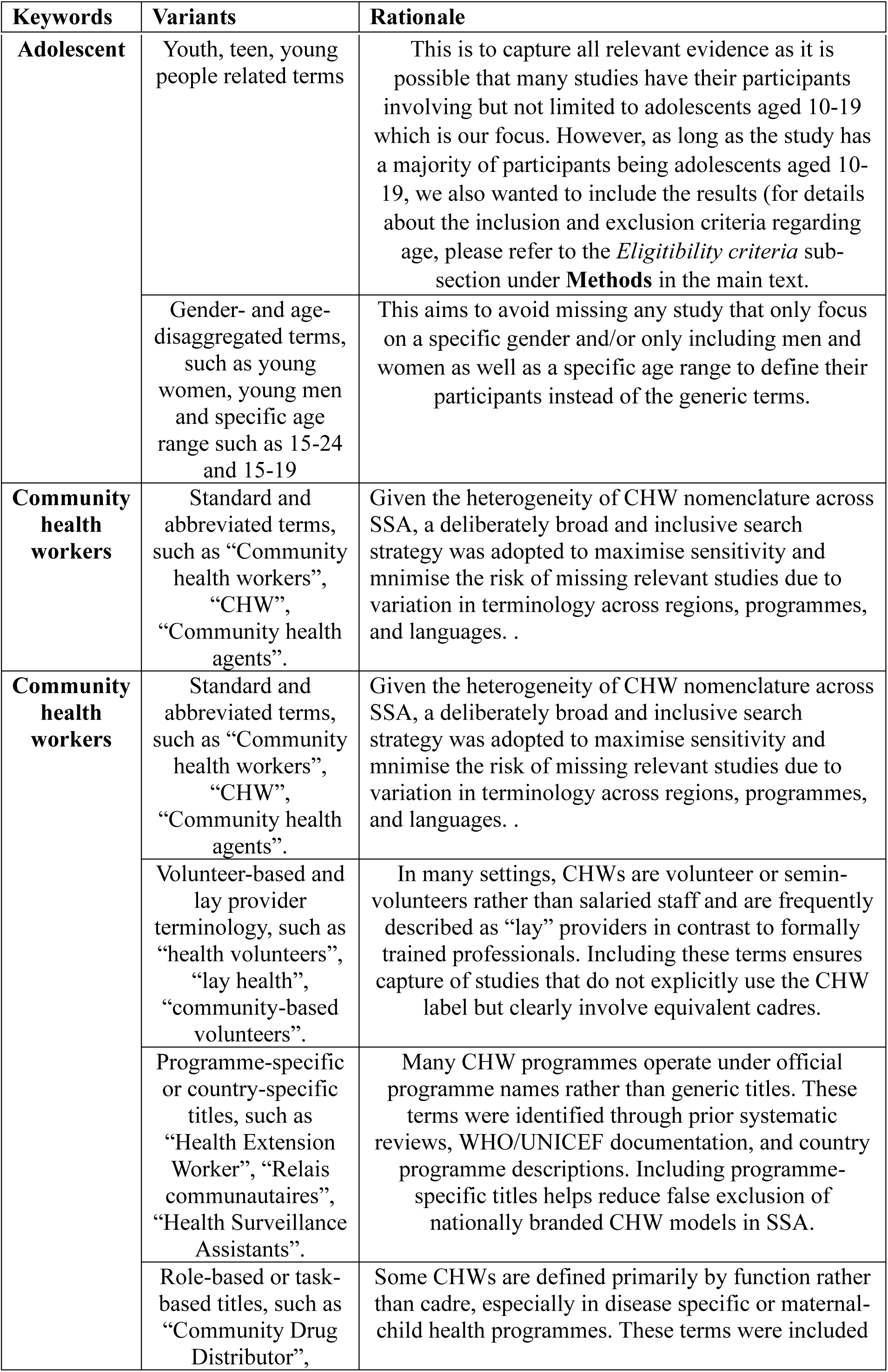
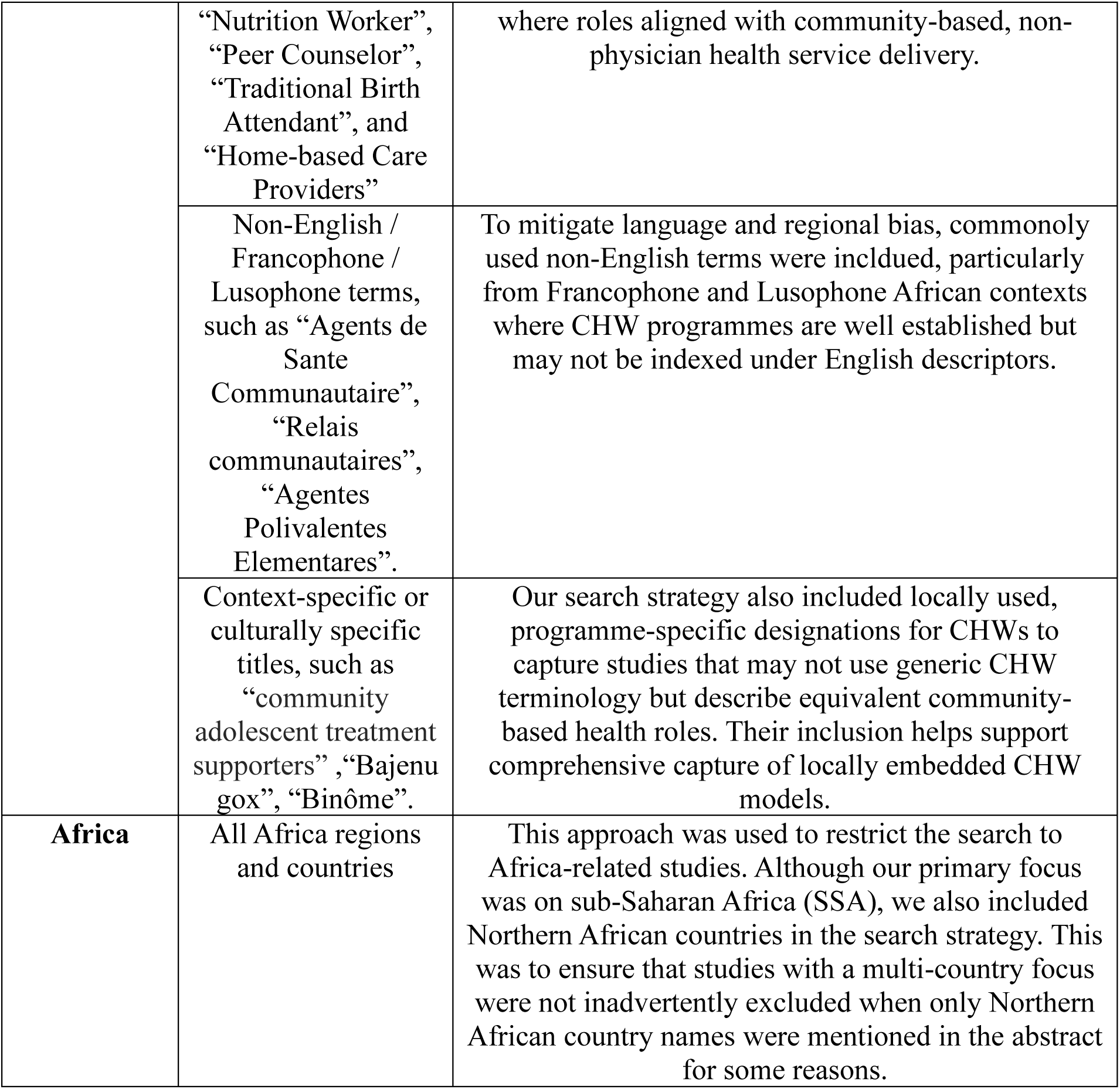

### Appendix A.4 Search Terms for Grey Literature Databases and Platforms

#### A.4.1 John Snow Inc., Population Council, FHI 360, PSI, Pathfinder International, PATH, Last Mile Health, OPM

The above sites were searched to identify documents related to community health workers and adolescent health in Sub-Saharan Africa. Two complementary approaches were used.

First, searches were conducted directly on the JSI website using combinations of keywords related to adolescents (and synonyms as used for electronic database search), community health workers, and Africa. Because the website does not support long strings of words linked by Boolean operators, simple keyword searches were applied (e.g., “community health worker adolescent Nigeria”). The search terms were developed based on the concepts of population, intervention, and geography. Geographic terms included two categories: (A) regional descriptors such as “Africa” and “Sub-Saharan Africa,” and (B) individual country names, as the use of regional descriptors alone returned limited results.

However, the internal JSI website search produced relatively few relevant results. Therefore, additional searches were conducted using Google site-search (e.g., site:jsi.com “community health worker” adolescent Kenya or site: jsi.com “community health worker” adolescent Africa) to identify relevant programme reports and technical documents hosted on the JSI website.

For each search, the first 50–100 results were screened for relevance.

##### A.4.1.1 Search Terms

- Community health worker: similar terms as in Appendix A.2
- Adolescent: similar terms as in Appendix A.2
- Region terms: Africa, sub-Saharan Africa
- Country terms: Angola, Benin, Botswana, Burkina Faso, Burundi, Cameroon, Central African Republic, Chad, Comoros, Congo, Democratic Republic of the Congo, Djibouti, Equatorial Guinea, Eritrea, Eswatini, Ethiopia, Gabon, Gambia, Ghana, Guinea, Guinea-Bissau, Kenya, Lesotho, Liberia, Madagascar, Malawi, Mali, Mauritania, Mozambique, Namibia, Niger, Nigeria, Rwanda, São Tomé and Príncipe, Senegal, Seychelles, Sierra Leone, Somalia, South Africa, South Sudan, Sudan, Tanzania, Togo, Uganda, Zambia, and Zimbabwe

##### A.4.1.2 Example Strings on John Snow Inc. Website

community health worker youth Africa

community health volunteer adolescent Africa

adolescent community health worker Africa

community health worker adolescent Kenya

community health worker young women Nigeria

community health worker young men Ethiopia

community health volunteer adolescent Uganda

community health worker adolescent Ghana

community health worker youth Tanzania

##### A.4.1.3 Example Search Strings on Google-site for John Snow Inc.-related Documents

site:jsi.com “community health worker” adolescent Africa

site:jsi.com “community health worker” youth Africa

site:jsi.com “community health worker” adolescent Kenya

site:jsi.com “community health worker” youth Nigeria

site:jsi.com “community health worker” adolescent Ethiopia

site:jsi.com “community health worker” adolescent Uganda

site:jsi.com “community health worker” adolescent Ghana

site:jsi.com “community health worker” adolescent Tanzania

#### Appendix A.4.2 WHO, UNICEF, World Bank, IFPRI

Grey literature searches were conducted on the above websites to identify reports, guidelines, technical briefs, and policy documents related to community health workers and adolescent health in Africa. Because websites including the WHO search engine has limited functionality compared with bibliographic databases such as Ovid, complex Boolean search strings were not used. Instead, shorter combinations of keywords and phrases were applied. Where possible, Boolean operators such as AND were used to combine key concepts. If the results internally returned limited hits, then it was supplemented with google site searching

##### A.4.2.1 Search Terms

- Community health worker: similar terms as in Appendix A.2
- Adolescent: similar terms as in Appendix A.2
- Region terms: Africa, sub-Saharan Africa
- Country terms: Angola, Benin, Botswana, Burkina Faso, Burundi, Cameroon, Central African Republic, Chad, Comoros, Congo, Democratic Republic of the Congo, Djibouti, Equatorial Guinea, Eritrea, Eswatini, Ethiopia, Gabon, Gambia, Ghana, Guinea, Guinea-Bissau, Kenya, Lesotho, Liberia, Madagascar, Malawi, Mali, Mauritania, Mozambique, Namibia, Niger, Nigeria, Rwanda, São Tomé and Príncipe, Senegal, Seychelles, Sierra Leone, Somalia, South Africa, South Sudan, Sudan, Tanzania, Togo, Uganda, Zambia, and Zimbabwe

##### A.4.2.2 Example Strings on WHO Website

“community health worker” AND adolescent AND Africa

“community health worker” AND youth AND Africa

“community health volunteer” AND adolescent AND Africa

“community health worker” AND adolescent AND Kenya

“community health worker” AND youth AND Nigeria

“community health worker” AND adolescent AND Ethiopia

“community health volunteer” AND adolescent AND Uganda

##### A.4.2.3 Example Strings on Google-site for WHO-related Documents

site:who.int “community health worker” adolescent Africa

site:who.int “community health worker” youth Africa

site:who.int “community health worker” adolescent Kenya

site:who.int “community health worker” youth Nigeria

site:who.int “community health worker” adolescent Ethiopia

site:who.int “community health worker” adolescent Uganda

site:who.int “community health worker” adolescent Ghana

site:who.int “community health worker” adolescent Tanzania

#### A.4.3 Overton, Eldis, BASE, CIAO, Google Scholar

Additional grey literature searches were conducted in the above online policy and research repositories that support Boolean searching. Searches combined terms relating to the population (adolescents and related terms), the intervention (community health workers and related cadres), and the geographic setting (Africa). Boolean operators (AND, OR) were used to structure the search strings, with synonyms combined using OR and the three concept groups combined using AND. Phrase searching using quotation marks was also applied where appropriate (e.g., “community health worker”). The geographic component included both regional descriptors (e.g., “Africa” and “Sub-Saharan Africa”) and selected country names to ensure retrieval of documents referring to specific national programmes. Search results were screened based on titles and available summaries to identify potentially relevant reports, working papers, and policy documents related to our topic. Relevant documents were downloaded and assessed for eligibility, and all searches were recorded to ensure transparency and reproducibility.

##### A.4.3.1 Search Strings

(adolescent OR adolescents OR youth OR “young people” OR “young person” OR teen OR teens OR teenager OR “young women” OR “young men”)

AND

(“community health worker” OR “community health workers” OR CHW OR CHWs OR “community health volunteer” OR “community health volunteers” OR “village health worker” OR “village health workers” OR “lay health worker” OR “health extension worker” OR “community health assistant” OR “community health aide” OR “community health agent” OR “community mobilizer” OR “peer counsellor” OR “traditional birth attendant”)

AND

(Africa OR “sub-Saharan Africa” OR “sub Saharan Africa” OR Kenya OR Nigeria OR Ethiopia OR Uganda OR Ghana OR Tanzania OR Malawi OR Zambia OR Rwanda OR Senegal OR Mozambique OR South Africa)

### Appendix A.5 Rationale for the Grey Literature Search Strategy

Grey literature searches were conducted across relevant organisational websites and online repositories to identify programme reports, technical briefs, and evaluation documents relating to community health workers and adolescent health in Africa. A similar search strategy to that used in the electronic database searches was applied. However, because some websites did not support advanced Boolean searching, searches were conducted using combinations of keywords and phrases rather than complex Boolean search strings.

Websites such as John Snow Inc., Population Council, FHI 360, Population Services International, Pathfinder International, PATH, Last Mile Health, Oxford Policy Management, and the Transfer Project have limited search functionality and therefore required simple keyword searches. Keywords identified as effective during the electronic database searches, including terms relating to adolescents, youth, community health workers, and Africa, were used during the website searches. Geographic terms included both regional descriptors (e.g., “Africa” and “Sub-Saharan Africa”) and individual African country names to maximise retrieval of relevant documents.

For websites with limited internal search capabilities, additional searches were conducted using Google site-search (e.g., site:who.int “community health worker” adolescent Africa) to identify relevant documents from organisations such as the World Health Organisation, UNICEF, the World Bank, and the International Food Policy Research Institute. Where websites returned a limited number of results, all available publications within relevant sections were screened manually. Titles and summaries were reviewed to identify potentially relevant reports, and documents of potential relevance to the review question were downloaded and assessed for eligibility. All searches and results were recorded to ensure transparency and reproducibility of the search process.

### Appendix B. Definition for Key Concepts

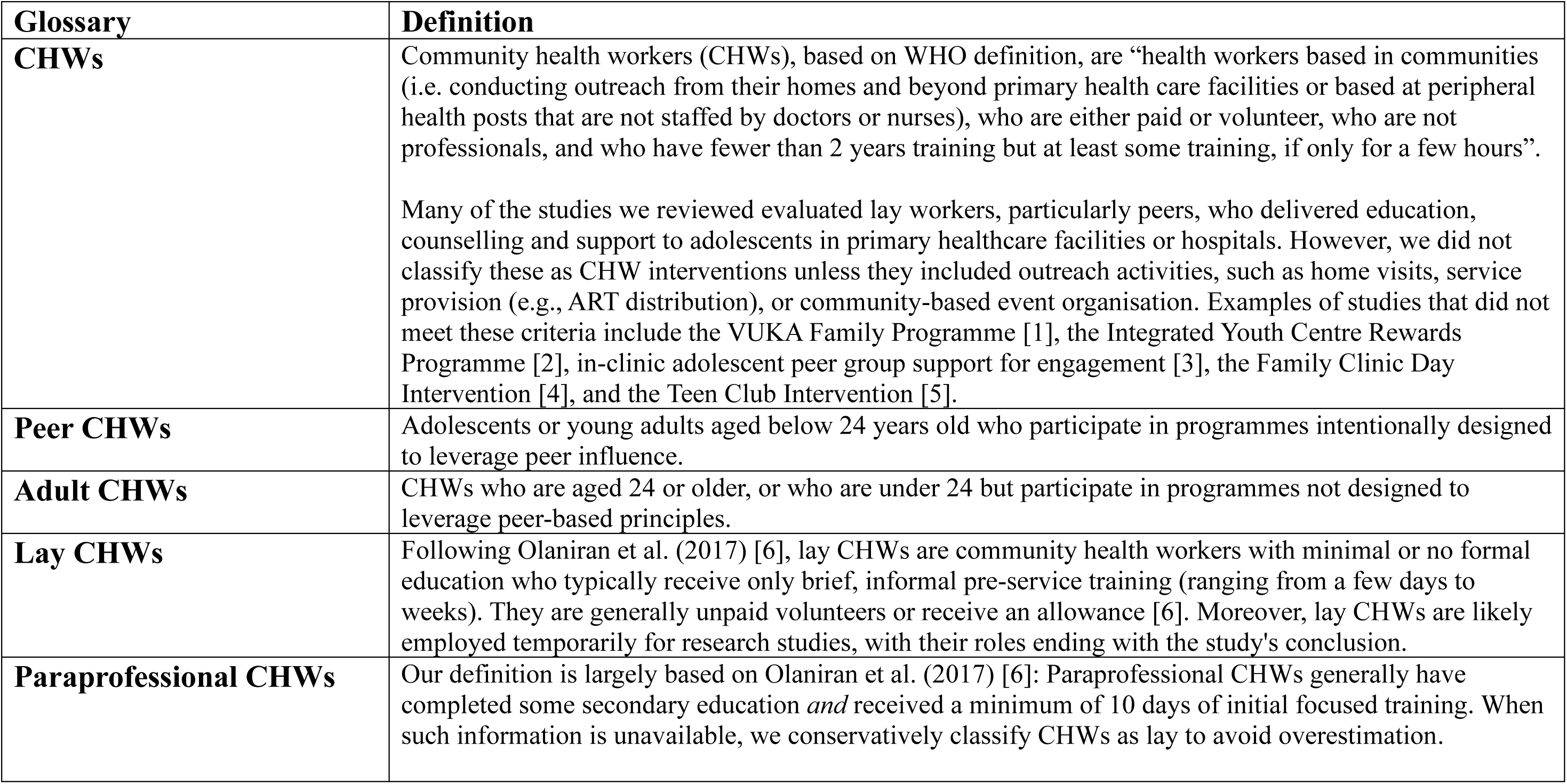
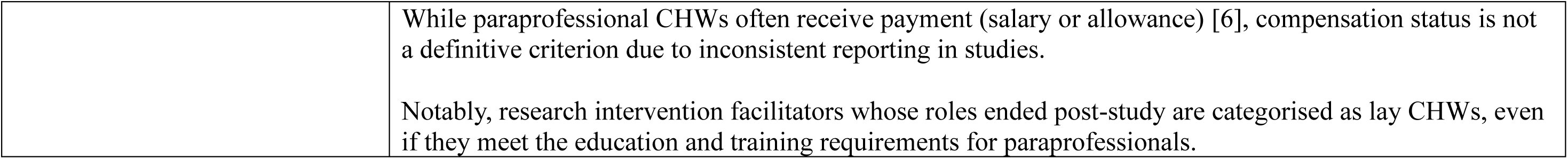

Appendix B. Definition for Key Concepts.
| Glossary | Definition |
| --- | --- |
| **CHWs** | Community health workers (CHWs), based on WHO definition, are “health workers based in communities (i.e. conducting outreach from their homes and beyond primary health care facilities or based at peripheral health posts that are not staffed by doctors or nurses), who are either paid or volunteer, who are not professionals, and who have fewer than 2 years training but at least some training, if only for a few hours”.<br>Many of the studies we reviewed evaluated lay workers, particularly peers, who delivered education, counselling and support to adolescents in primary healthcare facilities or hospitals. However, we did not classify these as CHW interventions unless they included outreach activities, such as home visits, service provision (e.g., ART distribution), or community-based event organisation. Examples of studies that did not meet these criteria include the VUKA Family Programme [1], the Integrated Youth Centre Rewards Programme [2], in-clinic adolescent peer group support for engagement [3], the Family Clinic Day Intervention [4], and the Teen Club Intervention [5]. |
| **Peer CHWs** | Adolescents or young adults aged below 24 years old who participate in programmes intentionally designed to leverage peer influence. |
| **Adult CHWs** | CHWs who are aged 24 or older, or who are under 24 but participate in programmes not designed to leverage peer-based principles. |
| **Lay CHWs** | Following Olaniran et al. (2017) [6], lay CHWs are community health workers with minimal or no formal education who typically receive only brief, informal pre-service training (ranging from a few days to weeks). They are generally unpaid volunteers or receive an allowance [6]. Moreover, lay CHWs are likely employed temporarily for research studies, with their roles ending with the study's conclusion. |
| **Paraprofessional CHWs** | Our definition is largely based on Olaniran et al. (2017) [6]: Paraprofessional CHWs generally have completed some secondary education *and* received a minimum of 10 days of initial focused training. When such information is unavailable, we conservatively classify CHWs as lay to avoid overestimation. |
|  | While paraprofessional CHWs often receive payment (salary or allowance) [6], compensation status is not a definitive criterion due to inconsistent reporting in studies.<br>Notably, research intervention facilitators whose roles ended post-study are categorised as lay CHWs, even if they meet the education and training requirements for paraprofessionals. |

### Appendix C. Data Items

Information sought from included studies included: (i) Study identification details: first author, title, year of publication, peer reviewed publication (Yes/No); (ii) Study characteristics: region and country of study, urban or rural settings, aim of the study, study design, data collection timeline, and sample size; (iii) Participant information: target population and the age, gender and socioeconomic status of the population, (iv) Programme details: name of the programme, description of the intervention, age, gender and educational profile of CHW, training of CHW, whether it was delivered by a governmental or non-governmental entity and the control condition; and (v) Outcome-related information: the health knowledge, perception, or well-being outcome measured, the indicator(s) used to measure outcomes, description of this indicator(s); (vi) Effect related analysis for quantitative studies: direction, size and significance of the effect, 95% confidence interval; (vii) Gender-related analysis: any gender-disaggregated results.

### Appendix D. PRISMA flowchart diagram

**Figure D.1.**
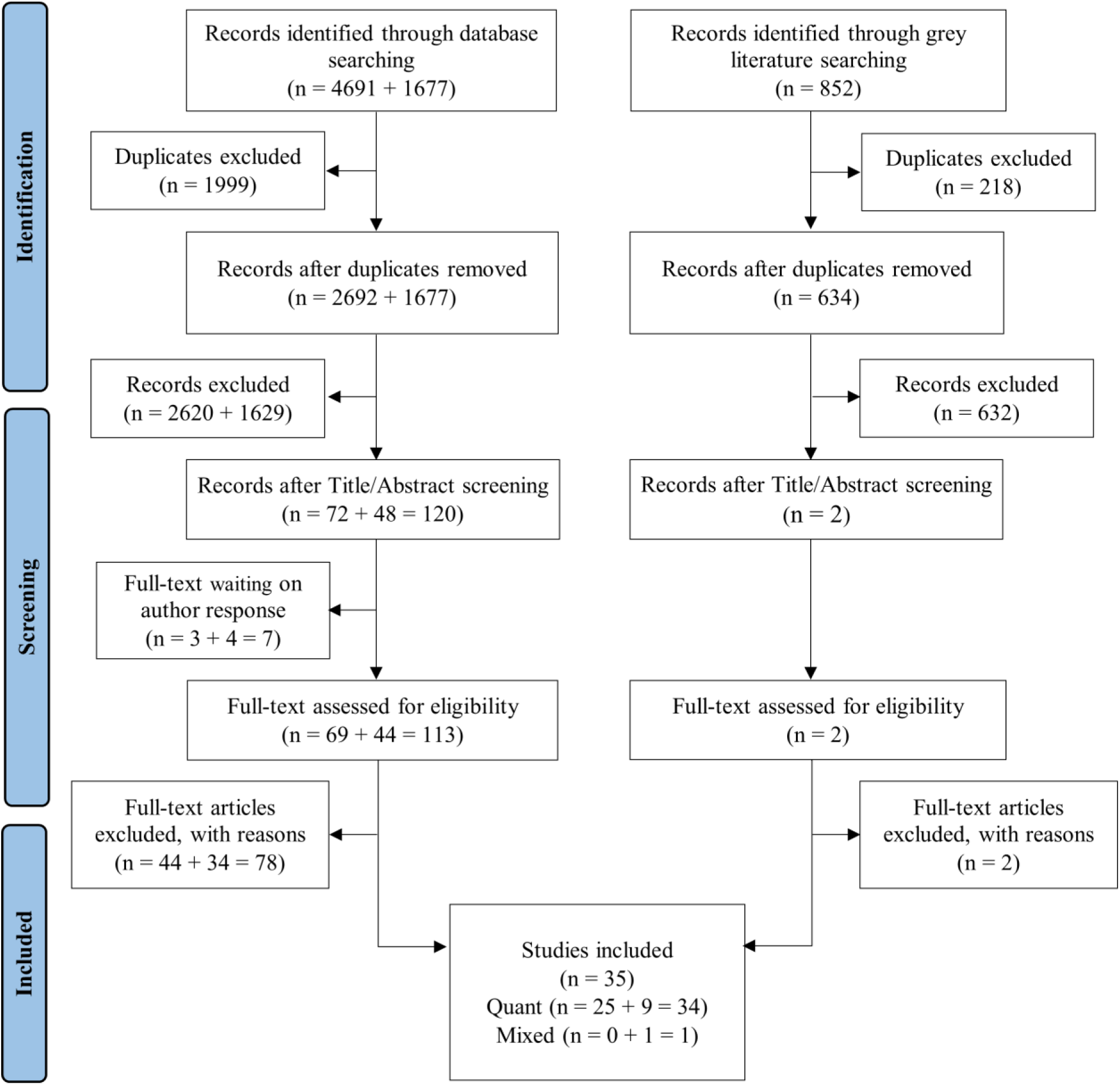
PRISMA flowchart diagram. **Note:** When two numbers were connected with a “+” sign, the first referred to records from the electronic database search, and the second to records identified through backward searching of reference lists from all electronic database papers and two highly relevant systematic reviews [7,8].

### Appendix E. Summary of Excluded Papers and Reasons at Full-Text Screening Stage

Appendix E. Summary of Excluded Papers and Reasons at Full-Text Screening Stage.
| No. | Author, Year | Title | Reason for exclusion |
| --- | --- | --- | --- |
| 1 | Adeomi et al., 2014 | Evaluation of the Effectiveness of Peer Education in Improving HIV Knowledge, Attitude, and Sexual Behaviours among In-School Adolescents in Osun State, Nigeria | Quantitative results are insufficient to draw robust conclusions about the effectiveness of interventions. |
| 2 | Adam et al., 2015 | Relationship between implementing interpersonal communication and mass education campaigns in emergency settings and use of reproductive healthcare services: evidence from Darfur, Sudan | Median age is between 20-30, which is outside our age criteria, and there are no appropriate age-disaggregated results. |
| 3 | Ahmed et al., 2023 | A Systematic Review of Peer Support Interventions for Adolescents Living with HIV in Sub-Saharan Africa | This is a review paper. |
| 4 | Maclean et al., 2023 | Addressing the Treatment Gap in Adolescent Mental Healthcare in Botswana Using the Safe Haven Intervention: A Pilot Study | Full-text is not available from the author's reply. |
| 5 | Mangombe et al., 2020 | Does peer education go beyond giving reproductive health information? Cohort study in Bulawayo and Mount Darwin, Zimbabwe | There is a lack of a control group. Specifically, this is a cross-sectional study without a control group. |
| 6 | Morgan et al., 2020 | Effects of a Community-Based Program on Voluntary Modern Contraceptive Uptake Among Young First-Time Parents in Cross River State, Nigeria | Mean age is 20.6, which is outside our age criteria, and there are no age-disaggregated results. |
| 7 | Nakalega, 2023 | Feasibility and acceptability of peer-delivered HIV self-testing and PrEP for young women in Kampala, Uganda | Median age is 20, which is outside our age criteria, and there are no age-disaggregated results. |
| 8 | Shaame et al., 2023 | Outreach service-an effective approach to reducing teenage pregnancy in katavi region-experience from Amref Health Africa in Tanzania | Full-text is not available. |
| 9 | Shah et al., 2019 | A Pilot Study of "Peer Navigators" to Promote Uptake of HIV Testing, Care and Treatment Among Street-Connected Children and Youth in Eldoret, Kenya | There is a lack of a control group. |
| 10 | Speizer et al., 2003 | An Evaluation of the "Entre Nous Jeunes" Peer-Educator Program for Adolescents in Cameroon | Year of publication is before 2014. |
| 11 | Undie et al., 2012 | Expanding access to comprehensive reproductive health and HIV information and services for married adolescent girls in Nyanza Province | Year of publication is before 2014. |
| 12 | Usman et al., 2019 | Achieving the third 90: Keeping adolescents living with HIV virally suppressed in rural Nigeria in the era of test and treat | Full-text is not available. |
| **13** | Vu et al., 2017 | Increasing Uptake of HIV, Sexually Transmitted Infection, and Family Planning Services, and Reducing HIV-Related Risk Behaviors Among Youth Living With HIV in Uganda | Median age is 20, which is outside our age criteria, and there are no age-disaggregated results available for key outcomes. |
| **14** | Indravudh et al., 2019 | Community-led delivery of HIV self-testing targeting adolescents and men in rural Malawi: A cluster-randomised trial | This is a protocol for a paper that was included. |
| **15** | Puffer et al., 2020 | Family Functioning and Mental Health Changes Following a Family Therapy Intervention in Kenya: a Pilot Trial | Sample size (n=10) in the quantitative section is limited for regression. |
| **16** | Harrison et al., 2023 | Better Together: acceptability, feasibility and preliminary impact of chronic illness peer support groups for South African adolescents and young adults | The intervention is clinic-based peer support group, which doesn't fit our criteria on CHW-led intervention. |
| **17** | Mathew et al., 2019 | Effects of Peer Education Intervention for Hepatitis B on Level of Knowledge and Beliefs of School Adolescents in Jos, Plateau State Nigeria | Lack of clear information that makes it impossible to extract a reliable effect size |
| **18** | Kebede et al., 2020 | School-based social and behavior change communication (SBCC) advances community exposure to malaria messages, acceptance, and preventive practices in Ethiopia: A pre-posttest study | Although the intervention targeted school students/adolescents, the outcomes were measured for households or respondents aged over 37, which doesn't fit our criterion of focusing on adolescent outcomes. |
| **19** | Fikree et al., 2018 | The effect of peer education in dispelling myths and misconceptions about long-acting reversible contraception among ethiopian youth | The study setting is YFS units in 20 health centres and there are very limited outreach activities. Furthermore, there is no age information about the participants. The data source is peer educators' assessment reports. |
| **20** | Bhana et al., 2014 | The VUKA Family Program: Piloting a family-based psychosocial intervention to promote health and mental health among HIV-infected early adolescents in South Africa | The intervention primarily took place at two clinical sites, though the exact locations varied based on participant and provider feedback. Additionally, there is no evidence of facilitators engaging in outreach activities such as home visits, community-based service provision, or organising community group meetings. Lastly, the intervention facilitators include both lay counsellors and one master's-level psychologist. |
| **21** | Hensen et al., 2022 | Access to menstrual hygiene products through incentivised, community-based, peer-led sexual and reproductive health services before and during the COVID-19 pandemic: findings from the Yathu Yathu trial | The study took place within intervention communities only. In other words, it doesn't have a control group. |
| **22** | Phiri et al., 2022 | Community-based, peer-led SRH services increase coverage in AYP (Yathu Yathu Trial) | This paper has the same author list but with different orders as a previously included paper. In the analysis of RCT results in this paper, no formal tests were performed, and they only reported the proportional changes. |
| **23** | Hensen et al., 2021 | Uptake of HIV testing services through novel community-based sexual and reproductive health services: An analysis of the pilot implementation phase of the Yathu Yathu intervention for adolescents and young people aged 15-24 in Lusaka, Zambia | This is a pilot phase paper with only a poster. Its formal version is included in the study. |
| **24** | Black et al., 2014 | Improving HIV testing amongst adolescents through an integrated Youth Centre rewards program: Insights from South Africa | The main intervention evaluated is clinic-based and mainly facilitated by clinical nurse practitioner rather than youth counsellor. |
| **25** | Rosenberg et al., 2018 | Comparing Youth Friendly Health Services to the Standard of Care through "Girl Power-Malawi": A Quasi-Experimental Cohort Study | It is not possible to unpack how much of the intervention is due to clinic adjustments vs. behavioural intervention. |
| **26** | Shanaube et al., 2021 | HIV Care Cascade Among Adolescents in a "Test and Treat" Community-Based Intervention: HPTN 071 (PopART) for Youth Study | No statistical test was performed. |
| **27** | Barker et al., 2019 | In-Clinic Adolescent Peer Group Support for Engagement in Sub-Saharan Africa: A Feasibility and Acceptability Trial | The intervention is based in a clinic likely staffed by professionals, and facilitators were four bachelor-level health-care professionals (who were already working in the clinic as counsellors or research assistants). |
| **28** | Comfort et al., 2019 | Do home pregnancy tests bring women to community health workers for antenatal care counselling? A randomized controlled trial in Madagascar | Neither is this study adolescent-specific, nor does it have age-disaggregated results. |
| **29** | Robb, 2020 | Support Group to Reduce Diabetes Distress in Adolescents with Type 1 Diabetes | Though not explicitly mentioned, it is likely based in the USA according to the author affiliation |
| **30** | Denison et al., 2020 | Project YES! Youth Engaging for Success: A randomized controlled trial assessing the impact of a clinic-based peer mentoring program on viral suppression, adherence and internalized stigma among HIV-positive youth (15-24 years) in Ndola, Zambia | In the trial registration (link, please see below), it's quite clear that the study is in a clinic setting.<br>https://clinicaltrials.gov/study/NCT04115813?id=NCT04115813&rank=1#study-plan |
| **31** | Chinoda et al., 2020 | Effectiveness of a peer-led adolescent mental health intervention on HIV virological suppression and mental health in Zimbabwe: protocol of a cluster-randomised trial | This is a protocol for a paper that we've already included. |
| **32** | Buchwald et al., 2016 | Bed net use among school-aged children after a universal bed net campaign in Malawi | In the intervention paper CHWs only distributed a fraction of bed nets:<br>https://journals.plos.org/plosone/article?id=10.1371/journal.pone.0134061#sec002 |
| **33** | Elul et al., 2017 | A combination intervention strategy to improve linkage to and retention in HIV care following diagnosis in Mozambique: A cluster randomized study | The paper does not effectively involve CHWs (wrong intervention) and focuses on adults (wrong population). |
| **34** | McNairy et al., 2017 | Effectiveness of a combination strategy for linkage and retention in adult HIV care in Swaziland: The Link4Health cluster randomized trial | The paper does not effectively involve CHWs (wrong intervention) and focuses on adults (wrong population). |
| **35** | Galárraga et al., 2019 | Group-based economic incentives to improve adherence to antiretroviral therapy among youth living with HIV: safety and preliminary efficacy from a pilot trial | Clinic-based peer groups in a hospital. |
| **36** | Graves et al., 2018 | Impact of a Family Clinic Day intervention on paediatric and adolescent appointment adherence and retention in antiretroviral therapy: A cluster randomized controlled trial in Uganda | The intervention is based in quite well-equipped health centres (Level III, level IV, and hospitals). |
| **37** | MacKenzie et al., 2017 | Greater retention in care among adolescents on antiretroviral treatment accessing “Teen Club” an adolescent-centred differentiated care model compared with standard of care: a nested case-control study at a tertiary referral hospital in Malawi | The teen club is based in hospital. |
| **38** | Tozan et al., 2021 | The efficacy and cost-effectiveness of a family-based economic | All activities were held at participating health centres and primarily facilitated by lay |
|  |  | empowerment intervention (Suubi + Adherence) on suppression of HIV viral loads among adolescents living with HIV: results from a Cluster Randomized Controlled Trial in southern Uganda | counsellors and programme staff |
| 40 | Chory et al., 2022 | A Pilot Study of a Mobile Intervention to Support Mental Health and Adherence Among Adolescents Living with HIV in Western Kenya | This study explores a mobile intervention, which doesn't fit our criteria. |
| 41 | Kasimonje et al., 2021 | Group counselling for adherence support among young people failing first-line antiretroviral therapy in Zimbabwe | Peer counsellors only provided support alongside professional psychologists. |
| 42 | Ssewamala et al., 2020 | The long-term effects of a family based economic empowerment intervention (Suubi+Adherence) on suppression of HIV viral loads among adolescents living with HIV in southern Uganda: Findings from 5-year cluster randomized trial | The intervention is based in quite well-equipped health centres (Level III, level IV, and hospitals). |
| 43 | Cavazos-Rehg et al., 2021 | The Impact of a Family-Based Economic Intervention on the Mental Health of HIV-Infected Adolescents in Uganda: Results From Suubi + Adherence | The intervention is based in quite well-equipped health centres (Level III, level IV, and hospitals). |
| 44 | Agbaje, 2015 | Mitigating the impact of HIV/AIDS among in and out of school youth through peer education using family life HIV education in federal capital territory abuja Nigeria | Fail to contact authors for more information to help with our inclusion/exclusion decision. |
| 45 | Ivanova et al., 2019 | Evaluation of the elimika pilot project: Improving art adherence among hiv positive youth using an ehealth intervention in Mombasa, Kenya | Fail to contact authors for more information to help with our inclusion/exclusion decision. |
| 46 | Agbede et al., 2021 | An outcome of educational intervention on the menstrual hygiene practices among school girls in Ogun State, Nigeria: a quasi-experimental study | Fail to contact authors for more information to help with our inclusion/exclusion decision. |
| 47 | Ibitoye et al., 2021 | The Impact of Education on Knowledge Attitude and Practice of Breast Self-Examination Among Adolescents Girls at the Fiwasaye Girls Grammar School Akure, Nigeria | Fail to contact authors for more information to help with our inclusion/exclusion decision. |
| 48 | Nnabueze et al., 2021 | Impact of a Peer Public Health Education Programme on Students' Knowledge of HIV//AIDS and Attitude Towards People Living with HIV/AIDS in Abia State, South East, Nigeria | Fail to contact authors for more information to help with our inclusion/exclusion decision.<sup>49</sup> |
| 49 | Scheel et al., 2018 | The impact of a peer support group for children with rheumatic heart disease in Uganda | Fail to contact authors for more information to help with our inclusion/exclusion decision. |
| **50** | Chowdhary et al., 2022 | Building sustainable and scalable peer-based programming: promising approaches from TESFA in Ethiopia | Fail to contact authors for more information to help with our inclusion/exclusion decision. |
| **51** | Henning et al., 2020 | Adolescent Mothers' Experiences With Community Health Assistants in Rural Zambia | Though it met our inclusion criteria, it is a qualitative study, which we excluded for this review. This will be included in another review that focuses on qualitative insights. |
| **52** | Atujuna et al., 2021 | Khuluma: Using Participatory, Peer-Led and Digital Methods to Deliver Psychosocial Support to Young People Living With HIV in South Africa | Though it met our inclusion criteria, it is a qualitative study, which we excluded for this review. This will be included in another review that focuses on qualitative insights. |
| **53** | Broström et al., 2021 | "Seeing a Brighter Future"—Experiences of Adolescents with Common Mental Disorders Receiving the Problem-Solving Therapy "Youth Friendship Bench" in Zimbabwe | Though it met our inclusion criteria, it is a qualitative study, which we excluded for this review. This will be included in another review that focuses on qualitative insights. |
| **54** | Kereta et al., 2021 | Role and contribution of peer educators in youth-friendly health services in Ethiopia: evidence from programmatic experience with a peer education intervention | Qualitative study with no insights from adolescents. |
| **55** | Kereta et al., 2021 | Role and contribution of peer educators in youth-friendly health services in Ethiopia: evidence from programmatic experience with a peer education intervention | Duplicate with no. 50 above. |
| **56** | Garriott et al., 2024 | Youth Perspectives of Working with Near Peer Lay Counsellors: The Safe Haven Pilot | Though it met our inclusion criteria, it is a qualitative study, which we excluded for this review. This will be included in another review that focuses on qualitative insights. |
| **57** | Bedingar et al., 2024 | Exploring the barriers and facilitators to HIV information and health services among youth in N'Djamena, Chad: a qualitative descriptive study | Though it met our inclusion criteria, it is a qualitative study, which we excluded for this review. This will be included in another review that focuses on qualitative insights. |
| **58** | Burke et al. 2024 | Documenting the Provision of Emergency Contraceptive Pills Through Youth-Serving Delivery Channels: Exploratory Mixed Methods Research on Malawi's Emergency Contraception Strategy | Though it met our inclusion criteria, it is a qualitative study, which we excluded for this review. This will be included in another review that focuses on qualitative insights. |
| **59** | Kawuma et al., 2024 | Gendered risks: access to and utilization of sexual and reproductive health services among young migrants in Southwestern Uganda: the role of the 'lending a hand' intervention | Though it met our inclusion criteria, it is a qualitative study, which we excluded for this review. This will be included in another review that focuses on qualitative insights. |
| **60** | Kitetele et al., 2022 | "The Peer Educator Is the Game-Changer of My Life": Perceptions of Adolescents Living with HIV in DR Congo on Involving Peer Educators in the Process of HIV Disclosure | Qualitative study that was based in health facility rather than community. |
| **61** | Miyingo et al., 2023 | HIV Treatment and Care of Adolescents: Perspectives of Adolescents on Community-Based Models in Northern Uganda | Qualitative study with not enough data on actual experiences of CHWs. |
| **62** | Okonji et al., 2022 | Psychosocial Support Programme Improves Adherence and Health Systems Experiences for Adolescents on Antiretroviral Therapy in Mpumalanga Province, South Africa | Qualitative study based in health facilities. |
| **63** | Tuhebwe et al., 2024 | Improving HPV vaccine acceptance through peer-to-peer education among adolescent girls in the urban poor settings of Kisenyi, Kampala, Uganda | Qualitative study with results mainly from peer educators. |
| **64** | Roberts et al., 2023 | How a menu of adherence support strategies facilitated high adherence to HIV prevention products among adolescent girls and young women in sub-Saharan Africa: a mixed methods analysis | Although this study was labelled as mixed-methods, it lacked robust quantitative data and was therefore treated as purely qualitative. As our review focuses on quantitative evidence, it was excluded from the analysis. |
| **65** | Wogrin et al., 2019 | Development and piloting of a novel, peer-led bereavement intervention for young people living with HIV in Zimbabwe | Qualitative study with not enough data on experiences of CHWs. |
| **66** | N'goran et al., 2024 | Lessons learned from peer-led support groups for HIV-positive adolescents using a storytelling model in Côte d'Ivoire | Qualitative study from grey literature with intervention conducted within health facilities. |
| **67** | Population Council, 2016 | Using peers to improve sexual and reproductive health and rights of young people living with HIV in Uganda: Findings from a Link Up evaluation | The median age of the participants (ranging from 15 to 24) is 20 years old, which exceeds our requirement. Moreover, the qualitative section of the study did not have specified adolescent perspective. |
| **68** | Murray et al., 2014 | Counselor and client perspectives of Trauma-focused Cognitive Behavioral | Qualitative study with not enough data from adolescents |
|  |  | Therapy for children in Zambia: a qualitative study | (also unclear if it includes adolescents as they mention children, but not the age range) |
| **69** | Tinago et al., 2021 | Development and assessment of feasibility of a community-based peer support intervention to mitigate social isolation and stigma of adolescent motherhood in Harare, Zimbabwe | Qualitative study with no actual experiences mentioned |
| **70** | Broström et al., 2021 | “Seeing a Brighter Future” – Experiences of Adolescents with Common Mental Disorders Receiving the ProblemSolving Therapy “Youth Friendship Bench” in Zimbabwe | Duplicate with a qualitative study that met our inclusion criteria. |
| **71** | Tembo et al., 2022 | Integration of a menstrual health intervention in a community-based sexual and reproductive health service for young people in ZimbabweL a qualitative acceptability study | Though it met our inclusion criteria, it is a qualitative study, which we excluded for this review. This will be included in another review that focuses on qualitative insights. |
| **72** | Stangl et al., 2021 | Feasibility, acceptability and preliminary efficacy of Tikambisane (‘Let’s Talk to Each Other’): A pilot support group intervention for adolescent girls living with HIV in Zambia. | Though it met our inclusion criteria, it is a qualitative study, which we excluded for this review. This will be included in another review that focuses on qualitative insights. |
| **73** | Ajuwon et al. (2015) | Effects of peer education on the knowledge and use of HIV counselling and testing services among young persons in Ibadan, Nigeria | Uncontrolled pre-post design, which does not meet our methodology requirement |
| **74** | Akuiyibo et al. (2021) | Impact of peer education on sexual health knowledge among adolescents and young persons in two North Western states of Nigeria | Uncontrolled pre-post design, which does not meet our methodology requirement |
| **75** | Ezelote et al. (2024) | Effect of peer health education intervention on HIV/AIDS knowledge amongst in-school adolescents in secondary schools in Imo State, Nigeria | Uncontrolled pre-post design, which does not meet our methodology requirement |
| **76** | Sadoh et al. (2018) | Effect of peer education on knowledge of human papilloma virus and cervical cancer among female adolescent students in Benin city, Nigeria | Uncontrolled pre-post with repeated cross-sectional design, which does not meet our methodology requirement |
| **77** | Sadoh et al. (2021) | Improving knowledge about breast cancer and breast self-examination in female Nigerian adolescents using peer education: a pre-post interventional study | Uncontrolled pre-post with repeated cross-sectional design, which does not meet our methodology requirement |
| **78** | Olaoye et al. (2024) | Evaluation of a school-based health education program on hepatitis B virus infection prevention practice in rural South-Western, Nigeria | Uncontrolled pre-post design, which does not meet our methodology requirement |
| **79** | Chizoba et al. (2020) | Effects of Peer and Provider-Based Education Interventions on HIV/AIDS Knowledge and Behaviour-Risk among in-School Adolescents in Ebonyi State, Nigeria | Uncontrolled pre-post design, which does not meet our methodology requirement |
| **80** | Boeke et al. (2018) | Results from a proactive follow-up intervention to improve linkage and retention among people living with HIV in Uganda: a pre-/post- study | Uncontrolled pre-post with repeated cross-sectional design, which does not meet our methodology requirement |
| **81** | Ali et al. (2015) | The Effect of AIDS Peer Health Education on Knowledge, Attitudes, and Practices of Secondary School Students in Khartoum, Sudan | Uncontrolled pre-post design, which does not meet our methodology requirement |
| **82** | Alhassan et al. (2024) | Four antenatal care visits by four months of pregnancy and four vital tests for pregnant mothers: impact of a community-facility health systems strengthening intervention in Migori County, Kenya | Uncontrolled pre-post with repeated cross-sectional design, which does not meet our methodology requirement |
| **83** | Weidert et al. (2017) | Community health workers as social marketers of injectable contraceptives: A case study from Ethiopia | Uncontrolled pre-post design, which does not meet our methodology requirement |
| **84** | O'Donnell et al. (2014) | Treating Unresolved Grief and Posttraumatic Stress Symptoms in Orphaned Children in Tanzania: Group-Based Trauma-Focused Cognitive-Behavioral Therapy | Uncontrolled pre-post design, which does not meet our methodology requirement |
| **85** | Timol et al. (2016) | Addressing adolescents' risk and protective factors related to risky behaviours: Findings from a school-based peer-education evaluation in the Western Cape | Uncontrolled pre-post design, which does not meet our methodology requirement |
| **86** | Benoni et al. (2024) | Assessing the impact of a community-based psychodrama intervention on mental health promotion of adolescents and young adults in Mozambique: A mixed-methods study | Uncontrolled pre-post design, which does not meet our methodology requirement |
| **87** | Shanaube et al. (2017) | Community intervention improves knowledge of HIV status of adolescents in Zambia: findings from HPTN 071-PopART for youth study | Uncontrolled pre-post design, which does not meet our methodology requirement |

Note: This table lists all studies excluded at the full-text screening stage (n = 87). These comprised 44 studies identified through the first-round electronic database search, 34 through backward searching, and 2 from grey literature. In addition, 7 studies were excluded due to a lack of author response to requests for additional information required to determine eligibility.

### Appendix F. Description of Included Studies

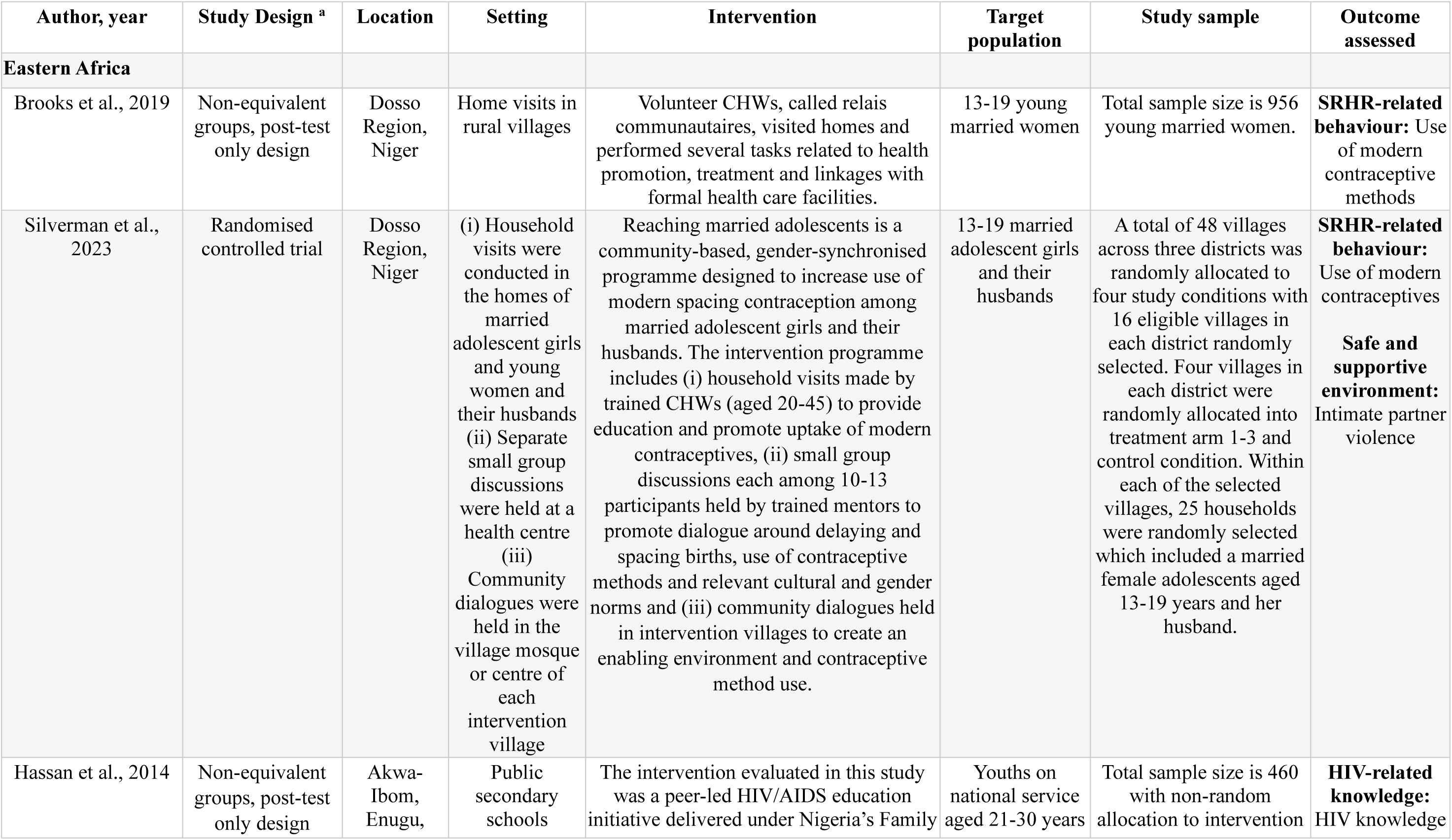
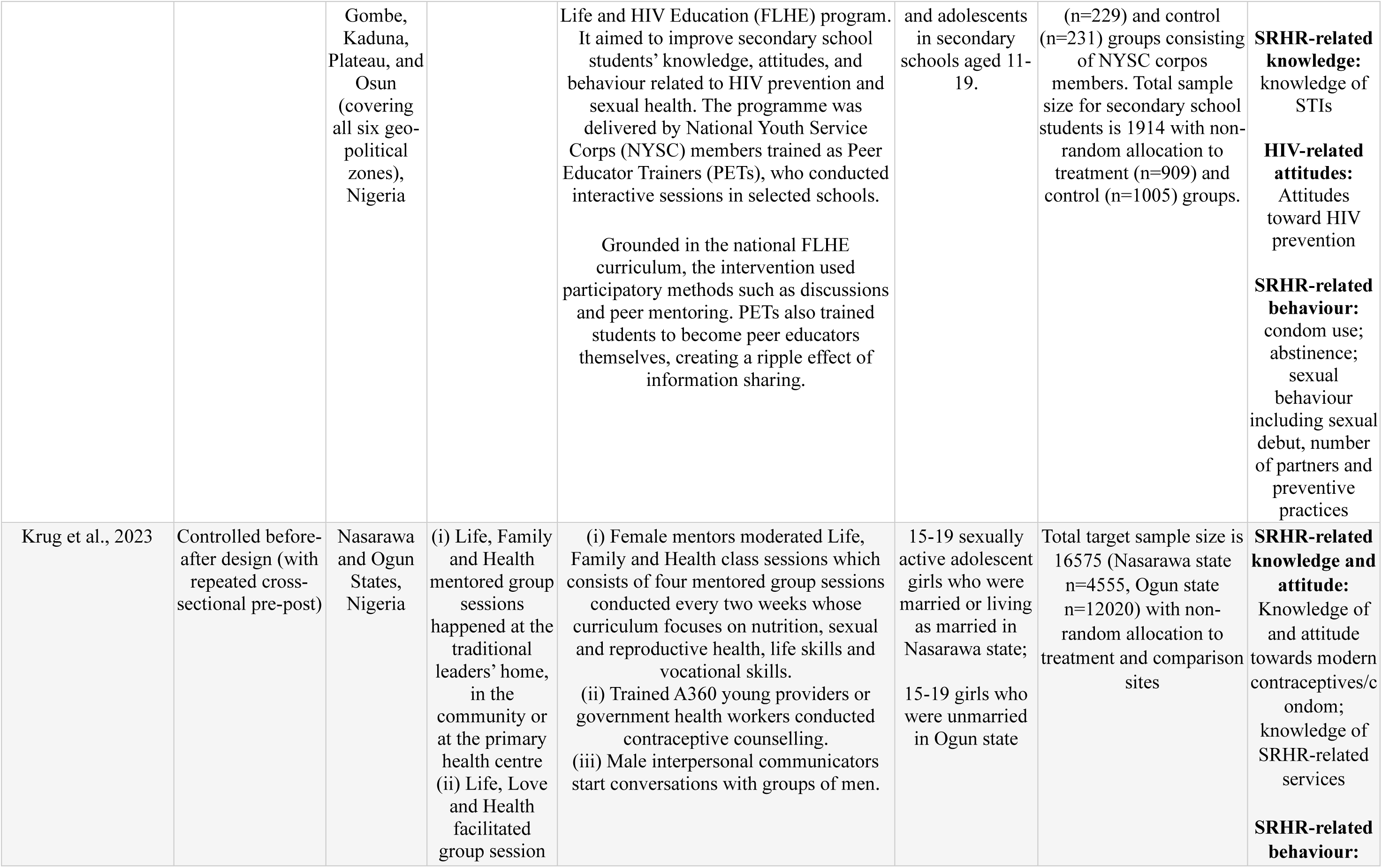
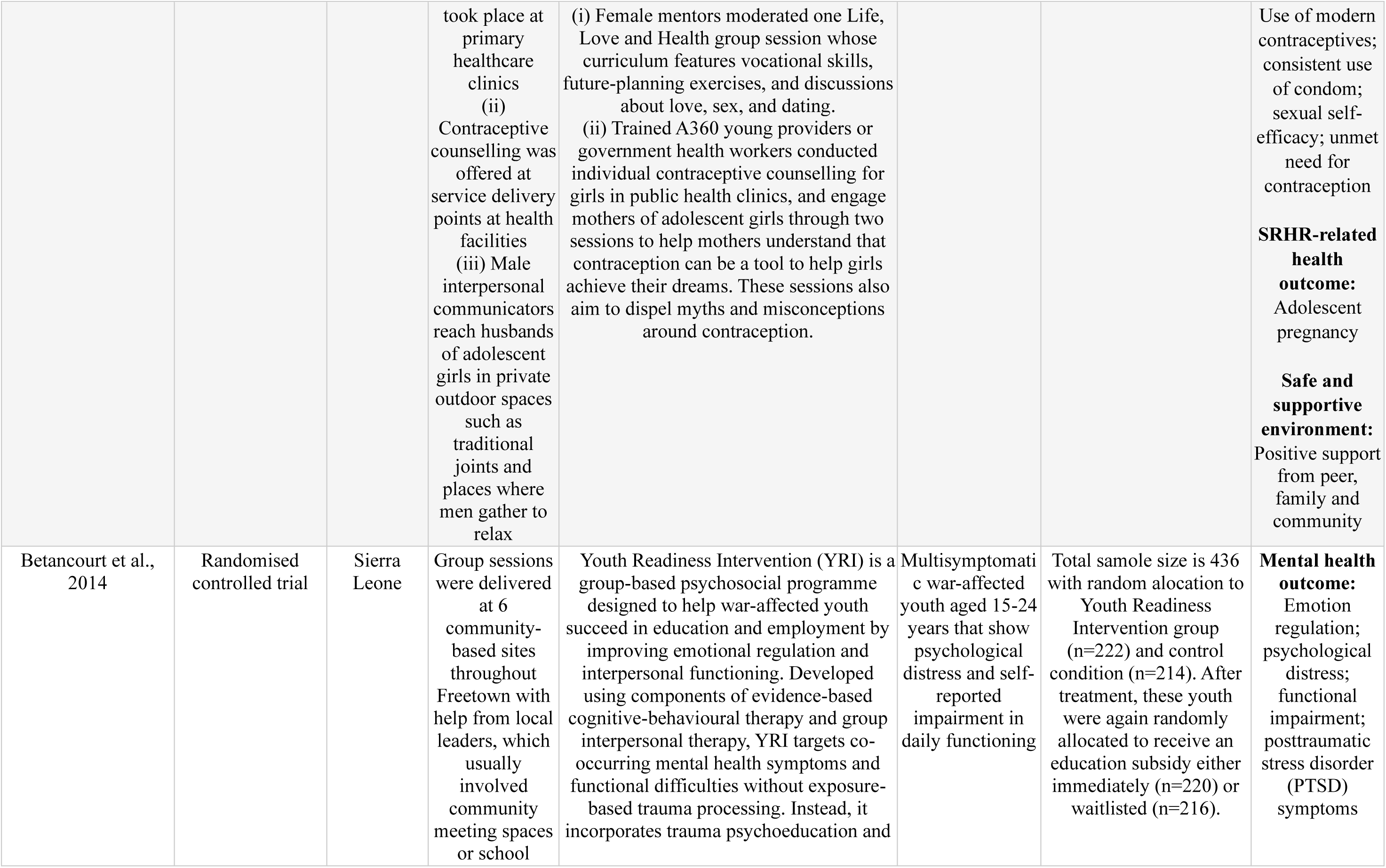
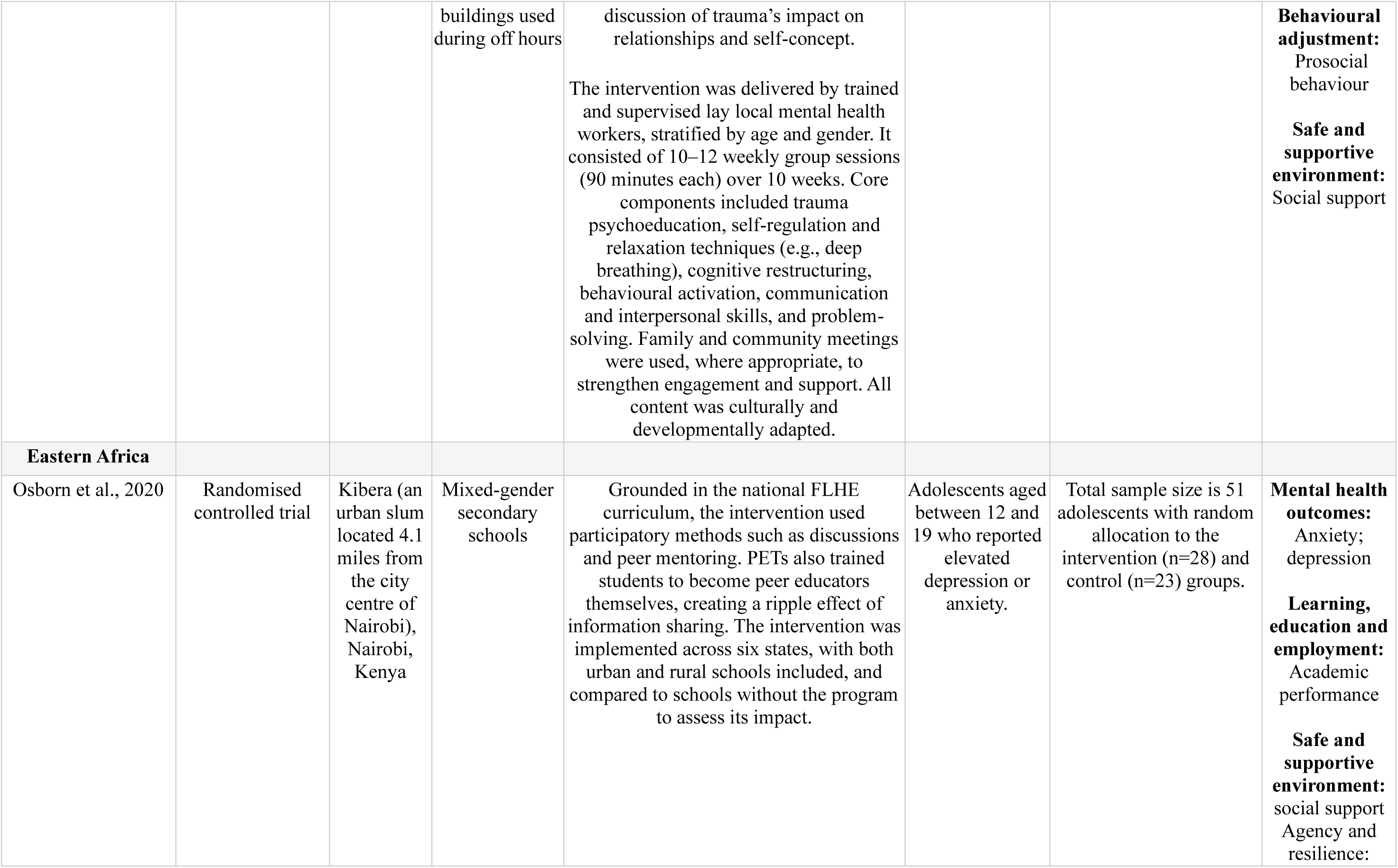
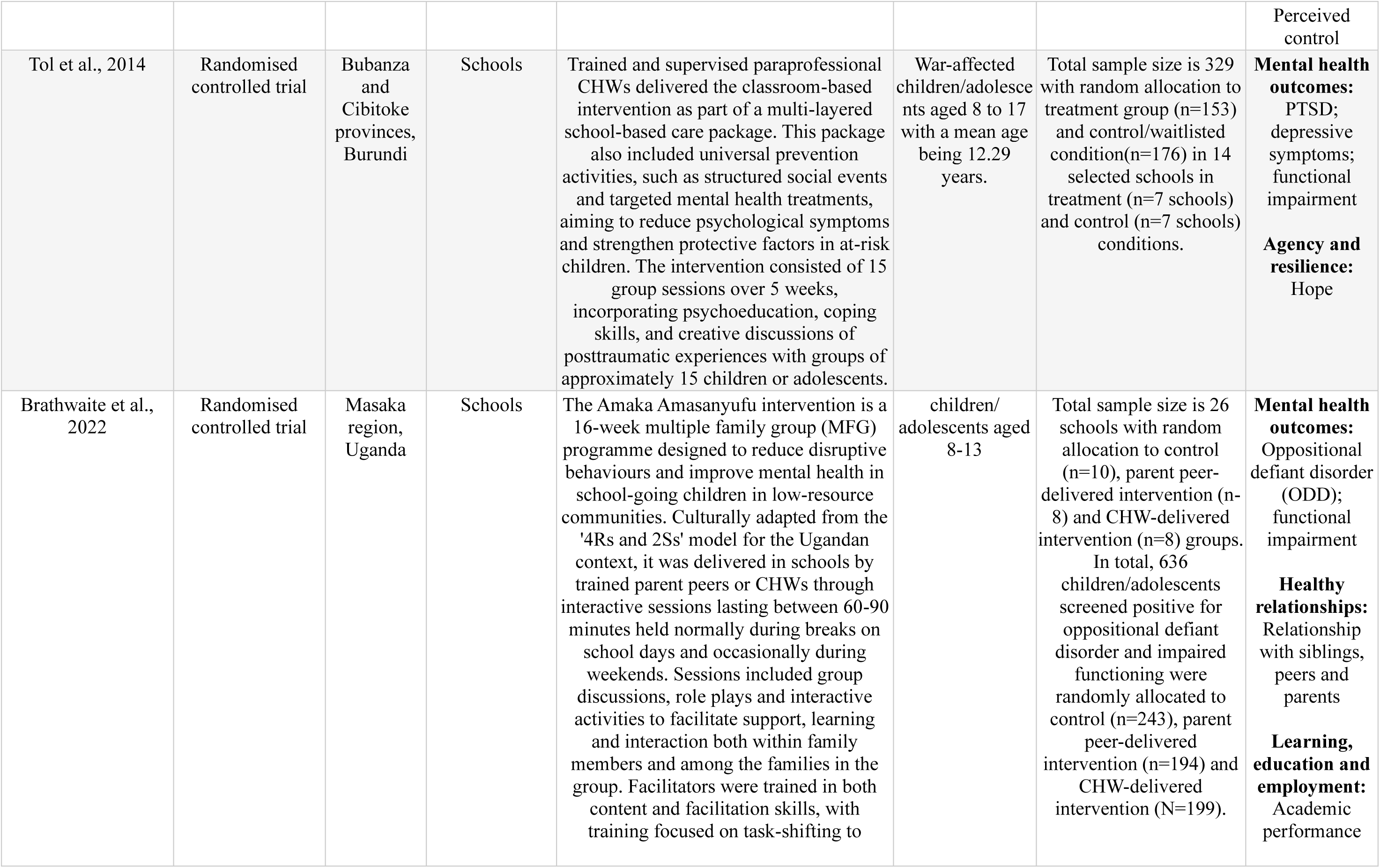
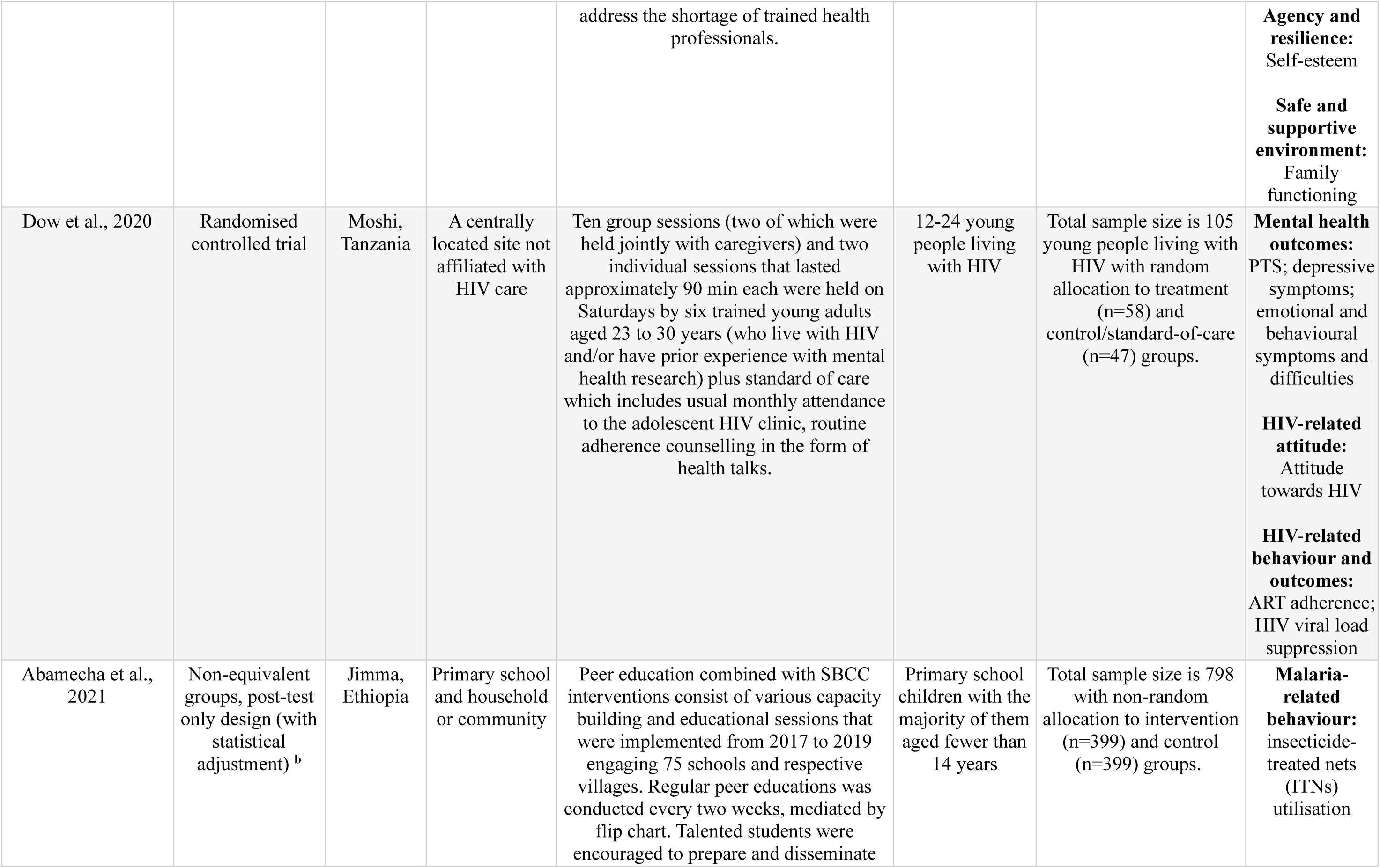
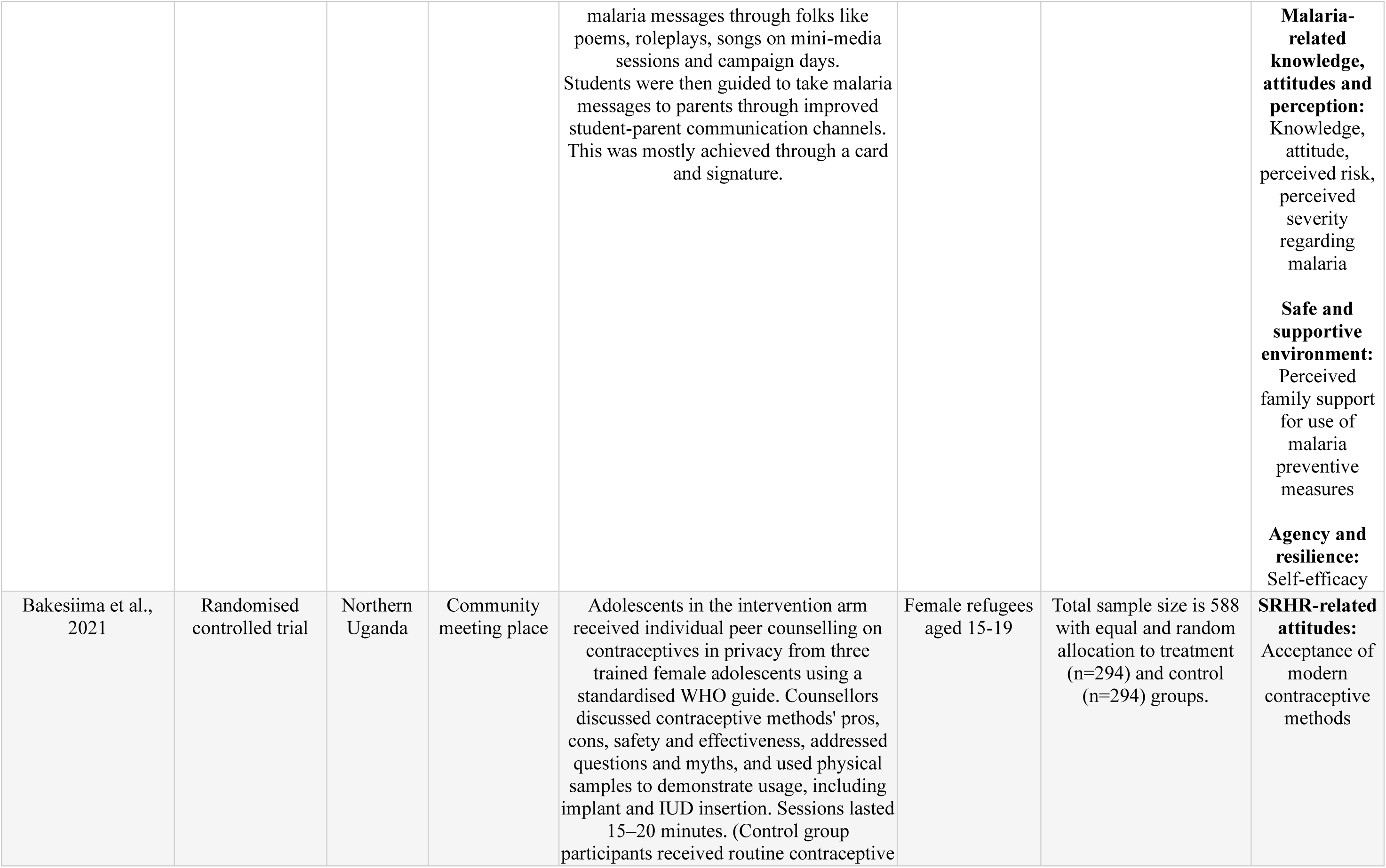
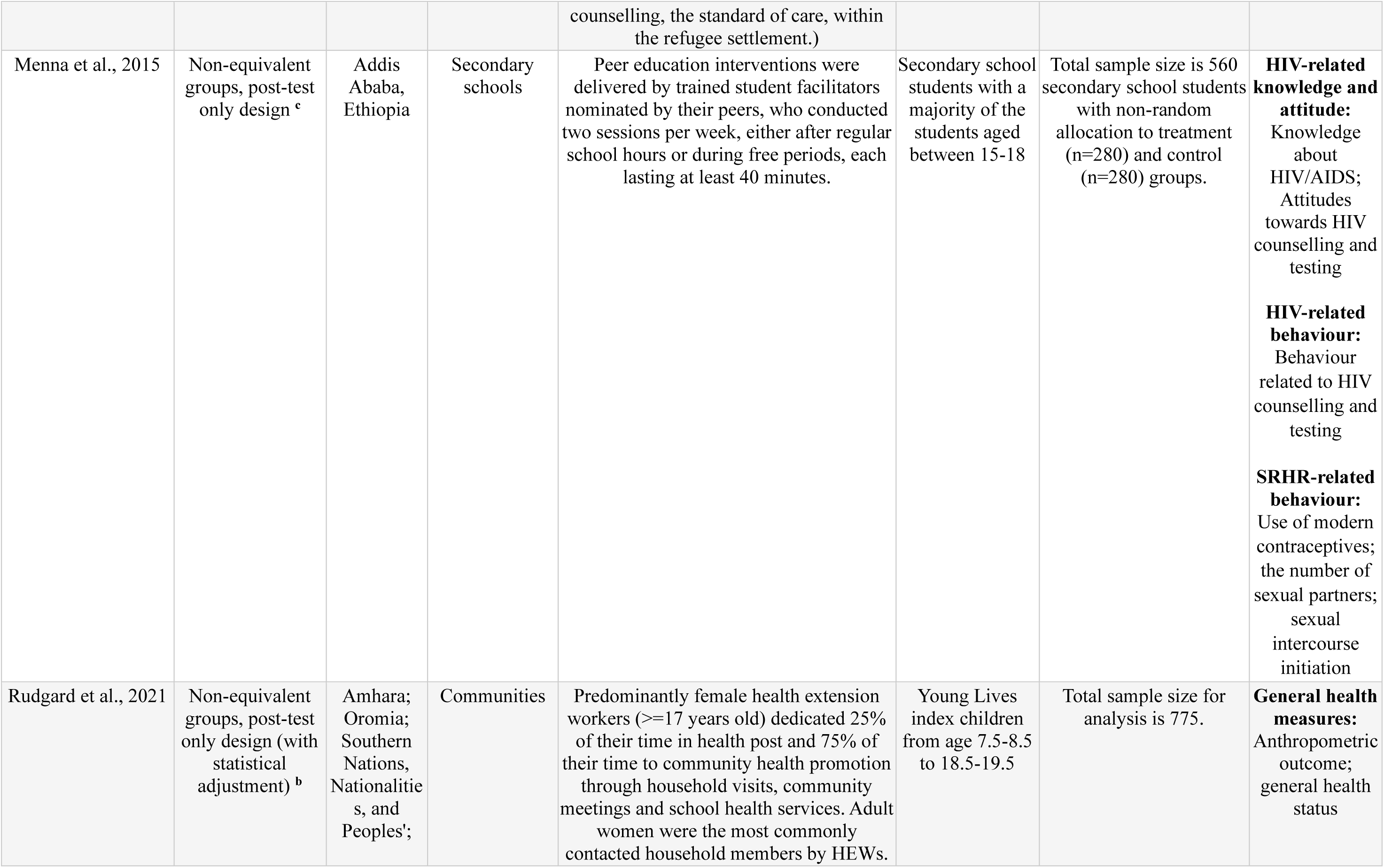
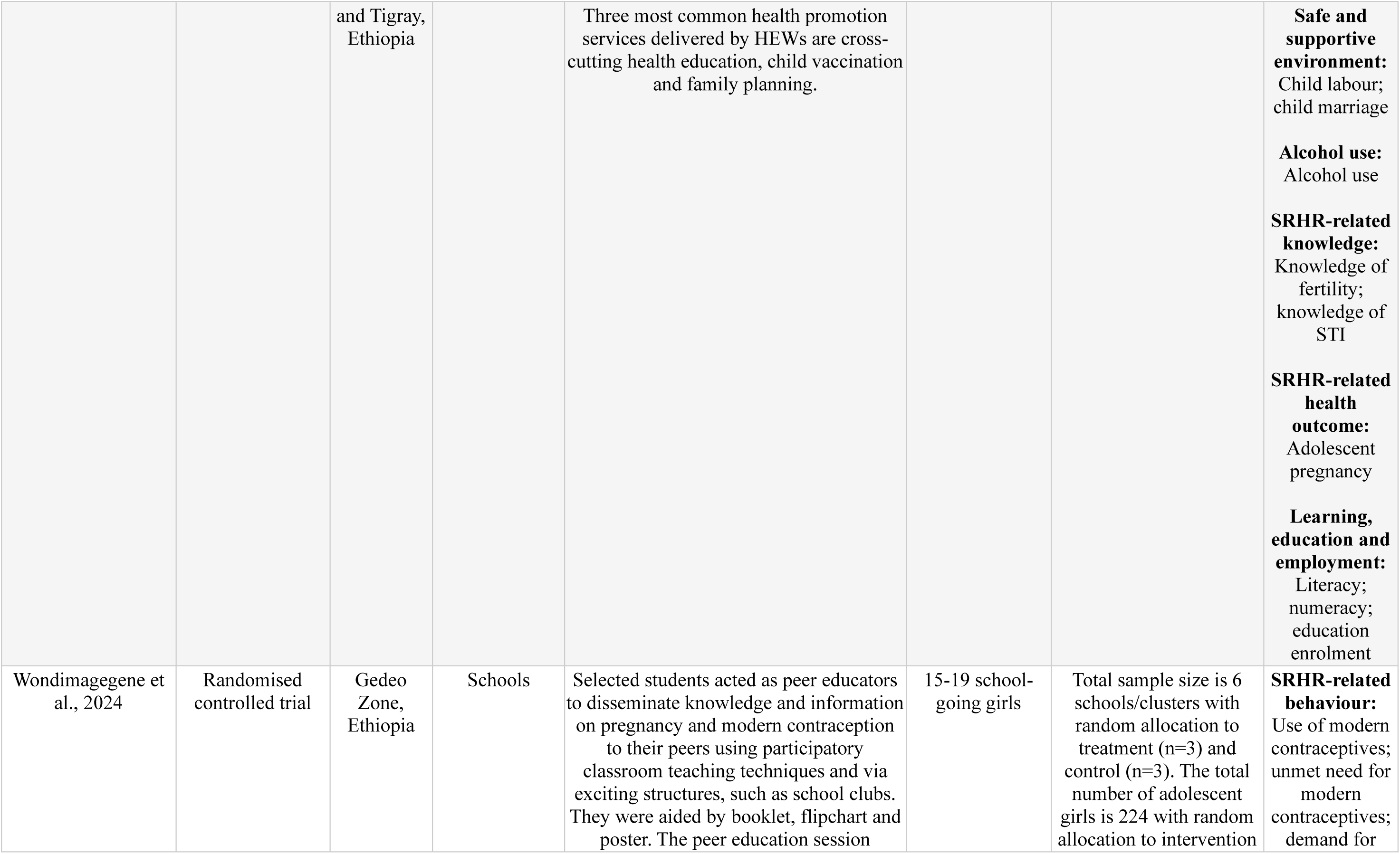
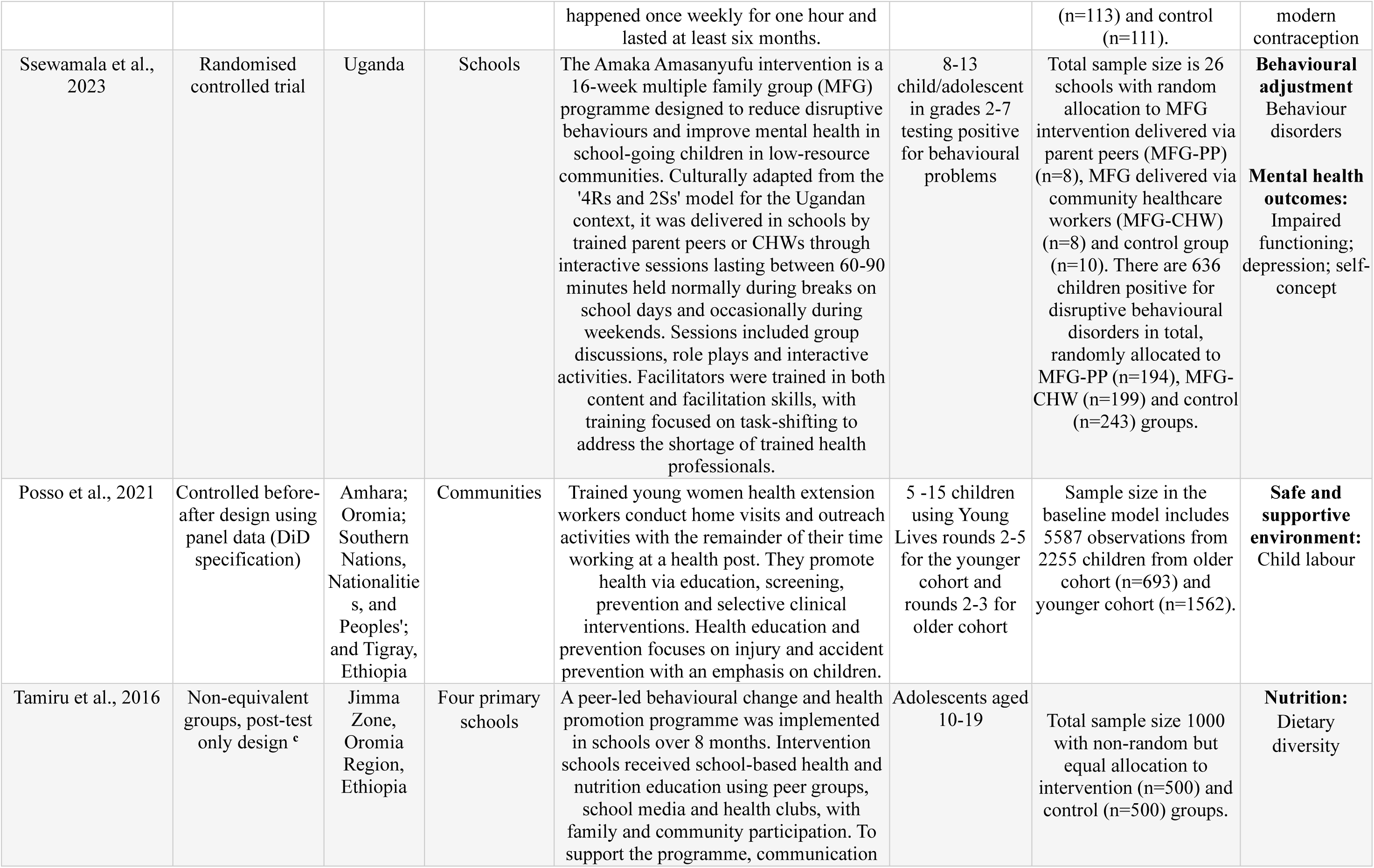
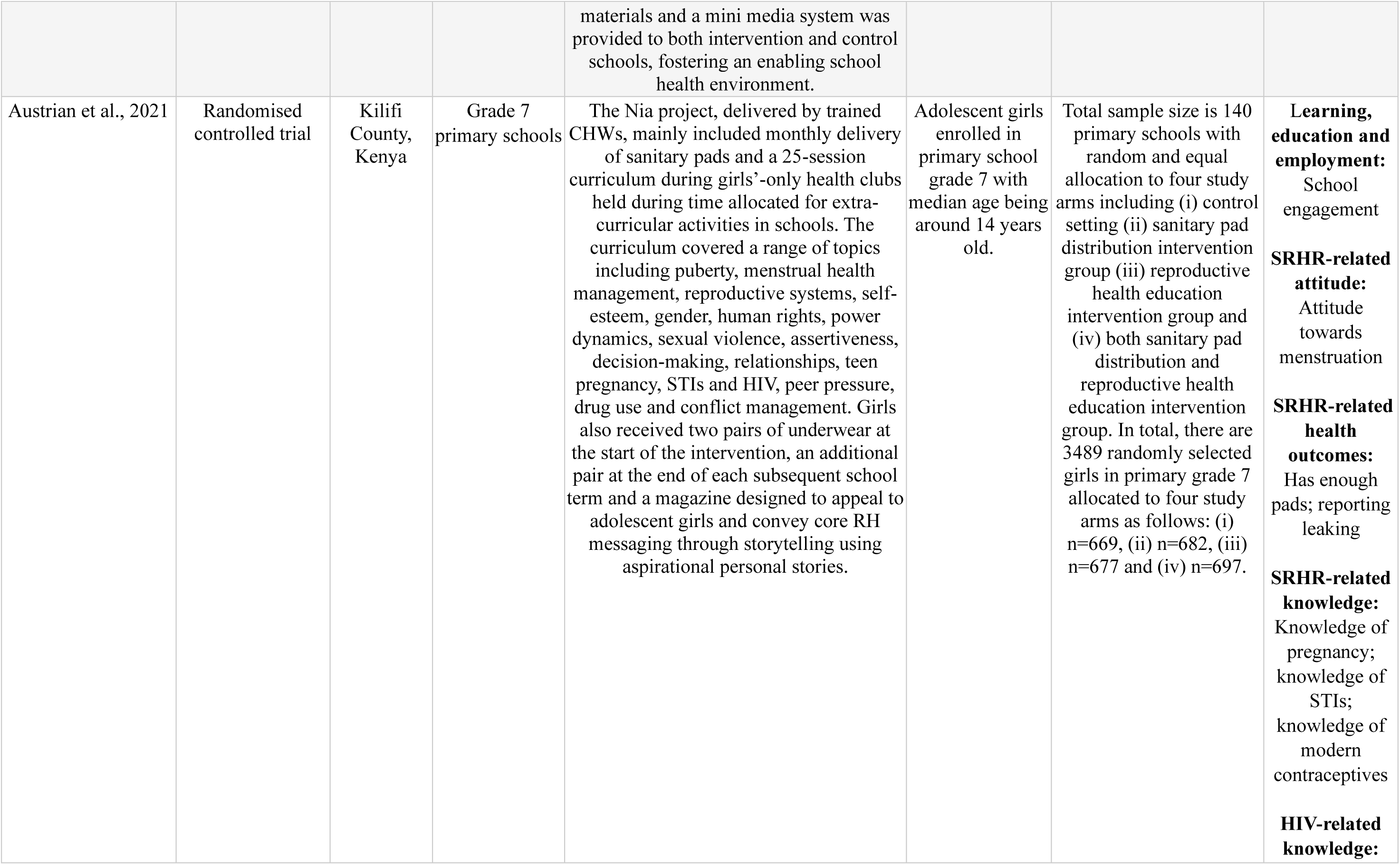
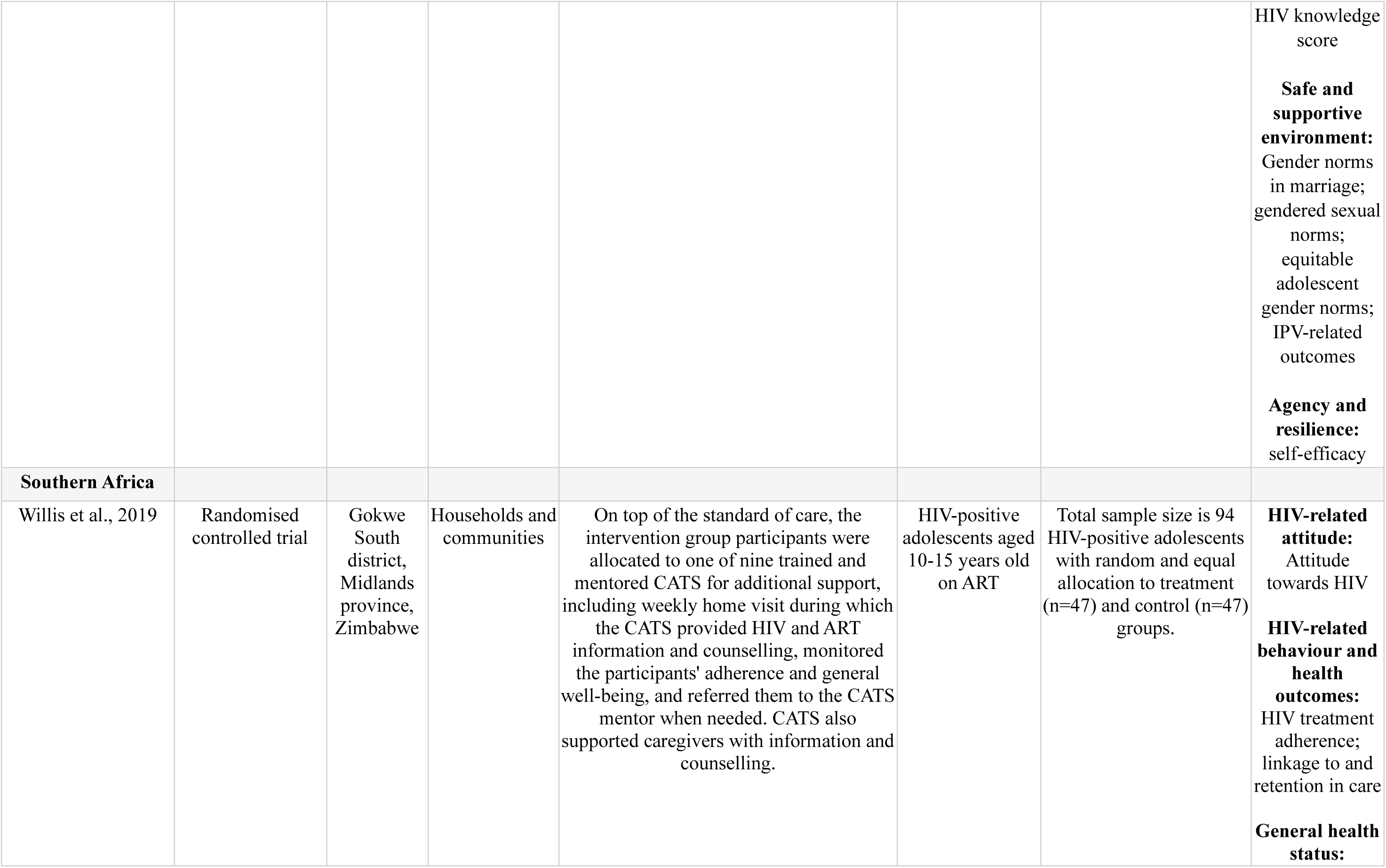
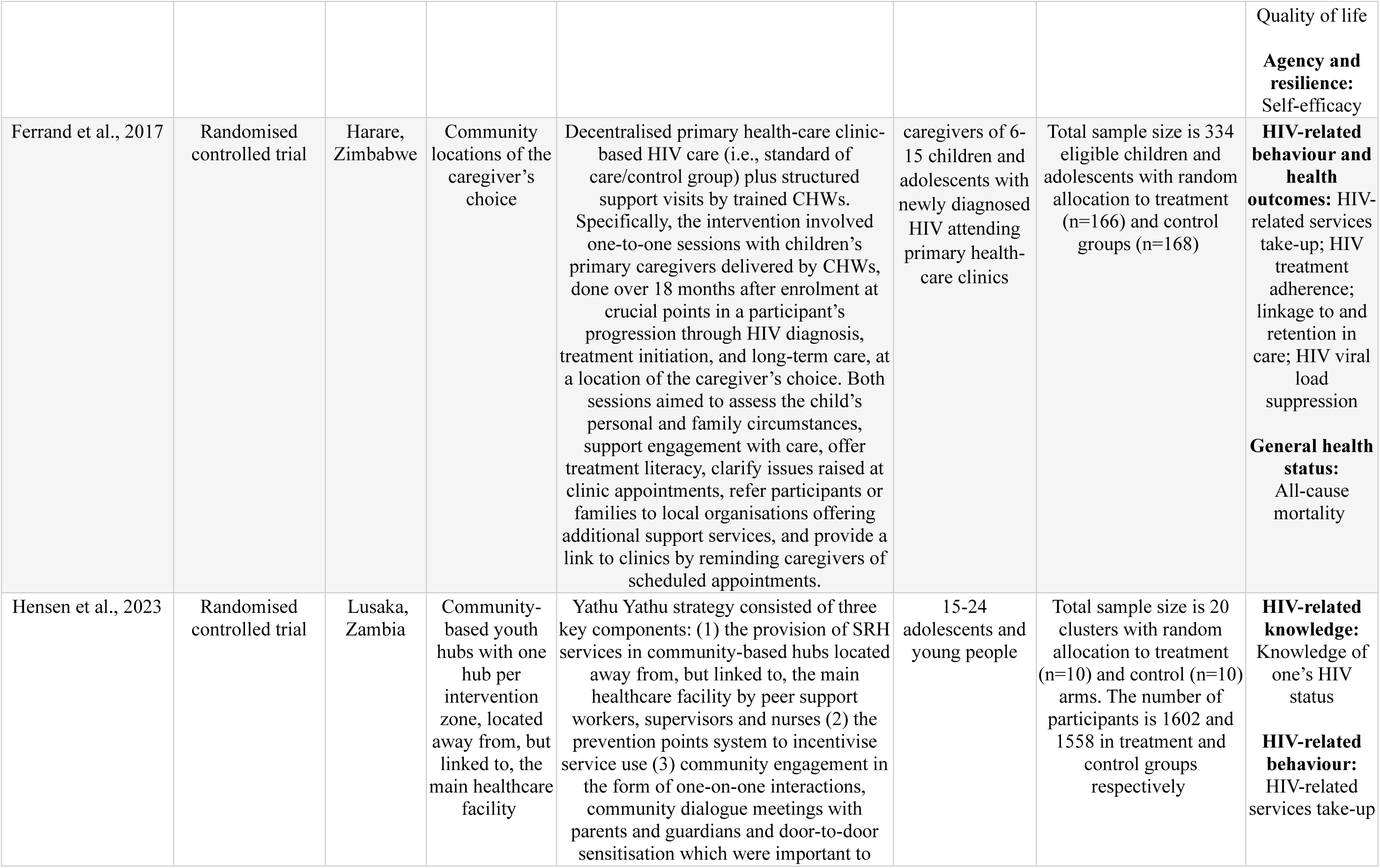
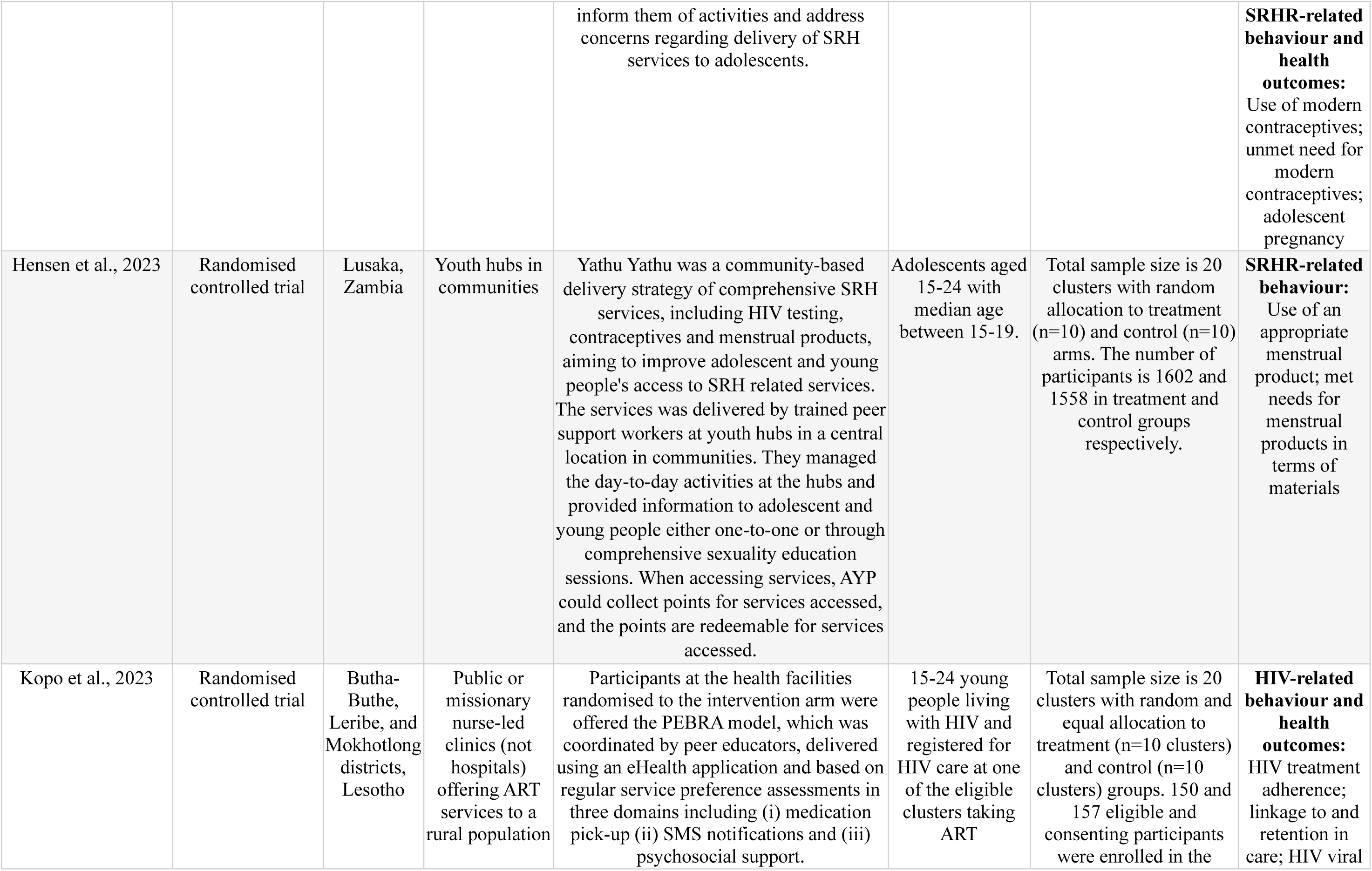
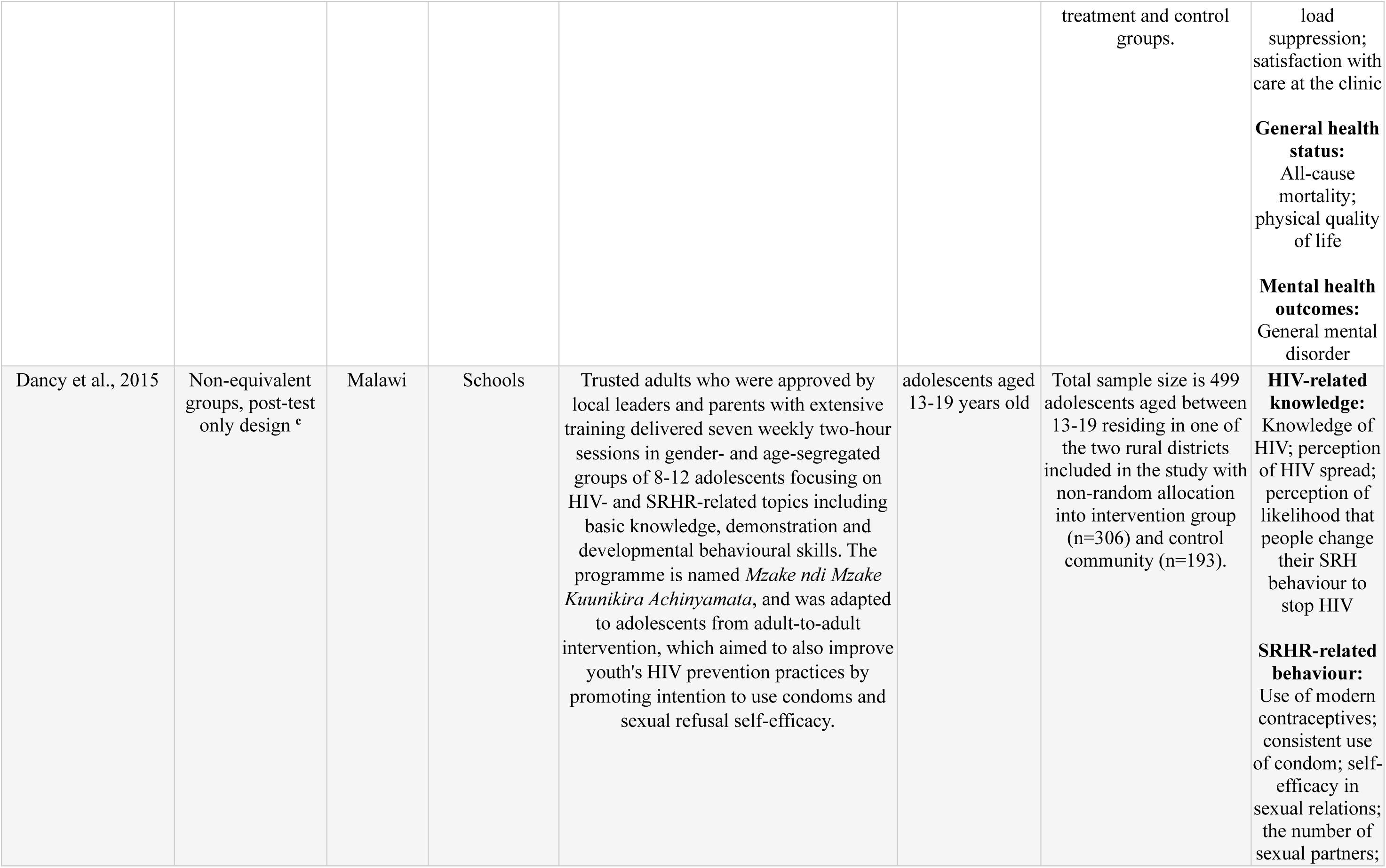
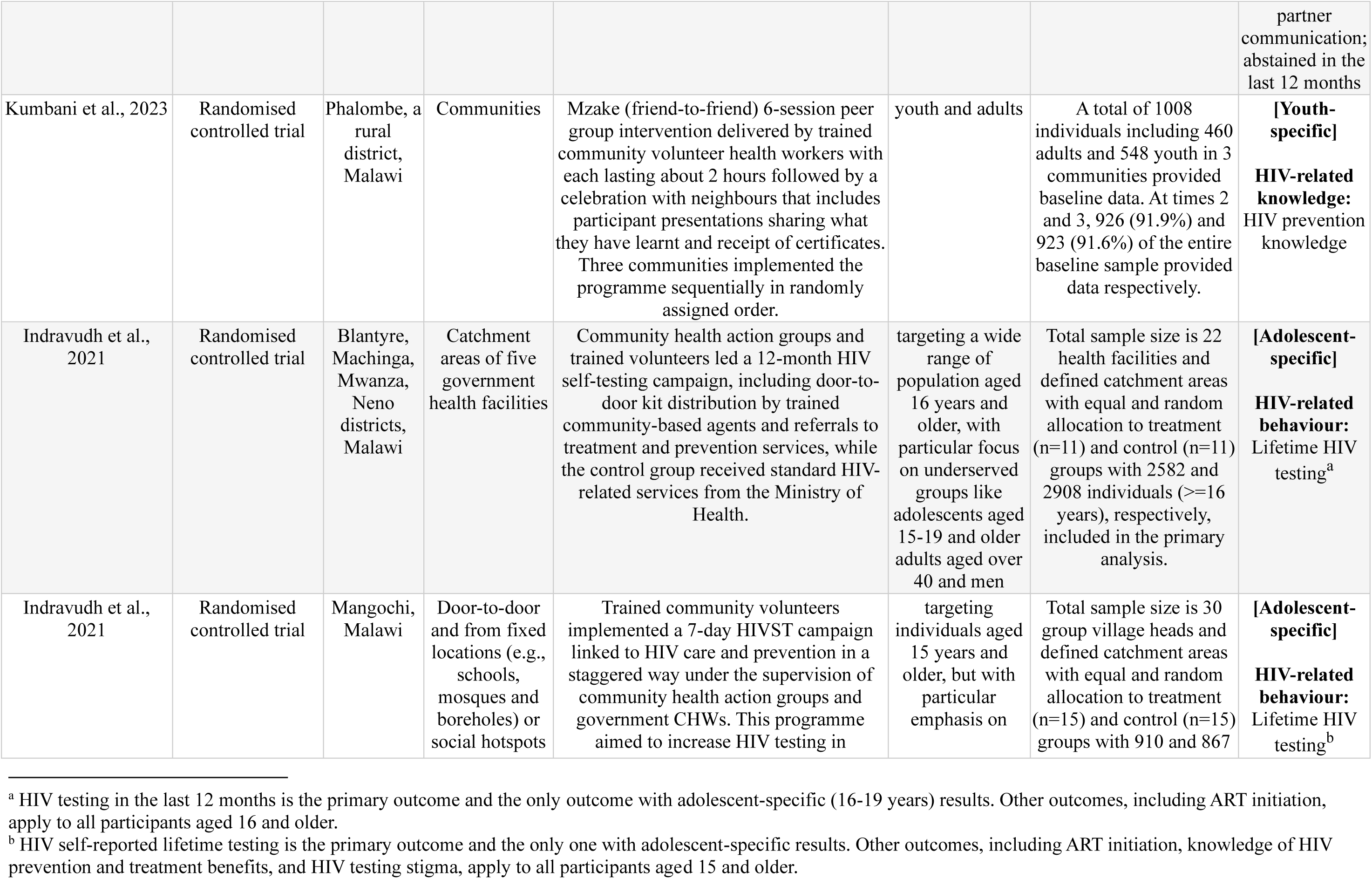
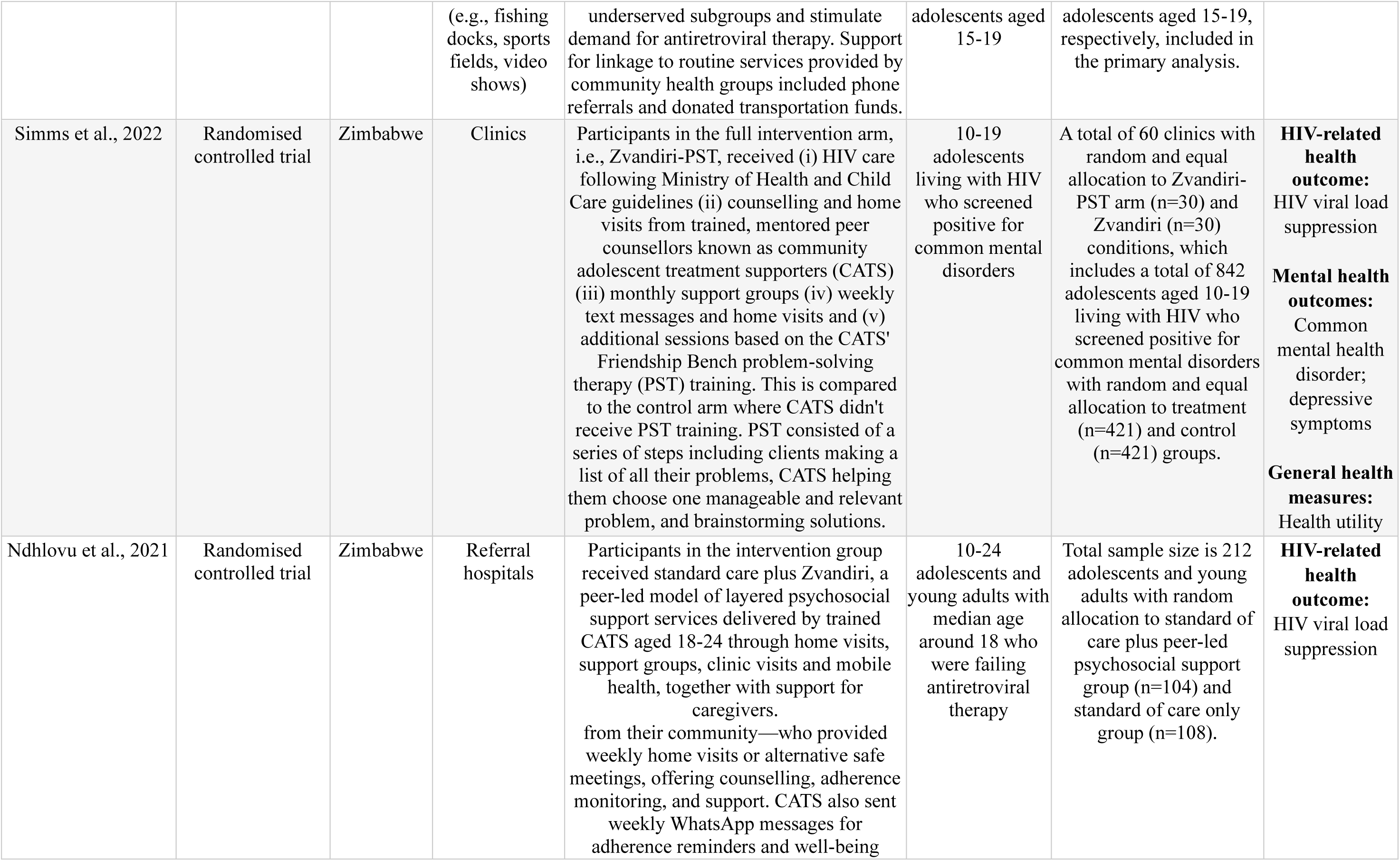
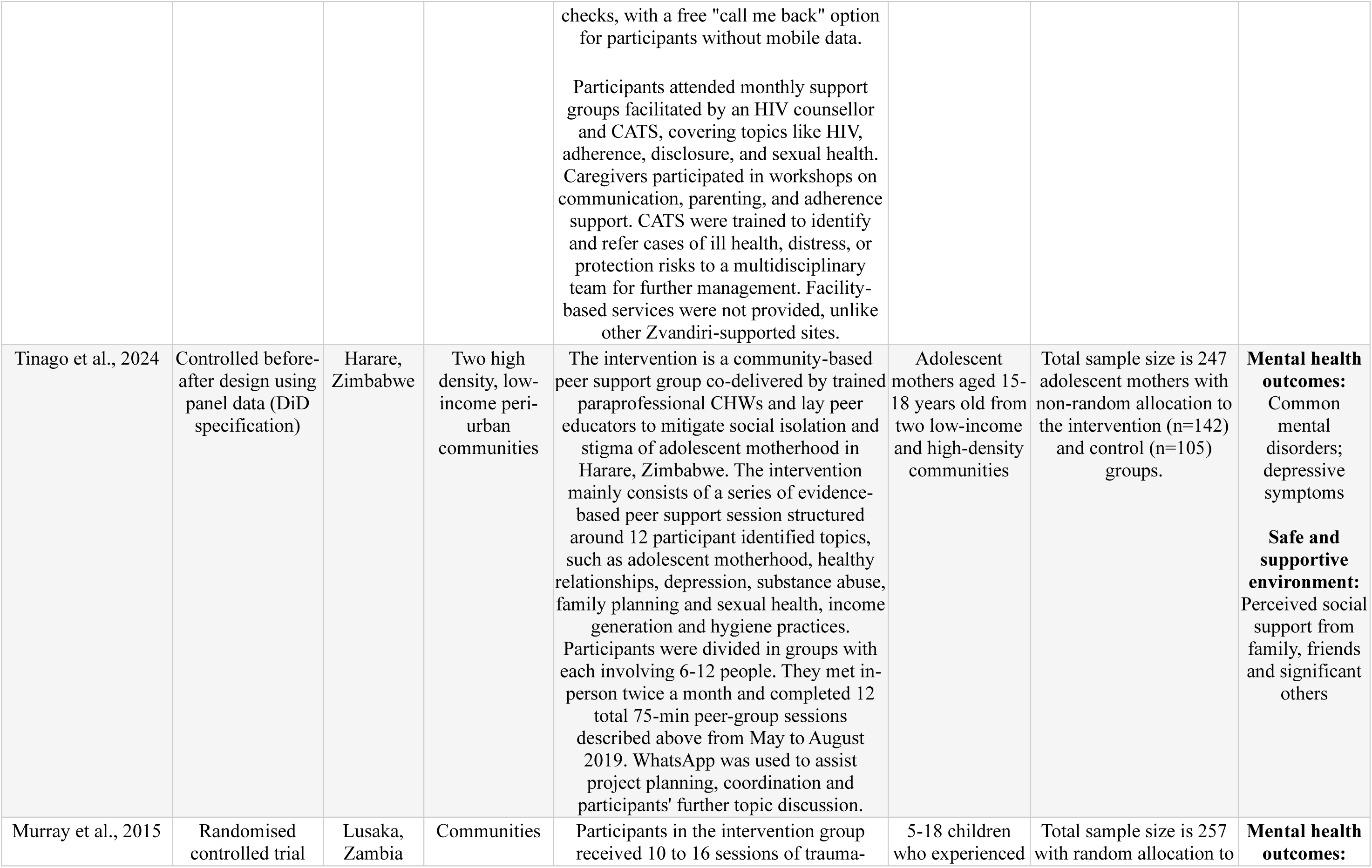
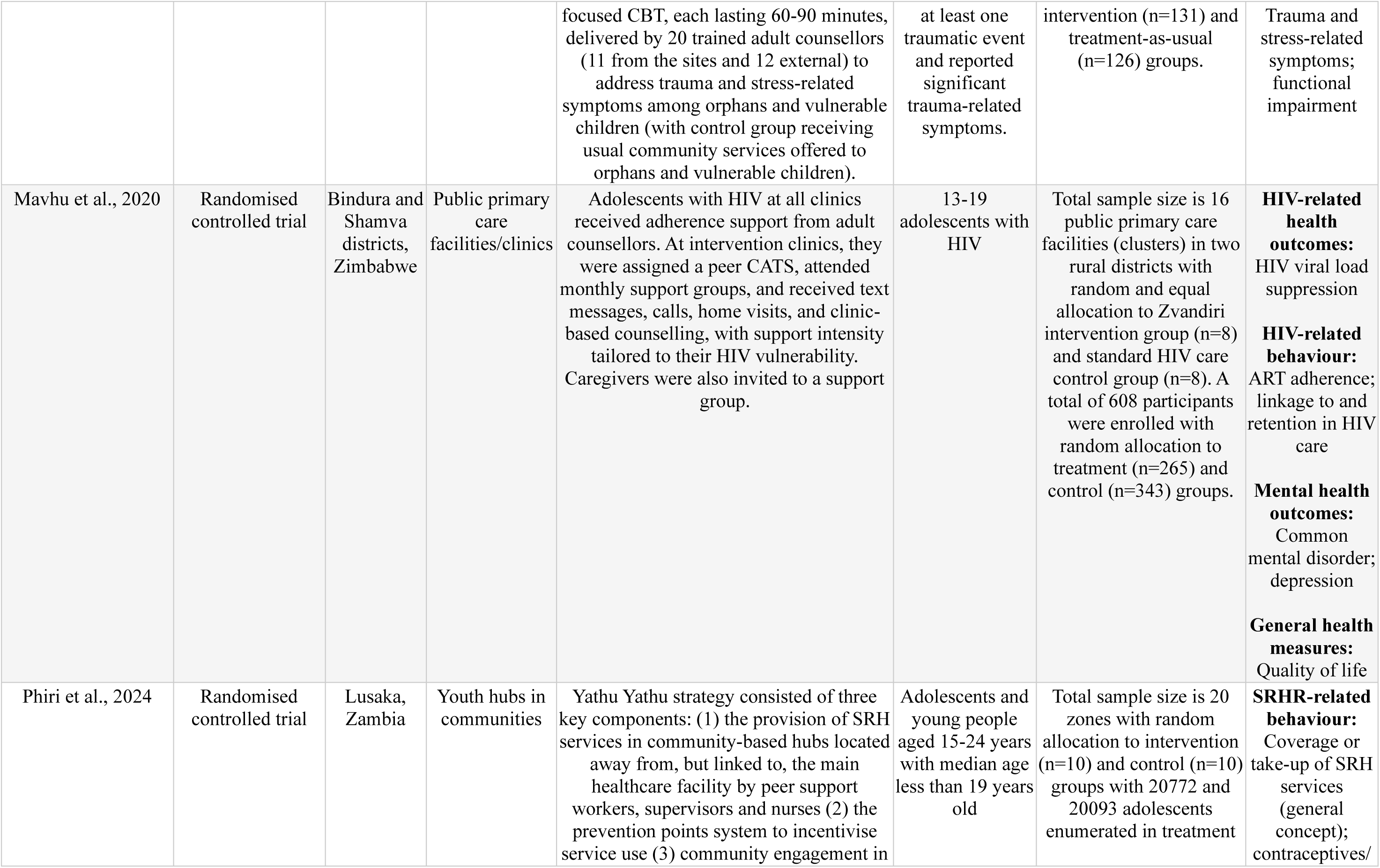
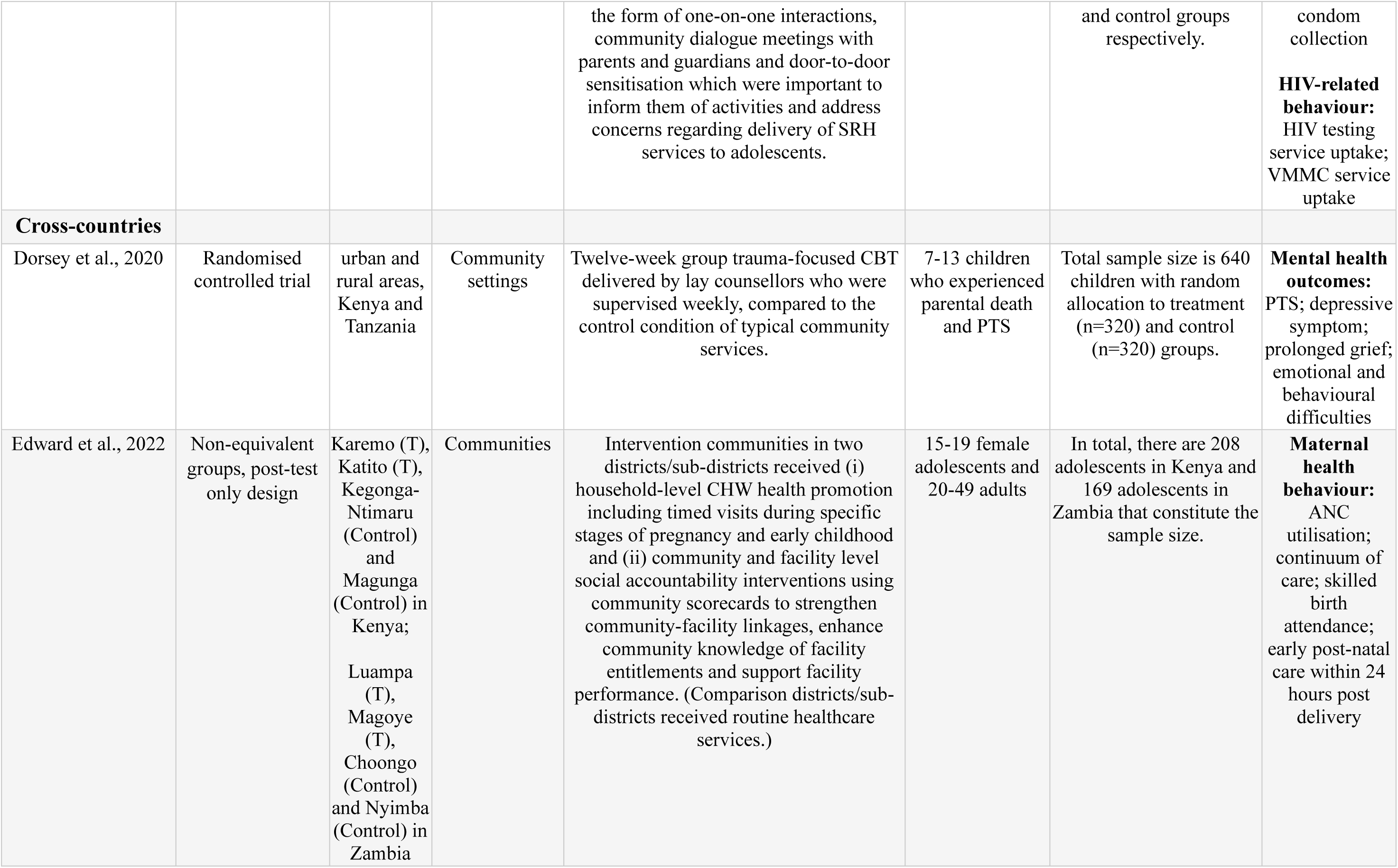
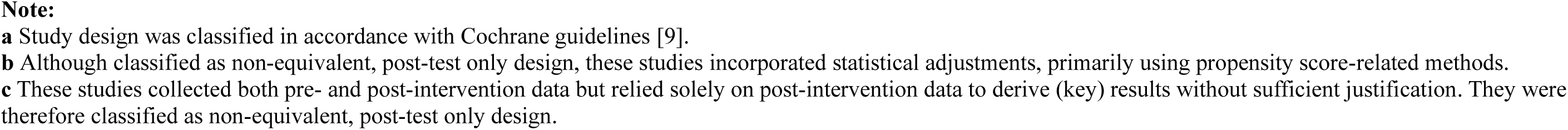

Appendix F. Description of Included Studies.
| Author, year | Study Design <sup>a</sup> | Location | Setting | Intervention | Target population | Study sample | Outcome assessed |
| --- | --- | --- | --- | --- | --- | --- | --- |
| **Eastern Africa** |  |  |  |  |  |  |  |
| Brooks et al., 2019 | Non-equivalent groups, post-test only design | Dosso Region, Niger | Home visits in rural villages | Volunteer CHWs, called relais communautaires, visited homes and performed several tasks related to health promotion, treatment and linkages with formal health care facilities. | 13-19 young married women | Total sample size is 956 young married women. | **SRHR-related behaviour:** Use of modern contraceptive methods |
| Silverman et al., 2023 | Randomised controlled trial | Dosso Region, Niger | (i) Household visits were conducted in the homes of married adolescent girls and young women and their husbands<br>(ii) Separate small group discussions were held at a health centre<br>(iii) Community dialogues were held in the village mosque or centre of each intervention village | Reaching married adolescents is a community-based, gender-synchronised programme designed to increase use of modern spacing contraception among married adolescent girls and their husbands. The intervention programme includes (i) household visits made by trained CHWs (aged 20-45) to provide education and promote uptake of modern contraceptives, (ii) small group discussions each among 10-13 participants held by trained mentors to promote dialogue around delaying and spacing births, use of contraceptive methods and relevant cultural and gender norms and (iii) community dialogues held in intervention villages to create an enabling environment and contraceptive method use. | 13-19 married adolescent girls and their husbands | A total of 48 villages across three districts was randomly allocated to four study conditions with 16 eligible villages in each district randomly selected. Four villages in each district were randomly allocated into treatment arm 1-3 and control condition. Within each of the selected villages, 25 households were randomly selected which included a married female adolescents aged 13-19 years and her husband. | **SRHR-related behaviour:** Use of modern contraceptives<br>**Safe and supportive environment:** Intimate partner violence |
| Hassan et al., 2014 | Non-equivalent groups, post-test only design | Akwa-Ibom, Enugu, | Public secondary schools | The intervention evaluated in this study was a peer-led HIV/AIDS education initiative delivered under Nigeria's Family | Youths on national service aged 21-30 years | Total sample size is 460 with non-random allocation to intervention | **HIV-related knowledge:** HIV knowledge |
|  |  | Gombe, Kaduna, Plateau, and Osun (covering all six geo-political zones), Nigeria |  | Life and HIV Education (FLHE) program. It aimed to improve secondary school students' knowledge, attitudes, and behaviour related to HIV prevention and sexual health. The programme was delivered by National Youth Service Corps (NYSC) members trained as Peer Educator Trainers (PETs), who conducted interactive sessions in selected schools.<br>Grounded in the national FLHE curriculum, the intervention used participatory methods such as discussions and peer mentoring. PETs also trained students to become peer educators themselves, creating a ripple effect of information sharing. | and adolescents in secondary schools aged 11-19. | (n=229) and control (n=231) groups consisting of NYSC corps members. Total sample size for secondary school students is 1914 with non-random allocation to treatment (n=909) and control (n=1005) groups. | **SRHR-related knowledge:** knowledge of STIs<br>**HIV-related attitudes:** Attitudes toward HIV prevention<br>**SRHR-related behaviour:** condom use; abstinence; sexual behaviour including sexual debut, number of partners and preventive practices |
| Krug et al., 2023 | Controlled before-after design (with repeated cross-sectional pre-post) | Nasarawa and Ogun States, Nigeria | (i) Life, Family and Health mentored group sessions happened at the traditional leaders' home, in the community or at the primary health centre<br>(ii) Life, Love and Health facilitated group session | (i) Female mentors moderated Life, Family and Health class sessions which consists of four mentored group sessions conducted every two weeks whose curriculum focuses on nutrition, sexual and reproductive health, life skills and vocational skills.<br>(ii) Trained A360 young providers or government health workers conducted contraceptive counselling.<br>(iii) Male interpersonal communicators start conversations with groups of men. | 15-19 sexually active adolescent girls who were married or living as married in Nasarawa state;<br>15-19 girls who were unmarried in Ogun state | Total target sample size is 16575 (Nasarawa state n=4555, Ogun state n=12020) with non-random allocation to treatment and comparison sites | **SRHR-related knowledge and attitude:** Knowledge of and attitude towards modern contraceptives/condom; knowledge of SRHR-related services<br>**SRHR-related behaviour:** |
|  |  |  | took place at primary healthcare clinics<br>(ii) Contraceptive counselling was offered at service delivery points at health facilities<br>(iii) Male interpersonal communicators reach husbands of adolescent girls in private outdoor spaces such as traditional joints and places where men gather to relax | (i) Female mentors moderated one Life, Love and Health group session whose curriculum features vocational skills, future-planning exercises, and discussions about love, sex, and dating.<br>(ii) Trained A360 young providers or government health workers conducted individual contraceptive counselling for girls in public health clinics, and engage mothers of adolescent girls through two sessions to help mothers understand that contraception can be a tool to help girls achieve their dreams. These sessions also aim to dispel myths and misconceptions around contraception. |  |  | Use of modern contraceptives; consistent use of condom; sexual self-efficacy; unmet need for contraception<br>**SRHR-related health outcome:** Adolescent pregnancy<br>**Safe and supportive environment:** Positive support from peer, family and community |
| Betancourt et al., 2014 | Randomised controlled trial | Sierra Leone | Group sessions were delivered at 6 community-based sites throughout Freetown with help from local leaders, which usually involved community meeting spaces or school | Youth Readiness Intervention (YRI) is a group-based psychosocial programme designed to help war-affected youth succeed in education and employment by improving emotional regulation and interpersonal functioning. Developed using components of evidence-based cognitive-behavioural therapy and group interpersonal therapy, YRI targets co-occurring mental health symptoms and functional difficulties without exposure-based trauma processing. Instead, it incorporates trauma psychoeducation and | Multisymptomatic war-affected youth aged 15-24 years that show psychological distress and self-reported impairment in daily functioning | Total sample size is 436 with random allocation to Youth Readiness Intervention group (n=222) and control condition (n=214). After treatment, these youth were again randomly allocated to receive an education subsidy either immediately (n=220) or waitlisted (n=216). | **Mental health outcome:** Emotion regulation; psychological distress; functional impairment; posttraumatic stress disorder (PTSD) symptoms |
|  |  |  | buildings used during off hours | discussion of trauma's impact on relationships and self-concept.<br>The intervention was delivered by trained and supervised lay local mental health workers, stratified by age and gender. It consisted of 10–12 weekly group sessions (90 minutes each) over 10 weeks. Core components included trauma psychoeducation, self-regulation and relaxation techniques (e.g., deep breathing), cognitive restructuring, behavioural activation, communication and interpersonal skills, and problem-solving. Family and community meetings were used, where appropriate, to strengthen engagement and support. All content was culturally and developmentally adapted. |  |  | **Behavioural adjustment:**<br>Prosocial behaviour<br>**Safe and supportive environment:**<br>Social support |
| **Eastern Africa** |  |  |  |  |  |  |  |
| Osborn et al., 2020 | Randomised controlled trial | Kibera (an urban slum located 4.1 miles from the city centre of Nairobi), Nairobi, Kenya | Mixed-gender secondary schools | Grounded in the national FLHE curriculum, the intervention used participatory methods such as discussions and peer mentoring. PETs also trained students to become peer educators themselves, creating a ripple effect of information sharing. The intervention was implemented across six states, with both urban and rural schools included, and compared to schools without the program to assess its impact. | Adolescents aged between 12 and 19 who reported elevated depression or anxiety. | Total sample size is 51 adolescents with random allocation to the intervention (n=28) and control (n=23) groups. | **Mental health outcomes:**<br>Anxiety; depression<br>**Learning, education and employment:**<br>Academic performance<br>**Safe and supportive environment:**<br>social support<br>Agency and resilience: |
|  |  |  |  |  |  |  | Perceived control |
| Tol et al., 2014 | Randomised controlled trial | Bubanza and Cibitoke provinces, Burundi | Schools | Trained and supervised paraprofessional CHWs delivered the classroom-based intervention as part of a multi-layered school-based care package. This package also included universal prevention activities, such as structured social events and targeted mental health treatments, aiming to reduce psychological symptoms and strengthen protective factors in at-risk children. The intervention consisted of 15 group sessions over 5 weeks, incorporating psychoeducation, coping skills, and creative discussions of posttraumatic experiences with groups of approximately 15 children or adolescents. | War-affected children/adolescents aged 8 to 17 with a mean age being 12.29 years. | Total sample size is 329 with random allocation to treatment group (n=153) and control/waitlisted condition(n=176) in 14 selected schools in treatment (n=7 schools) and control (n=7 schools) conditions. | **Mental health outcomes:**<br>PTSD;<br>depressive symptoms;<br>functional impairment<br>**Agency and resilience:**<br>Hope |
| Brathwaite et al., 2022 | Randomised controlled trial | Masaka region, Uganda | Schools | The Amaka Amasanyufu intervention is a 16-week multiple family group (MFG) programme designed to reduce disruptive behaviours and improve mental health in school-going children in low-resource communities. Culturally adapted from the '4Rs and 2Ss' model for the Ugandan context, it was delivered in schools by trained parent peers or CHWs through interactive sessions lasting between 60-90 minutes held normally during breaks on school days and occasionally during weekends. Sessions included group discussions, role plays and interactive activities to facilitate support, learning and interaction both within family members and among the families in the group. Facilitators were trained in both content and facilitation skills, with training focused on task-shifting to | children/adolescents aged 8-13 | Total sample size is 26 schools with random allocation to control (n=10), parent peer-delivered intervention (n=8) and CHW-delivered intervention (n=8) groups.<br>In total, 636 children/adolescents screened positive for oppositional defiant disorder and impaired functioning were randomly allocated to control (n=243), parent peer-delivered intervention (n=194) and CHW-delivered intervention (N=199). | **Mental health outcomes:**<br>Oppositional defiant disorder (ODD);<br>functional impairment<br>**Healthy relationships:**<br>Relationship with siblings, peers and parents<br>**Learning, education and employment:**<br>Academic performance |
|  |  |  |  | address the shortage of trained health professionals. |  |  | **Agency and resilience:**<br>Self-esteem<br>**Safe and supportive environment:**<br>Family functioning |
| Dow et al., 2020 | Randomised controlled trial | Moshi, Tanzania | A centrally located site not affiliated with HIV care | Ten group sessions (two of which were held jointly with caregivers) and two individual sessions that lasted approximately 90 min each were held on Saturdays by six trained young adults aged 23 to 30 years (who live with HIV and/or have prior experience with mental health research) plus standard of care which includes usual monthly attendance to the adolescent HIV clinic, routine adherence counselling in the form of health talks. | 12-24 young people living with HIV | Total sample size is 105 young people living with HIV with random allocation to treatment (n=58) and control/standard-of-care (n=47) groups. | **Mental health outcomes:**<br>PTS; depressive symptoms; emotional and behavioural symptoms and difficulties<br>**HIV-related attitude:**<br>Attitude towards HIV<br>**HIV-related behaviour and outcomes:**<br>ART adherence; HIV viral load suppression |
| Abamecha et al., 2021 | Non-equivalent groups, post-test only design (with statistical adjustment) <sup>b</sup> | Jimma, Ethiopia | Primary school and household or community | Peer education combined with SBCC interventions consist of various capacity building and educational sessions that were implemented from 2017 to 2019 engaging 75 schools and respective villages. Regular peer educations was conducted every two weeks, mediated by flip chart. Talented students were encouraged to prepare and disseminate | Primary school children with the majority of them aged fewer than 14 years | Total sample size is 798 with non-random allocation to intervention (n=399) and control (n=399) groups. | **Malaria-related behaviour:**<br>insecticide-treated nets (ITNs) utilisation |
|  |  |  |  | malaria messages through folks like poems, roleplays, songs on mini-media sessions and campaign days. Students were then guided to take malaria messages to parents through improved student-parent communication channels. This was mostly achieved through a card and signature. |  |  | **Malaria-related knowledge, attitudes and perception:** Knowledge, attitude, perceived risk, perceived severity regarding malaria<br>**Safe and supportive environment:** Perceived family support for use of malaria preventive measures<br>**Agency and resilience:** Self-efficacy |
| Bakesiima et al., 2021 | Randomised controlled trial | Northern Uganda | Community meeting place | Adolescents in the intervention arm received individual peer counselling on contraceptives in privacy from three trained female adolescents using a standardised WHO guide. Counsellors discussed contraceptive methods' pros, cons, safety and effectiveness, addressed questions and myths, and used physical samples to demonstrate usage, including implant and IUD insertion. Sessions lasted 15–20 minutes. (Control group participants received routine contraceptive<br>counselling, the standard of care, within the refugee settlement.) | Female refugees aged 15-19 | Total sample size is 588 with equal and random allocation to treatment (n=294) and control (n=294) groups. | **SRHR-related attitudes:** Acceptance of modern contraceptive methods |
| Menna et al., 2015 | Non-equivalent groups, post-test only design <sup>c</sup> | Addis Ababa, Ethiopia | Secondary schools | Peer education interventions were delivered by trained student facilitators nominated by their peers, who conducted two sessions per week, either after regular school hours or during free periods, each lasting at least 40 minutes. | Secondary school students with a majority of the students aged between 15-18 | Total sample size is 560 secondary school students with non-random allocation to treatment (n=280) and control (n=280) groups. | **HIV-related knowledge and attitude:**<br>Knowledge about HIV/AIDS;<br>Attitudes towards HIV counselling and testing<br>**HIV-related behaviour:**<br>Behaviour related to HIV counselling and testing<br>**SRHR-related behaviour:**<br>Use of modern contraceptives; the number of sexual partners; sexual intercourse initiation |
| Rudgard et al., 2021 | Non-equivalent groups, post-test only design (with statistical adjustment) <sup>b</sup> | Amhara; Oromia; Southern Nations, Nationalities, and Peoples'; | Communities | Predominantly female health extension workers ($\geq 17$ years old) dedicated 25% of their time in health post and 75% of their time to community health promotion through household visits, community meetings and school health services. Adult women were the most commonly contacted household members by HEWs. | Young Lives index children from age 7.5-8.5 to 18.5-19.5 | Total sample size for analysis is 775. | **General health measures:**<br>Anthropometric outcome; general health status |
|  |  | and Tigray,<br>Ethiopia |  | Three most common health promotion services delivered by HEWs are cross-cutting health education, child vaccination and family planning. |  |  | **Safe and supportive environment:**<br>Child labour;<br>child marriage<br>**Alcohol use:**<br>Alcohol use<br>**SRHR-related knowledge:**<br>Knowledge of fertility;<br>knowledge of STI<br>**SRHR-related health outcome:**<br>Adolescent pregnancy<br>**Learning, education and employment:**<br>Literacy;<br>numeracy;<br>education enrolment |
| Wondimagegene et al., 2024 | Randomised controlled trial | Gedeo Zone,<br>Ethiopia | Schools | Selected students acted as peer educators to disseminate knowledge and information on pregnancy and modern contraception to their peers using participatory classroom teaching techniques and via exciting structures, such as school clubs. They were aided by booklet, flipchart and poster. The peer education session | 15-19 school-going girls | Total sample size is 6 schools/clusters with random allocation to treatment (n=3) and control (n=3). The total number of adolescent girls is 224 with random allocation to intervention | **SRHR-related behaviour:**<br>Use of modern contraceptives; unmet need for modern contraceptives; demand for |
|  |  |  |  | happened once weekly for one hour and lasted at least six months. |  | (n=113) and control (n=111). | modern contraception |
| Ssewamala et al., 2023 | Randomised controlled trial | Uganda | Schools | The Amaka Amasanyufu intervention is a 16-week multiple family group (MFG) programme designed to reduce disruptive behaviours and improve mental health in school-going children in low-resource communities. Culturally adapted from the '4Rs and 2Ss' model for the Ugandan context, it was delivered in schools by trained parent peers or CHWs through interactive sessions lasting between 60-90 minutes held normally during breaks on school days and occasionally during weekends. Sessions included group discussions, role plays and interactive activities. Facilitators were trained in both content and facilitation skills, with training focused on task-shifting to address the shortage of trained health professionals. | 8-13 child/adolescent in grades 2-7 testing positive for behavioural problems | Total sample size is 26 schools with random allocation to MFG intervention delivered via parent peers (MFG-PP) (n=8), MFG delivered via community healthcare workers (MFG-CHW) (n=8) and control group (n=10). There are 636 children positive for disruptive behavioural disorders in total, randomly allocated to MFG-PP (n=194), MFG-CHW (n=199) and control (n=243) groups. | **Behavioural adjustment**<br>Behaviour disorders<br>**Mental health outcomes:**<br>Impaired functioning; depression; self-concept |
| Posso et al., 2021 | Controlled before-after design using panel data (DiD specification) | Amhara; Oromia; Southern Nations, Nationalities, and Peoples'; and Tigray, Ethiopia | Communities | Trained young women health extension workers conduct home visits and outreach activities with the remainder of their time working at a health post. They promote health via education, screening, prevention and selective clinical interventions. Health education and prevention focuses on injury and accident prevention with an emphasis on children. | 5 -15 children using Young Lives rounds 2-5 for the younger cohort and rounds 2-3 for older cohort | Sample size in the baseline model includes 5587 observations from 2255 children from older cohort (n=693) and younger cohort (n=1562). | **Safe and supportive environment:**<br>Child labour |
| Tamiru et al., 2016 | Non-equivalent groups, post-test only design <sup>c</sup> | Jimma Zone, Oromia Region, Ethiopia | Four primary schools | A peer-led behavioural change and health promotion programme was implemented in schools over 8 months. Intervention schools received school-based health and nutrition education using peer groups, school media and health clubs, with family and community participation. To support the programme, communication materials and a mini media system was provided to both intervention and control schools, fostering an enabling school health environment. | Adolescents aged 10-19 | Total sample size 1000 with non-random but equal allocation to intervention (n=500) and control (n=500) groups. | **Nutrition:**<br>Dietary diversity |
| Austrian et al., 2021 | Randomised controlled trial | Kilifi County, Kenya | Grade 7 primary schools | The Nia project, delivered by trained CHWs, mainly included monthly delivery of sanitary pads and a 25-session curriculum during girls'-only health clubs held during time allocated for extra-curricular activities in schools. The curriculum covered a range of topics including puberty, menstrual health management, reproductive systems, self-esteem, gender, human rights, power dynamics, sexual violence, assertiveness, decision-making, relationships, teen pregnancy, STIs and HIV, peer pressure, drug use and conflict management. Girls also received two pairs of underwear at the start of the intervention, an additional pair at the end of each subsequent school term and a magazine designed to appeal to adolescent girls and convey core RH messaging through storytelling using aspirational personal stories. | Adolescent girls enrolled in primary school grade 7 with median age being around 14 years old. | Total sample size is 140 primary schools with random and equal allocation to four study arms including (i) control setting (ii) sanitary pad distribution intervention group (iii) reproductive health education intervention group and (iv) both sanitary pad distribution and reproductive health education intervention group. In total, there are 3489 randomly selected girls in primary grade 7 allocated to four study arms as follows: (i) n=669, (ii) n=682, (iii) n=677 and (iv) n=697. | **Learning, education and employment:**<br>School engagement<br>**SRHR-related attitude:**<br>Attitude towards menstruation<br>**SRHR-related health outcomes:**<br>Has enough pads; reporting leaking<br>**SRHR-related knowledge:**<br>Knowledge of pregnancy; knowledge of STIs; knowledge of modern contraceptives<br>**HIV-related knowledge:** |
|  |  |  |  |  |  |  | HIV knowledge score<br>**Safe and supportive environment:**<br>Gender norms in marriage; gendered sexual norms; equitable adolescent gender norms; IPV-related outcomes<br>**Agency and resilience:**<br>self-efficacy |
| **Southern Africa** |  |  |  |  |  |  |  |
| Willis et al., 2019 | Randomised controlled trial | Gokwe South district, Midlands province, Zimbabwe | Households and communities | On top of the standard of care, the intervention group participants were allocated to one of nine trained and mentored CATS for additional support, including weekly home visit during which the CATS provided HIV and ART information and counselling, monitored the participants' adherence and general well-being, and referred them to the CATS mentor when needed. CATS also supported caregivers with information and counselling. | HIV-positive adolescents aged 10-15 years old on ART | Total sample size is 94 HIV-positive adolescents with random and equal allocation to treatment (n=47) and control (n=47) groups. | **HIV-related attitude:**<br>Attitude towards HIV<br>**HIV-related behaviour and health outcomes:**<br>HIV treatment adherence; linkage to and retention in care<br>**General health status:** |
|  |  |  |  |  |  |  | Quality of life |
|  |  |  |  |  |  |  | **Agency and resilience:**<br>Self-efficacy |
| Ferrand et al., 2017 | Randomised controlled trial | Harare, Zimbabwe | Community locations of the caregiver's choice | Decentralised primary health-care clinic-based HIV care (i.e., standard of care/control group) plus structured support visits by trained CHWs. Specifically, the intervention involved one-to-one sessions with children's primary caregivers delivered by CHWs, done over 18 months after enrolment at crucial points in a participant's progression through HIV diagnosis, treatment initiation, and long-term care, at a location of the caregiver's choice. Both sessions aimed to assess the child's personal and family circumstances, support engagement with care, offer treatment literacy, clarify issues raised at clinic appointments, refer participants or families to local organisations offering additional support services, and provide a link to clinics by reminding caregivers of scheduled appointments. | caregivers of 6-15 children and adolescents with newly diagnosed HIV attending primary health-care clinics | Total sample size is 334 eligible children and adolescents with random allocation to treatment (n=166) and control groups (n=168) | **HIV-related behaviour and health outcomes:** HIV-related services take-up; HIV treatment adherence; linkage to and retention in care; HIV viral load suppression<br>**General health status:** All-cause mortality |
| Hensen et al., 2023 | Randomised controlled trial | Lusaka, Zambia | Community-based youth hubs with one hub per intervention zone, located away from, but linked to, the main healthcare facility | Yathu Yathu strategy consisted of three key components: (1) the provision of SRH services in community-based hubs located away from, but linked to, the main healthcare facility by peer support workers, supervisors and nurses (2) the prevention points system to incentivise service use (3) community engagement in the form of one-on-one interactions, community dialogue meetings with parents and guardians and door-to-door sensitisation which were important to | 15-24 adolescents and young people | Total sample size is 20 clusters with random allocation to treatment (n=10) and control (n=10) arms. The number of participants is 1602 and 1558 in treatment and control groups respectively | **HIV-related knowledge:** Knowledge of one's HIV status<br>**HIV-related behaviour:** HIV-related services take-up |
|  |  |  |  | inform them of activities and address concerns regarding delivery of SRH services to adolescents. |  |  | **SRHR-related behaviour and health outcomes:**<br>Use of modern contraceptives; unmet need for modern contraceptives; adolescent pregnancy |
| Hensen et al., 2023 | Randomised controlled trial | Lusaka, Zambia | Youth hubs in communities | Yathu Yathu was a community-based delivery strategy of comprehensive SRH services, including HIV testing, contraceptives and menstrual products, aiming to improve adolescent and young people's access to SRH related services. The services was delivered by trained peer support workers at youth hubs in a central location in communities. They managed the day-to-day activities at the hubs and provided information to adolescent and young people either one-to-one or through comprehensive sexuality education sessions. When accessing services, AYP could collect points for services accessed, and the points are redeemable for services accessed. | Adolescents aged 15-24 with median age between 15-19. | Total sample size is 20 clusters with random allocation to treatment (n=10) and control (n=10) arms. The number of participants is 1602 and 1558 in treatment and control groups respectively. | **SRHR-related behaviour:**<br>Use of an appropriate menstrual product; met needs for menstrual products in terms of materials |
| Kopo et al., 2023 | Randomised controlled trial | Butha-Buthe, Leribe, and Mokhotlong districts, Lesotho | Public or missionary nurse-led clinics (not hospitals) offering ART services to a rural population | Participants at the health facilities randomised to the intervention arm were offered the PEBRA model, which was coordinated by peer educators, delivered using an eHealth application and based on regular service preference assessments in three domains including (i) medication pick-up (ii) SMS notifications and (iii) psychosocial support. | 15-24 young people living with HIV and registered for HIV care at one of the eligible clusters taking ART | Total sample size is 20 clusters with random and equal allocation to treatment (n=10 clusters) and control (n=10 clusters) groups. 150 and 157 eligible and consenting participants were enrolled in the treatment and control groups. | **HIV-related behaviour and health outcomes:**<br>HIV treatment adherence; linkage to and retention in care; HIV viral<br>load suppression; satisfaction with care at the clinic<br>**General health status:**<br>All-cause mortality; physical quality of life<br>**Mental health outcomes:**<br>General mental disorder |
| Dancy et al., 2015 | Non-equivalent groups, post-test only design <sup>c</sup> | Malawi | Schools | Trusted adults who were approved by local leaders and parents with extensive training delivered seven weekly two-hour sessions in gender- and age-segregated groups of 8-12 adolescents focusing on HIV- and SRHR-related topics including basic knowledge, demonstration and developmental behavioural skills. The programme is named *Mzake ndi Mzake Kuunikira Achinyamata*, and was adapted to adolescents from adult-to-adult intervention, which aimed to also improve youth's HIV prevention practices by promoting intention to use condoms and sexual refusal self-efficacy. | adolescents aged 13-19 years old | Total sample size is 499 adolescents aged between 13-19 residing in one of the two rural districts included in the study with non-random allocation into intervention group (n=306) and control community (n=193). | **HIV-related knowledge:**<br>Knowledge of HIV; perception of HIV spread; perception of likelihood that people change their SRH behaviour to stop HIV<br>**SRHR-related behaviour:**<br>Use of modern contraceptives; consistent use of condom; self-efficacy in sexual relations; the number of sexual partners; |
|  |  |  |  |  |  |  | partner communication; abstained in the last 12 months |
| Kumbani et al., 2023 | Randomised controlled trial | Phalombe, a rural district, Malawi | Communities | Mzake (friend-to-friend) 6-session peer group intervention delivered by trained community volunteer health workers with each lasting about 2 hours followed by a celebration with neighbours that includes participant presentations sharing what they have learnt and receipt of certificates. Three communities implemented the programme sequentially in randomly assigned order. | youth and adults | A total of 1008 individuals including 460 adults and 548 youth in 3 communities provided baseline data. At times 2 and 3, 926 (91.9%) and 923 (91.6%) of the entire baseline sample provided data respectively. | **[Youth-specific]**<br>**HIV-related knowledge:** HIV prevention knowledge |
| Indravudh et al., 2021 | Randomised controlled trial | Blantyre, Machinga, Mwanza, Neno districts, Malawi | Catchment areas of five government health facilities | Community health action groups and trained volunteers led a 12-month HIV self-testing campaign, including door-to-door kit distribution by trained community-based agents and referrals to treatment and prevention services, while the control group received standard HIV-related services from the Ministry of Health. | targeting a wide range of population aged 16 years and older, with particular focus on underserved groups like adolescents aged 15-19 and older adults aged over 40 and men | Total sample size is 22 health facilities and defined catchment areas with equal and random allocation to treatment (n=11) and control (n=11) groups with 2582 and 2908 individuals (>=16 years), respectively, included in the primary analysis. | **[Adolescent-specific]**<br>**HIV-related behaviour:** Lifetime HIV testing<sup>a</sup> |
| Indravudh et al., 2021 | Randomised controlled trial | Mangochi, Malawi | Door-to-door and from fixed locations (e.g., schools, mosques and boreholes) or social hotspots | Trained community volunteers implemented a 7-day HIVST campaign linked to HIV care and prevention in a staggered way under the supervision of community health action groups and government CHWs. This programme aimed to increase HIV testing in | targeting individuals aged 15 years and older, but with particular emphasis on | Total sample size is 30 group village heads and defined catchment areas with equal and random allocation to treatment (n=15) and control (n=15) groups with 910 and 867 | **[Adolescent-specific]**<br>**HIV-related behaviour:** Lifetime HIV testing<sup>b</sup> |

<sup>a</sup> HIV testing in the last 12 months is the primary outcome and the only outcome with adolescent-specific (16-19 years) results. Other outcomes, including ART initiation, apply to all participants aged 16 and older.
<sup>b</sup> HIV self-reported lifetime testing is the primary outcome and the only one with adolescent-specific results. Other outcomes, including ART initiation, knowledge of HIV prevention and treatment benefits, and HIV testing stigma, apply to all participants aged 15 and older.

|  |  |  |  |  |  |  |  |
| --- | --- | --- | --- | --- | --- | --- | --- |
|  |  |  | (e.g., fishing docks, sports fields, video shows) | underserved subgroups and stimulate demand for antiretroviral therapy. Support for linkage to routine services provided by community health groups included phone referrals and donated transportation funds. | adolescents aged 15-19 | adolescents aged 15-19, respectively, included in the primary analysis. |  |
| Simms et al., 2022 | Randomised controlled trial | Zimbabwe | Clinics | Participants in the full intervention arm, i.e., Zvandiri-PST, received (i) HIV care following Ministry of Health and Child Care guidelines (ii) counselling and home visits from trained, mentored peer counsellors known as community adolescent treatment supporters (CATS) (iii) monthly support groups (iv) weekly text messages and home visits and (v) additional sessions based on the CATS' Friendship Bench problem-solving therapy (PST) training. This is compared to the control arm where CATS didn't receive PST training. PST consisted of a series of steps including clients making a list of all their problems, CATS helping them choose one manageable and relevant problem, and brainstorming solutions. | 10-19 adolescents living with HIV who screened positive for common mental disorders | A total of 60 clinics with random and equal allocation to Zvandiri-PST arm (n=30) and Zvandiri (n=30) conditions, which includes a total of 842 adolescents aged 10-19 living with HIV who screened positive for common mental disorders with random and equal allocation to treatment (n=421) and control (n=421) groups. | **HIV-related health outcome:**<br>HIV viral load suppression<br>**Mental health outcomes:**<br>Common mental health disorder; depressive symptoms<br>**General health measures:**<br>Health utility |
| Ndhlovu et al., 2021 | Randomised controlled trial | Zimbabwe | Referral hospitals | Participants in the intervention group received standard care plus Zvandiri, a peer-led model of layered psychosocial support services delivered by trained CATS aged 18-24 through home visits, support groups, clinic visits and mobile health, together with support for caregivers.<br>from their community—who provided weekly home visits or alternative safe meetings, offering counselling, adherence monitoring, and support. CATS also sent weekly WhatsApp messages for adherence reminders and well-being<br>checks, with a free "call me back" option for participants without mobile data.<br>Participants attended monthly support groups facilitated by an HIV counsellor and CATS, covering topics like HIV, adherence, disclosure, and sexual health. Caregivers participated in workshops on communication, parenting, and adherence support. CATS were trained to identify and refer cases of ill health, distress, or protection risks to a multidisciplinary team for further management. Facility-based services were not provided, unlike other Zvandiri-supported sites. | 10-24 adolescents and young adults with median age around 18 who were failing antiretroviral therapy | Total sample size is 212 adolescents and young adults with random allocation to standard of care plus peer-led psychosocial support group (n=104) and standard of care only group (n=108). | **HIV-related health outcome:**<br>HIV viral load suppression |
| Tinago et al., 2024 | Controlled before-after design using panel data (DiD specification) | Harare, Zimbabwe | Two high density, low-income peri-urban communities | The intervention is a community-based peer support group co-delivered by trained paraprofessional CHWs and lay peer educators to mitigate social isolation and stigma of adolescent motherhood in Harare, Zimbabwe. The intervention mainly consists of a series of evidence-based peer support session structured around 12 participant identified topics, such as adolescent motherhood, healthy relationships, depression, substance abuse, family planning and sexual health, income generation and hygiene practices. Participants were divided in groups with each involving 6-12 people. They met in-person twice a month and completed 12 total 75-min peer-group sessions described above from May to August 2019. WhatsApp was used to assist project planning, coordination and participants' further topic discussion. | Adolescent mothers aged 15-18 years old from two low-income and high-density communities | Total sample size is 247 adolescent mothers with non-random allocation to the intervention (n=142) and control (n=105) groups. | **Mental health outcomes:**<br>Common mental disorders; depressive symptoms<br>**Safe and supportive environment:**<br>Perceived social support from family, friends and significant others |
| Murray et al., 2015 | Randomised controlled trial | Lusaka, Zambia | Communities | Participants in the intervention group received 10 to 16 sessions of trauma- | 5-18 children who experienced | Total sample size is 257 with random allocation to | **Mental health outcomes:** |
|  |  |  |  | focused CBT, each lasting 60-90 minutes, delivered by 20 trained adult counsellors (11 from the sites and 12 external) to address trauma and stress-related symptoms among orphans and vulnerable children (with control group receiving usual community services offered to orphans and vulnerable children). | at least one traumatic event and reported significant trauma-related symptoms. | intervention (n=131) and treatment-as-usual (n=126) groups. | Trauma and stress-related symptoms; functional impairment |
| Mavhu et al., 2020 | Randomised controlled trial | Bindura and Shamva districts, Zimbabwe | Public primary care facilities/clinics | Adolescents with HIV at all clinics received adherence support from adult counsellors. At intervention clinics, they were assigned a peer CATS, attended monthly support groups, and received text messages, calls, home visits, and clinic-based counselling, with support intensity tailored to their HIV vulnerability. Caregivers were also invited to a support group. | 13-19 adolescents with HIV | Total sample size is 16 public primary care facilities (clusters) in two rural districts with random and equal allocation to Zvandiri intervention group (n=8) and standard HIV care control group (n=8). A total of 608 participants were enrolled with random allocation to treatment (n=265) and control (n=343) groups. | **HIV-related health outcomes:**<br>HIV viral load suppression<br>**HIV-related behaviour:**<br>ART adherence; linkage to and retention in HIV care<br>**Mental health outcomes:**<br>Common mental disorder; depression<br>**General health measures:**<br>Quality of life |
| Phiri et al., 2024 | Randomised controlled trial | Lusaka, Zambia | Youth hubs in communities | Yathu Yathu strategy consisted of three key components: (1) the provision of SRH services in community-based hubs located away from, but linked to, the main healthcare facility by peer support workers, supervisors and nurses (2) the prevention points system to incentivise service use (3) community engagement in | Adolescents and young people aged 15-24 years with median age less than 19 years old | Total sample size is 20 zones with random allocation to intervention (n=10) and control (n=10) groups with 20772 and 20093 adolescents enumerated in treatment | **SRHR-related behaviour:**<br>Coverage or take-up of SRH services (general concept); contraceptives/ |
|  |  |  |  | the form of one-on-one interactions, community dialogue meetings with parents and guardians and door-to-door sensitisation which were important to inform them of activities and address concerns regarding delivery of SRH services to adolescents. |  | and control groups respectively. | condom collection<br>**HIV-related behaviour:**<br>HIV testing service uptake; VMMC service uptake |
| **Cross-countries** |  |  |  |  |  |  |  |
| Dorsey et al., 2020 | Randomised controlled trial | urban and rural areas, Kenya and Tanzania | Community settings | Twelve-week group trauma-focused CBT delivered by lay counsellors who were supervised weekly, compared to the control condition of typical community services. | 7-13 children who experienced parental death and PTS | Total sample size is 640 children with random allocation to treatment (n=320) and control (n=320) groups. | **Mental health outcomes:**<br>PTS; depressive symptom; prolonged grief; emotional and behavioural difficulties |
| Edward et al., 2022 | Non-equivalent groups, post-test only design | Karemo (T), Katito (T), Kegonga-Ntimaru (Control) and Magunga (Control) in Kenya;<br>Luampa (T), Magoye (T), Choongo (Control) and Nyimba (Control) in Zambia | Communities | Intervention communities in two districts/sub-districts received (i) household-level CHW health promotion including timed visits during specific stages of pregnancy and early childhood and (ii) community and facility level social accountability interventions using community scorecards to strengthen community-facility linkages, enhance community knowledge of facility entitlements and support facility performance. (Comparison districts/sub-districts received routine healthcare services.) | 15-19 female adolescents and 20-49 adults | In total, there are 208 adolescents in Kenya and 169 adolescents in Zambia that constitute the sample size. | **Maternal health behaviour:**<br>ANC utilisation; continuum of care; skilled birth attendance; early post-natal care within 24 hours post delivery |

**Note:**

### Appendix G. Intervention/Programme Focus, by Age of CHWs and Setting

**Figure G.1.**
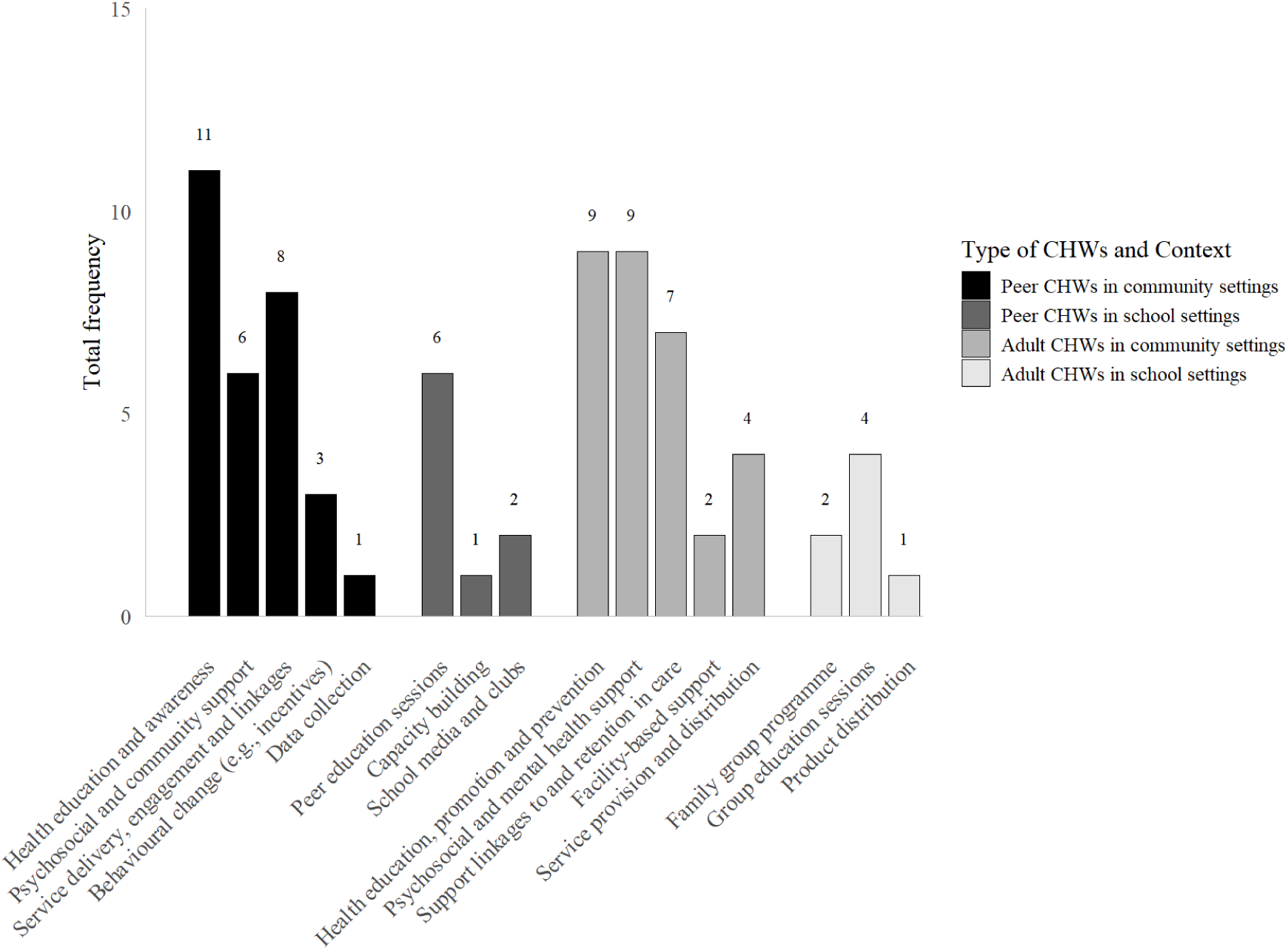
Intervention/Programme Focus, by Age of CHWs and by Intervention Setting

### Appendix H. Risk of Bias and GRADE Decisions for Eight Key Outcomes

#### Appendix H.1 Risk of Bias

**Table H.1.1.**
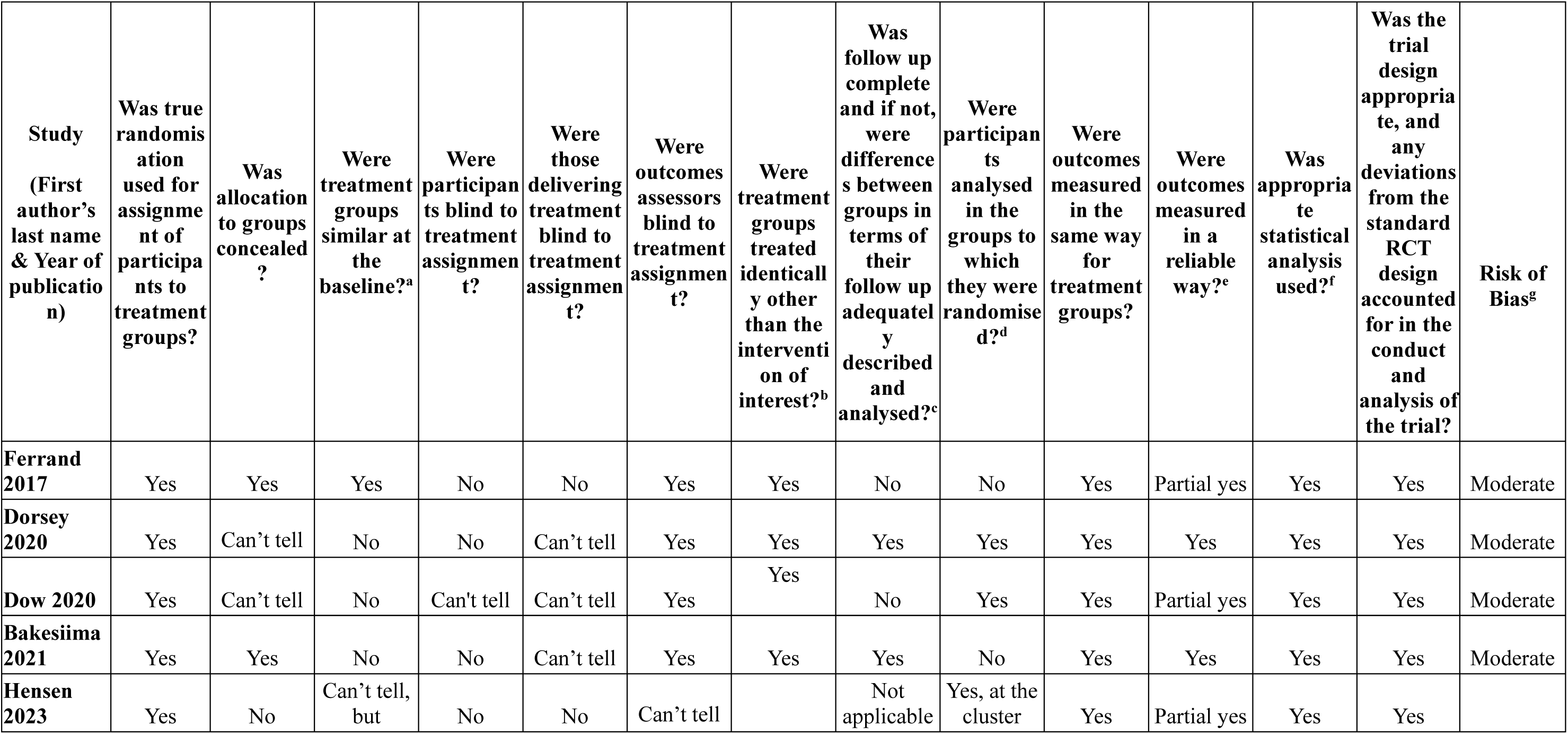
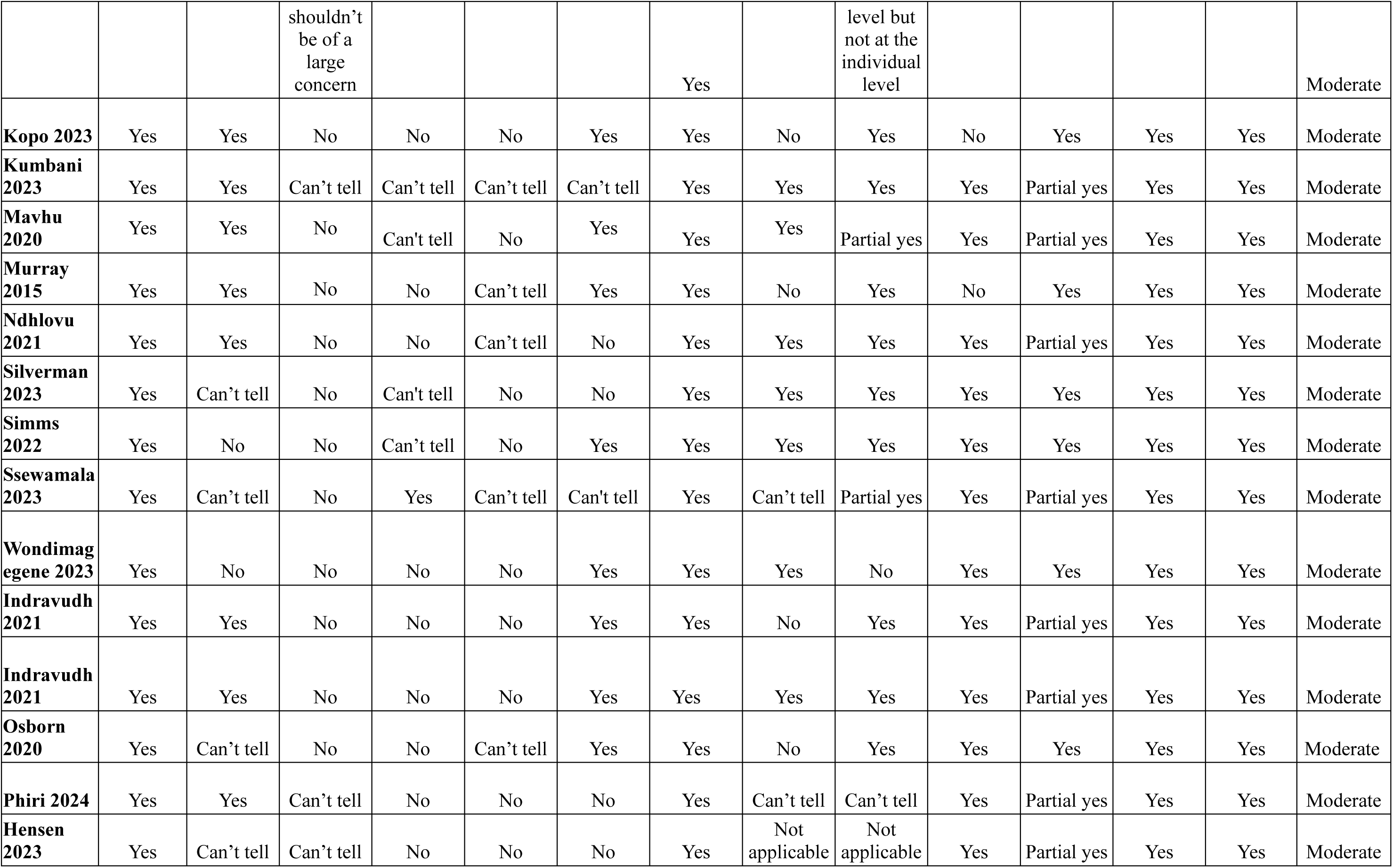
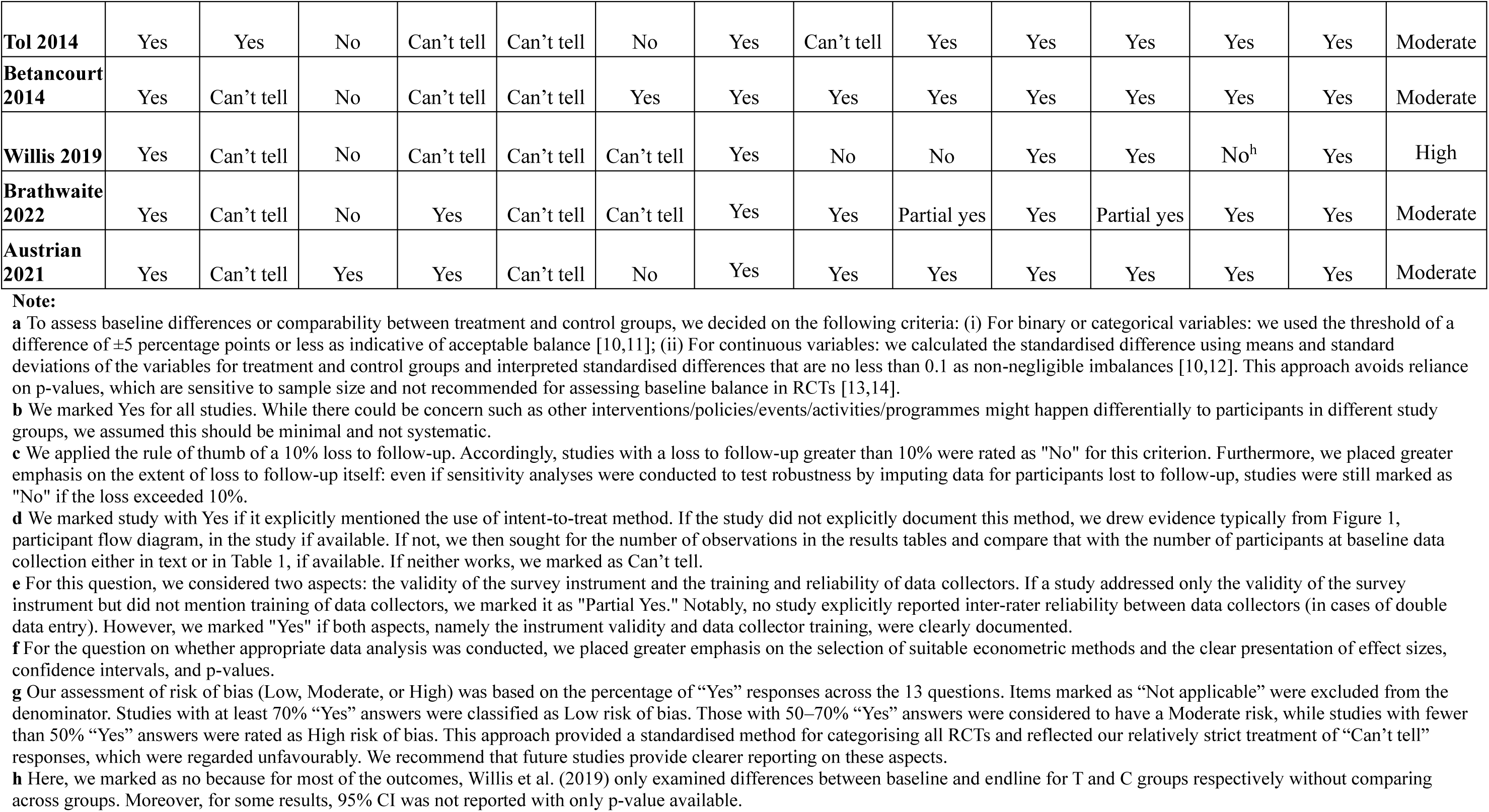
Risk of bias analysis for RCTs using the JBI tool.

**Table H.1.2.**
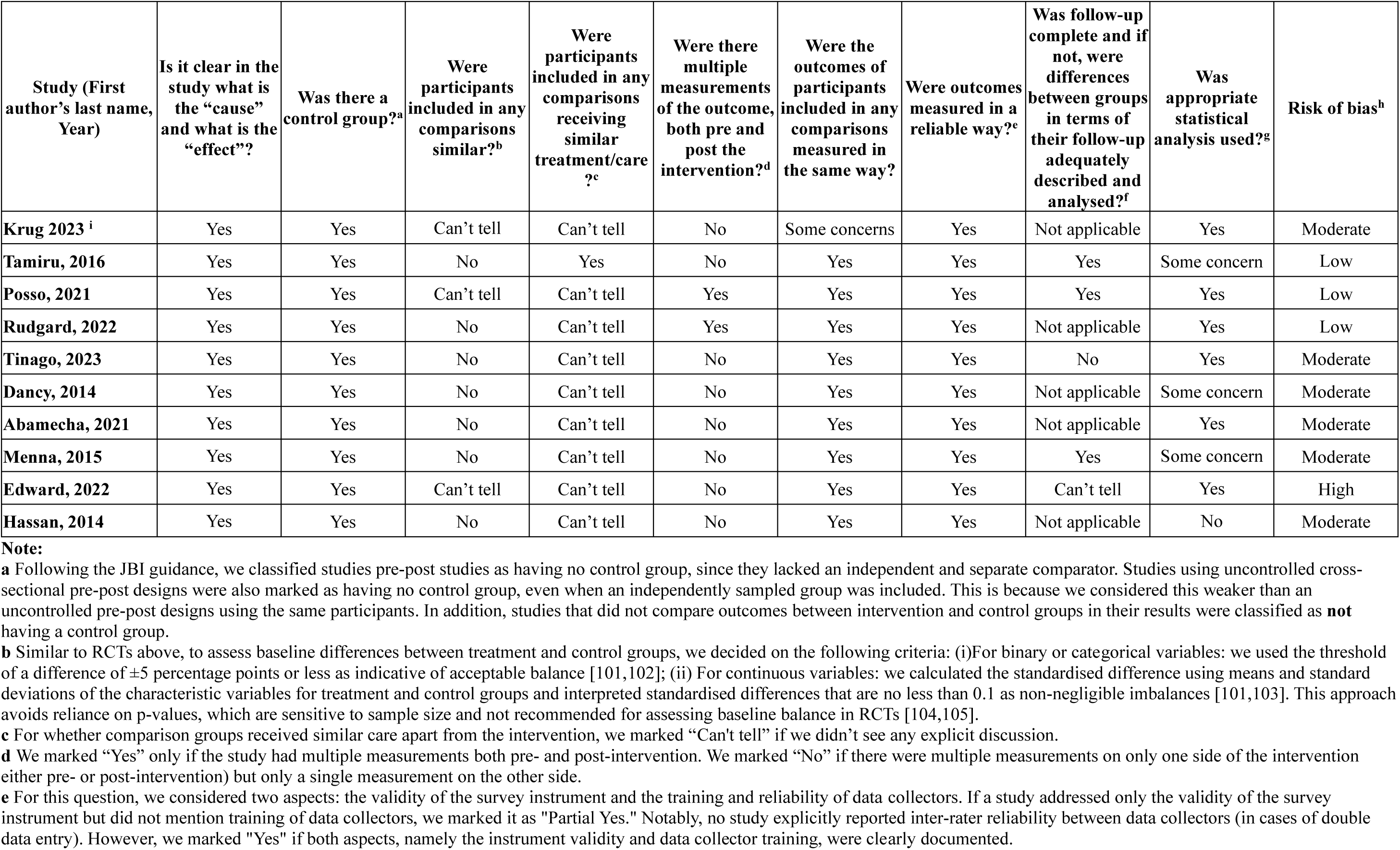
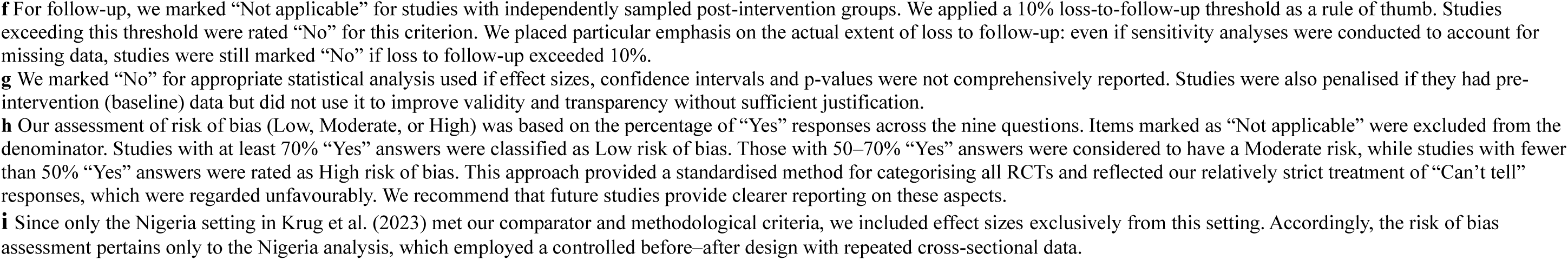
Risk of bias analysis for NRSIs using the JBI tool.

| Study (First author's last name, Year) | Is it clear in the study what is the "cause" and what is the "effect"? | Was there a control group?<sup>a</sup> | Were participants included in any comparisons similar?<sup>b</sup> | Were participants included in any comparisons receiving similar treatment/care?<sup>c</sup> | Were there multiple measurements of the outcome, both pre and post the intervention?<sup>d</sup> | Were the outcomes of participants included in any comparisons measured in the same way? | Were outcomes measured in a reliable way?<sup>e</sup> | Was follow-up complete and if not, were differences between groups in terms of their follow-up adequately described and analysed?<sup>f</sup> | Was appropriate statistical analysis used?<sup>g</sup> | Risk of bias<sup>h</sup> |
| --- | --- | --- | --- | --- | --- | --- | --- | --- | --- | --- |
| Krug 2023<sup>i</sup> | Yes | Yes | Can't tell | Can't tell | No | Some concerns | Yes | Not applicable | Yes | Moderate |
| Tamiru, 2016 | Yes | Yes | No | Yes | No | Yes | Yes | Yes | Some concern | Low |
| Posso, 2021 | Yes | Yes | Can't tell | Can't tell | Yes | Yes | Yes | Yes | Yes | Low |
| Rudgard, 2022 | Yes | Yes | No | Can't tell | Yes | Yes | Yes | Not applicable | Yes | Low |
| Tinago, 2023 | Yes | Yes | No | Can't tell | No | Yes | Yes | No | Yes | Moderate |
| Dancy, 2014 | Yes | Yes | No | Can't tell | No | Yes | Yes | Not applicable | Some concern | Moderate |
| Abamecha, 2021 | Yes | Yes | No | Can't tell | No | Yes | Yes | Not applicable | Yes | Moderate |
| Menna, 2015 | Yes | Yes | No | Can't tell | No | Yes | Yes | Yes | Some concern | Moderate |
| Edward, 2022 | Yes | Yes | Can't tell | Can't tell | No | Yes | Yes | Can't tell | Yes | High |
| Hassan, 2014 | Yes | Yes | No | Can't tell | No | Yes | Yes | Not applicable | No | Moderate |

**Note:**
**a** Following the JBI guidance, we classified studies pre-post studies as having no control group, since they lacked an independent and separate comparator. Studies using uncontrolled cross-sectional pre-post designs were also marked as having no control group, even when an independently sampled group was included. This is because we considered this weaker than an uncontrolled pre-post designs using the same participants. In addition, studies that did not compare outcomes between intervention and control groups in their results were classified as **not** having a control group.
**b** Similar to RCTs above, to assess baseline differences between treatment and control groups, we decided on the following criteria: (i) For binary or categorical variables: we used the threshold of a difference of $\pm 5$ percentage points or less as indicative of acceptable balance [101,102]; (ii) For continuous variables: we calculated the standardised difference using means and standard deviations of the characteristic variables for treatment and control groups and interpreted standardised differences that are no less than 0.1 as non-negligible imbalances [101,103]. This approach avoids reliance on p-values, which are sensitive to sample size and not recommended for assessing baseline balance in RCTs [104,105].
**c** For whether comparison groups received similar care apart from the intervention, we marked "Can't tell" if we didn't see any explicit discussion.
**d** We marked "Yes" only if the study had multiple measurements both pre- and post-intervention. We marked "No" if there were multiple measurements on only one side of the intervention (either pre- or post-intervention) but only a single measurement on the other side.
**e** For this question, we considered two aspects: the validity of the survey instrument and the training and reliability of data collectors. If a study addressed only the validity of the survey instrument but did not mention training of data collectors, we marked it as "Partial Yes." Notably, no study explicitly reported inter-rater reliability between data collectors (in cases of double data entry). However, we marked "Yes" if both aspects, namely the instrument validity and data collector training, were clearly documented.
**f** For follow-up, we marked “Not applicable” for studies with independently sampled post-intervention groups. We applied a 10% loss-to-follow-up threshold as a rule of thumb. Studies exceeding this threshold were rated “No” for this criterion. We placed particular emphasis on the actual extent of loss to follow-up: even if sensitivity analyses were conducted to account for missing data, studies were still marked “No” if loss to follow-up exceeded 10%.
**g** We marked “No” for appropriate statistical analysis used if effect sizes, confidence intervals and p-values were not comprehensively reported. Studies were also penalised if they had pre-intervention (baseline) data but did not use it to improve validity and transparency without sufficient justification.
**h** Our assessment of risk of bias (Low, Moderate, or High) was based on the percentage of “Yes” responses across the nine questions. Items marked as “Not applicable” were excluded from the denominator. Studies with at least 70% “Yes” answers were classified as Low risk of bias. Those with 50–70% “Yes” answers were considered to have a Moderate risk, while studies with fewer than 50% “Yes” answers were rated as High risk of bias. This approach provided a standardised method for categorising all RCTs and reflected our relatively strict treatment of “Can’t tell” responses, which were regarded unfavourably. We recommend that future studies provide clearer reporting on these aspects.
**i** Since only the Nigeria setting in Krug et al. (2023) met our comparator and methodological criteria, we included effect sizes exclusively from this setting. Accordingly, the risk of bias assessment pertains only to the Nigeria analysis, which employed a controlled before–after design with repeated cross-sectional data.

**Table H.1.3.** Risk of bias analysis for cross-sectional studies using the JBI tool.

| Study (First author’s last name, Year) | Were the criteria for inclusion in the sample clearly defined? | Were the study subjects and the setting described in detail? | Was the exposure measured in a valid and reliable way? | Were objective, standard criteria used for measurement of the condition? | Were confounding factors identified?<sup>a</sup> | Were strategies to deal with confounding factors stated?<sup>b</sup> | Were the outcomes measured in a valid and reliable way? | Was appropriate statistical analysis used?<sup>c</sup> | Risk of bias<sup>d</sup> |
| --- | --- | --- | --- | --- | --- | --- | --- | --- | --- |
| Brooks, 2019 | Yes | Yes | No | No | Yes | Yes | Yes | Yes | Low |

**Note:**
**a** Our agreed approach for this question was to mark “Yes” if the authors implemented efforts to account for covariates in the regression analysis and reported adjusted effect sizes. We acknowledge that residual confounding may still be present; thus, a “Yes” does not imply the study is free from endogeneity issues.
**b** We marked “Yes” if a study applied econometric techniques addressing confounding to some extent, such as multivariate logistic regression or propensity score matching.
**c** We marked “No” for appropriate statistical analysis if effect sizes, confidence intervals and p-values were not comprehensively reported.
**d** Our assessment of risk of bias (Low, Moderate, or High) was based on the percentage of “Yes” responses across the eight questions. Items marked as “Not applicable”, if any, were excluded from the denominator. Studies with at least 70% “Yes” answers were classified as Low risk of bias. Those with 50–70% “Yes” answers were considered to have a Moderate risk, while studies with fewer than 50% “Yes” answers were rated as High risk of bias. This approach provided a standardised method for categorising all RCTs and reflected our relatively strict treatment of “Can’t tell” responses, which were regarded unfavourably. We recommend that future studies provide clearer reporting on these aspects.

#### Appendix H.2 Publication Bias for Seven Key Outcomes

**Figure H.2.1.**
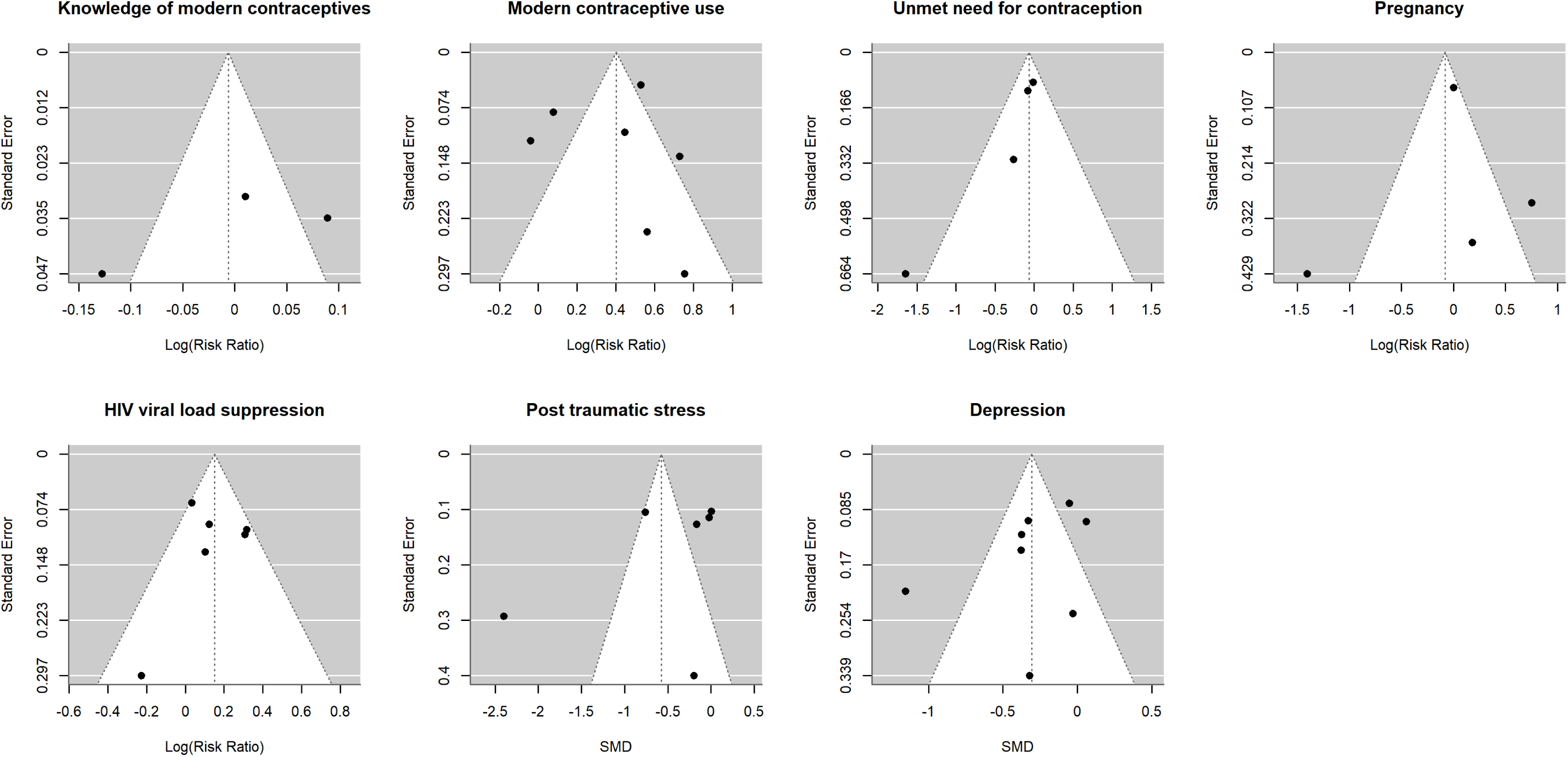
Funnel plots for Seven Key Outcomes Assessed in Meta-Analyses

#### Appendix H.3 GRADE Evaluation for Certainty of Cumulative Evidence for Seven Key Outcomes

**Table H.3.1.** Summary of GRADE Evidence Profile for Eight Key Outcomes.

| Outcome | No. of effect sizes<sup>a</sup> | Direction<sup>b</sup> | Risk of Bias | Indirectness | Inconsistency | Imprecision | Publication bias | Upgrading consideration | Certainty of evidence |
| --- | --- | --- | --- | --- | --- | --- | --- | --- | --- |
| HIV viral load suppression | 6 + 2 = 8 | Positive: 3;<br>Null: 5 | Serious concern<br>Downgrade (-1) | No concern | No serious concern | Serious concern;<br>Downgrade (-1) | Uncertain | Not upgrade | ⊕⊕○○<br>Low |
| Knowledge of modern contraception | 3 + 4 = 7 | Positive: 3;<br>Null: 4 | Serious concern<br>Downgrade (-1) | No concern | Serious concern;<br>Downgrade (-1) | Serious concern;<br>Downgrade (-1) | Uncertain | Not upgrade | ⊕○○○<br>Very Low |
| Use of modern contraceptive (including condom) | 7 + 8 = 15 | Positive: 8;<br>Null: 7 | Serious concern<br>Downgrade (-1) | No concern | Serious concern;<br>Downgrade (-1) | Some concern;<br>Downgrade (-1) | Some concern;<br>Downgrade (-1) | Not upgrade | ⊕○○○<br>Very Low |
| Unmet need for modern contraception | 4 + 0 = 4 | Positive: 1;<br>Null: 3 | Serious concern<br>Downgrade (-1) | No concern | No serious concern | Serious concern;<br>Downgrade (-1) | Uncertain | Not upgrade | ⊕○○○<br>Very Low |
| Adolescent pregnancy | 4 + 3 = 7 | Positive: 1;<br>Negative: 1;<br>Null: 5 | Serious concern<br>Downgrade (-1) | No concern | Serious concern;<br>Downgrade (-2) | Serious concern;<br>Downgrade (-1) | Uncertain | Not upgrade | ⊕○○○<br>Very Low |
| PTS | 6 + 3 = 9 | Positive: 4;<br>Null: 5 | Serious concern<br>Downgrade (-1) | No concern | Borderline decision | Serious concern;<br>Downgrade (-1) | Uncertain | Not upgrade | ⊕⊕○○<br>Low |
| Depression | 8 + 3 = 11 | Positive: 6;<br>Null: 5 | Serious concern<br>Downgrade (-1) | No concern | No serious concern | Some concern;<br>Downgrade (-1) | No serious concern | Not upgrade | ⊕⊕○○<br>Low |

**Note:**
**a** The first number refers to programmes included in the meta-analysis, while the second refers to the remaining effect sizes excluded from the meta-analysis. The count of effect sizes reflects intervention settings rather than individual studies. For example, Krug et al. (2023) [15] included two eligible settings in Nigeria, which were counted separately. Similarly, Dorsey et al. (2020) [16] included two disparate settings in Kenya and Tanzania that were counted as separate programmes.
**b** Direction here includes all relevant findings from the studies in this review. That is, even if a study did not report a specific effect size, we included it here the direction of the effect was indicated. The numbers in the column refer to the number of effect sizes corresponding to each direction.

To ensure transparency and stimulate academic discussion, we accompanied our decision on the quality of evidence in Table H.3.1 above with written documentation below.

### (1) HIV viral load suppression

We began the GRADE assessment at high certainty, as all studies included in the meta-analysis are RCTs.

#### Risk of bias

All six RCTs included in the meta-analysis were assessed as having a moderate risk of bias using JBI RCT checklist [17], with “Yes” responses ranging from 50–70%. Concerns primarily arose in domains such as baseline comparability^3^, randomisation masking, and loss-to-follow-up. Consequently, we downgraded the certainty of evidence by one level to reflect these serious concerns.

#### Indirectness

We assessed the PICO for all studies reporting HIV viral load suppression for indirectness^4^. Four studies focused on adolescents and young adults living with HIV, with age ranges extending up to 24 years, though the primary target group was adolescents aged 10–19 [18–21]. One study targeted caregiver of children and young adolescents aged 6-15 [18], while the remaining one examined exclusively adolescents aged 13-19 [22]. Although they focused on those with HIV or those with both HIV and mental health conditions, these subgroups fell well within the scope of our review.

The interventions primarily provided HIV treatment support though various approaches (see the “*Inconsistency*” subsection below for more details). Most studies used standard care or no intervention as comparators, reflecting real-world alternatives. One study used the Zvandiri programme as the control condition, which is the same programme served as the intervention in other studies, while its intervention group received Zvandiri programme combined with problem-solving therapy (PST)-based psychosocial support [20]. HIV viral load suppression was assessed using thresholds of either 400 or 1000 copies/mL across studies.

Given the overall alignment with our prespecified PICO criteria, we judged the body of evidence to be direct and therefore did not downgrade for indirectness.

#### Inconsistency

We observed the I² statistic was 45.86% with p-value being 0.084, indicating moderate variability across study results. Upon closer examination of the target populations, interventions, and comparators, this heterogeneity appears to be explainable. While most studies used either 400 or 1000 copies/mL as the viral load cutoff, variation exists across studies in terms of population targeted, types of interventions, and comparator conditions.

Specifically, for *target population*, for instance, one study targeted caregiver of younger children and adolescents aged 6-15 years old [18], and another on adolescents aged 13-19 [22]; both reported improvements in viral load suppression. In contrast, studies targeting broader age groups of adolescents and young adults up to 24 years old, despite also having median ages within 10-19, showed null effects. There are *differences in terms of participants’ baseline clinical characteristic*, even among studies of the Zvandiri programme. For example, Ndhlovu et al. (2021) included a higher-risk population with 86.3% had viral loads ≥1000 copies/mL [19], compared to 45%-49% in Mavhu et al. (2020) [22].

Regarding *interventions*, although several studies shared components, there was meaningful variation. Some interventions targeted caregivers [18–20,22,23], while others incorporated mental health or psychosocial elements [19,20,22,23], differentiated care approaches [21,22], or enhanced reminders for treatment adherence [19,20,22]. Even within the same Zvandiri programme, one study implemented a more comprehensive version [22], while another used a lighter version [19].

*Comparator conditions* also differed. Simms et al. (2022) used Zvandiri programme as control condition and added PST delivered by peer CATS to the intervention group [20]. In contrast, Mavhu et al. (2020) compared the Zvandiri programme to standard HIV care [22]. Thus, Simms et al. (2022) evaluated the added value of PST on top of Zvandiri, rather than Zvandiri itself [20]. Notably, both the Zvandiri-only and Zvandiri + PST arms in Simms et al. (2022) showed substantial reductions in virologically non-suppression, suggesting that the null effect pertained to the added PST component [20].

*Implementation factors* may also have contributed to heterogeneity. While all studies were RCTs, variations in execution could affect power and results. For example, Mavhu et al. (2020) had a larger sample size and strong follow-up, likely yielding more statistical power compared to Ndhlovu et al. (2021) [19,22].

Despite these potential sources of heterogeneity, the direction of effect sizes was largely consistent. Except for Simms et al. (2022) [20], all studies showed effect estimates in the positive direction, although not all were statistically significant. Furthermore, the confidence intervals across studies overlapped substantially. Therefore, given that the observed heterogeneity appears to be explainable and that confidence intervals overlapped, we judged the inconsistency as not serious and thus did not downgrade for inconsistency.

#### Imprecision

Due to data limitations, it is very hard to calculate the Optimal Information Size (OIS) directly, but given the large total sample size across studies (N=2,298), it is reasonable to assume that it exceeds the threshold of a single adequately powered RCT. For example, Mavhu et al. (2020) maintained that their RCT was already well-powered with a sample size of 496 [22]. The 95% confidence interval of the pooled relative risk, 0.99-1.36, included the null effect (Risk Ratio (RR) =1) and crossed the appreciable beneficial effect (say around 1.25). We decided to downgrade by one level to account for potential imprecision.

#### Publication bias

Visual inspection of the funnel plot suggested possible asymmetry as reflected by the lack of small-sample studies that reported negative results on HIV viral load suppression. While this may be an indication of publication bias, the small number of included studies (n=6) suggests that the evidence is not conclusive. We decided not to downgrade but flag this to be uncertain.

#### Upgrading Considerations

We found no compelling reasons to upgrade the quality of evidence. There was no evidence of a dose-response gradient. Moreover, there was unlikely uncontrolled residual confounding that might lead to underestimation of effects enough to justify an upgrade, particularly when all studies were RCTs. The effect sizes observed were mostly modest and close to the null, which ruled out a large effect. Thus, no upgrades were applied.

#### Overall

We downgraded the certainty of evidence by two levels from high certainty due to serious concerns related to risk of bias and imprecision. The final quality of evidence is **Low**.

### (2) Knowledge of modern contraceptives

Three effect sizes were included in the meta-analysis, derived from two studies: one NRSI and one RCT. As the NRSI contributed most participants, we initiated the certainty of evidence rating at low.

#### Risk of bias

Of the two studies included in the meta-analysis, the two Nigerian settings reported in Krug et al. (2023) [15] were assessed as having a moderate risk of bias using the JBI checklist for NRSIs [24]. Austrian et al. (2021) [25] was judged to have a moderate risk of bias according to the JBI checklist for RCTs [17].

We considered downgrading the certainty of evidence because the majority of the sample size was contributed by a study with design-related critical limitations [15], which, according to the GRADE handbook, may warrant automatic downgrading for study design concerns [26]. In addition, the RCT [25] was assessed as having a moderate risk of bias. Taken together, we downgraded the certainty of evidence by (at least) one level to reflect serious concerns regarding risk of bias.

#### Indirectness

We assessed the PICO elements of the two studies examining knowledge of modern contraceptives to evaluate indirectness. In terms of *population*, studies focused on adolescent girls aged 15–19 years or those in grade seven. Although these represent specific subgroups, they remain well within our target population. Interventions were delivered by lay-adult CHWs, and primarily involved group education, aligning closely with our intervention criteria. Comparators consisted of no-intervention control groups. Outcomes were consistently measured as awareness of modern contraceptives. Given this close alignment with our review objectives, we did not downgrade the certainty of evidence for indirectness.

#### Inconsistency

We judged the evidence for knowledge of modern contraceptives to have serious concerns regarding inconsistency and downgraded the certainty of evidence by one level. Visual inspection of the forest plot (Figure 5a) indicated limited overlap of confidence intervals, suggesting variability in effect estimates beyond what would be expected by chance alone. This was supported by statistical indicators of heterogeneity (I² = 88.16%, Q-test p < 0.001). However, the estimated between-study variance was small (τ^2^ ≈ 0.01), indicating that the absolute magnitude of heterogeneity was limited, and we acknowledge that, given the small number of studies (k = 3), I² may be inflated and should be interpreted with caution.

Nevertheless, there was variability in the magnitude of effect estimates across studies^5^, with some estimates suggesting positive effects and others being closer to the null, alongside limited overlap of confidence intervals. Taken together, this indicates meaningful inconsistency in effect estimates, and we therefore downgraded the certainty of evidence by one level.

#### Imprecision

Due to data limitations, it is very hard to calculate the OIS directly, but given the large total sample size across studies (N=10,878), it is reasonable to assume that it can exceed the threshold of a single adequately powered RCT. The 95% confidence interval of the pooled relative risk ratio, 0.76-1.30, includes both the null effect (RR=1) and the appreciable benefit thresholds (roughly RR larger than 1.25 or smaller than 0.75). We decided to downgrade by one level for imprecision.

#### Publication bias

Visual inspection of the funnel plot did not suggest clear asymmetry, with studies appearing relatively evenly distributed around the pooled estimate. While this may indicate a low risk of publication bias, the small number of included studies (n=3) limits the ability to draw firm conclusions. We therefore do not downgrade but note that the evidence remains uncertain.

#### Upgrading Considerations

We found no compelling reasons to upgrade the quality of evidence. There was no evidence of a dose-response gradient or residual confounding left unattended that might underestimate the effect enough to justify an upgrade. Observed effect sizes were mostly modest and close to null, ruling out a strong effect. Thus, no upgrades were applied.

#### Overall

Considering the serious concerns regarding risk of bias, inconsistency, and imprecision, the final quality of evidence was rated from low to **Very Low**.

### (3) Use of modern contraceptives

Despite the presence of two RCTs, most of the studies, which contributed the largest sample sizes, were NRSIs, plus one cross-sectional study. We therefore began our evaluation of the quality of evidence from low.

#### Risk of bias

Among the seven intervention settings, Silverman et al. (2023) [27] and Wondimagegene et al. (2023) [28] were assessed as having a moderate risk of bias using JBI checklist for RCTs [17]. Krug et al. (2023) (Nigeria settings) [15], Menna et al. (2015) [29], and Dancy et al. (2014) [30] were rated as having a moderate risk of bias using JBI checklist for NRSIs [24]. Brooks et al. (2019) [31] was evaluated as having a low risk of bias using the JBI checklist for analytical cross-sectional studies [32].

We considered downgrading the certainty of evidence, as the majority of the sample size was derived from studies with study designs subject to important limitations [15,29–31], which, according to the GRADE handbook, may warrant automatic downgrading for study design-related concerns [26]. In addition, the two RCTs [27,28] were assessed as having a moderate risk of bias. Taken together, we downgraded the certainty of evidence by (at least) one level to reflect serious concerns regarding risk of bias in results for this outcome.

#### Indirectness

Most studies examining modern contraceptive use focused on adolescent girls aged 13-19 years, with some specifically targeting married [15,27,31] or in-school [28,29] girls. Interventions were more commonly delivered by lay-peer CHWs and included household visits, education sessions, counselling and service provision. Comparators comprised individuals not exposed to the intervention or without CHW contact. Outcomes assessed included current use of modern contraceptives and consistent condom use. On this basis, we did not downgrade for indirectness.

#### Inconsistency

We assessed the evidence on unmet need for modern contraceptives as raising some concerns regarding consistency. First, Figure 5b in the main report shows that several confidence intervals do not overlap, suggesting variation in study results beyond what would be expected by chance. This concern is reinforced by the high level of heterogeneity (I^2^ = 87.83%, p < 0.001, τ^2^ ≈ 0.075), indicating substantial inconsistency across studies. In addition, the vote-counting analysis revealed a mixed pattern of findings: of the 15 total effect sizes, eight were significantly positive while seven were null. Taken together, these findings suggest notable inconsistency, leading us to downgrade the certainty of the evidence by one level for serious concerns.

#### Imprecision

Due to data limitations, it is very hard to calculate the OIS directly, but given the large total sample size across studies (N=15,834), it is reasonable to assume that it exceeds the threshold of a single adequately powered RCT. The 95% confidence interval of the pooled relative risk ratio spanned from 1.13 to 1.99, which did not include the no effect (RR=1) but included the appreciable benefit threshold (RR larger than 1.25), which led us to consider downgrading for quality by one level to be conservative.

#### Publication bias

Visual inspection of the funnel plot suggested some asymmetry, with a greater concentration of studies on one side of the pooled estimate and a relative absence of small-sample studies reporting null or negative results. While this may indicate potential publication bias or small-study effects, the limited number of studies (n=7) and possible heterogeneity across interventions and settings mean the evidence is not conclusive. To be conservative, we downgraded by one level to reflect potential risks.

#### Upgrading considerations

We found no compelling evidence to upgrade the quality. There was no evidence of a dose-response gradient or residual confounding left unattended that might underestimate the effect enough to justify an upgrade. Observed effect sizes from five studies were quite large [27–31]. However, the small estimates from Krug et al. (2023) appeared to dominate the overall effect given their huge sample size, making the pooled effect estimate not sufficiently large to justify an upgrade. Thus, no upgrades were applied.

#### Overall

We decided to downgrade the quality of evidence to be **Very Low**, considering risk of bias, serious concern regarding inconsistency, and some concerns over impreciseness and publication bias.

### (4) Unmet need for modern contraceptives

Although two of the three studies were clustered RCTs, two of the four effect sizes pooled came from a non-randomised study of interventions with a dominant sample size, employing controlled before-after with repeated cross-sectional design. Therefore, we began the evaluation with a low certainty.

#### Risk of bias

Among the four effect sizes, Wondimagegene et al. (2023) [28] and Hensen et al. (2023) [33] were assessed as having a moderate risk of bias using the JBI checklist for RCTs [17]. Krug et al. (2023) [15] was also judged to have a moderate risk of bias, based on the JBI checklist for NRSIs [24].

We considered downgrading the certainty of evidence because the majority of the sample size was contributed by studies with designs subject to critical limitations [15], which, according to the GRADE handbook, may warrant automatic downgrading for study design-related concerns [26]. In addition, the included RCTs [28,33] were themselves assessed as having moderate risk of bias. Taken together, we downgraded the certainty of evidence by (at least) one level to reflect serious concerns regarding internal validity.

#### Indirectness

All studies targeted adolescent girls aged between 15-19 years old, with some focusing specifically on married or in-school girls. The interventions were predominantly led by lay-peer CHWs. Intervention activities included education sessions, household visits, and facilitation of service access. Comparators comprised individuals not exposed to the intervention. Outcomes were consistently defined as the proportion of sexually active girls who did not wish to become pregnant, intended to use contraceptives, but failed to do so. On this basis, we did not downgrade for indirectness.

#### Inconsistency

We identified minimal concerns regarding the consistency of evidence for this outcome. The I^2^ was 0.01% with a Q-test p-value of 0.091 and τ^2^ ≈ 0, indicating negligible heterogeneity. Confidence intervals across studies showed substantial overlap, except for Wondimagegene et al. (2023) [28]. Only one [28] out of the four effect sizes indicated reduction in unmet need for modern contraceptives, while the remaining three [15,33] saw no effects. We decided not to downgrade for inconsistency.

#### Imprecision

Due to data limitations, it is very hard to calculate the OIS directly, but given the large total sample size across studies (N=16,915), it is reasonable to assume that it exceeds the threshold of a single adequately powered RCT. The 95% confidence interval of the pooled relative risk ratio spanned from 0.68 to 1.29, including both the no effect (RR=1) and the appreciable benefit and harm thresholds. Therefore, we considered downgrading by one level to reflect concerns regarding imprecision.

#### Publication bias

Visual inspection of the funnel plot suggested possible asymmetry, with an uneven distribution of studies and a lack of smaller studies on one side of the funnel. This pattern may indicate potential publication bias. However, the small number of included studies (n=4) limits interpretability. We do not downgrade but note the uncertainty.

#### Upgrading Considerations

We found no compelling reasons to upgrade the quality of evidence. There was no evidence of a dose-response gradient or residual confounding left unattended that might underestimate the effect enough to justify an upgrade. Observed effect sizes were mostly close to null, ruling out a strong effect. Thus, no upgrades were applied.

#### Overall

We decided to downgrade the certainty of evidence by one level due to risk of bias and imprecision. The final level was then **Very Low.**

### (5) Adolescent pregnancy

We started the evaluation of certainty of evidence with low level since 3/4 effect estimates came from observational studies.

#### Risk of bias

Of the four intervention settings, Rudgard et al. (2022) [34] was assessed as having a low risk of bias while the two Nigeria settings in Krug et al. (2023) [15] were found to have a moderate risk of bias, using the JBI checklist for NRSIs [24]. Hensen et al. (2023) [33] was considered as having a moderate risk of bias using the JBI checklist for RCTs [17]. We considered downgrading the certainty of evidence because the majority of the sample size came from studies whose designs had critical limitations [15,34], which, according to the GRADE handbook, may warrant automatic downgrading for design-related concerns [26]. Moreover, the RCT study [33] had a moderate risk of bias. Taken together, we downgraded the certainty of evidence by (at least) one level to reflect concerns regarding internal validity.

#### Indirectness

We assessed PICO for indirectness for all studies that examined this outcome and found no concerns. All interventions targeted adolescent girls aged between 15 and 19 years old. The interventions primarily involved household visits, group education, and service access, led mostly by lay-adult CHWs [15,34]. Comparators included individuals with no exposure to interventions. Outcomes were generally measured by birth within the last 12 months [15,33], except Rudgard et al. (2022), which assessed the absence of pregnancy by age 19 years old [34]. Given the strong alignment with our review objectives, we did not downgrade for indirectness.

#### Inconsistency

We identified very serious concerns regarding inconsistency for this outcome and downgraded the certainty of evidence by two levels. Statistical heterogeneity was very substantial (I² = 90.64%, p < 0.001; τ² ≈ 0.64), indicating considerable variability in effect estimates across studies. Unlike other outcomes (e.g., modern contraceptive use or HIV viral load suppression), this heterogeneity was not readily explained by differences in study design or intervention characteristics. For instance, in the two Nigeria settings reported by Krug et al. (2023), intervention with highly similar components and delivery mechanisms yielded markedly different results, with one showing no effect and the other indicating a substantial increase in adolescent pregnancy [15].

Furthermore, there was little to no overlap in confidence intervals across several studies (e.g., Rudgard et al. (2022) [34] versus Krug et al. (2023) [15] and Hensen et al. (2023) [33]), and both the direction and magnitude of effects varied widely across settings. Taken together, the large magnitude of heterogeneity, lack of clear explanation, and substantial variability in effect estimates indicate very serious inconsistency, warranting downgrading the certainty of evidence by two levels.

#### Imprecision

Due to data limitations, it is very hard to calculate the OIS directly, but given the large total sample size across studies (N=38,310), it is reasonable to assume that it can exceed the threshold of a single adequately powered RCT. The 95% confidence interval of the pooled relative risk ratio is very wide, spanning from 0.23 to 3.77. This included both no effect (RR=1) and appreciable harm and benefit thresholds (e.g., RR = 0.75 and 1.25 respectively). We considered downgrading by at least one level to reflect the concerns in imprecision.

*Publication bias*Given the presence of small studies reporting both positive and negative results, and the limited number of effect estimates (n=4), we did not downgrade for publication bias but note a moderate level of uncertainty.

#### Upgrading considerations

We found no compelling reasons to upgrade the quality of evidence. There was no evidence of a dose-response gradient or unaddressed residual confounding likely to underestimate the effect sufficiently to justify an upgrade. The observed effect sizes also did not support upgrading: although Rudgard et al. (2022) reported a large positive effect on reducing pregnancy [34], the other interventions showed either no effect [15,33] or large increases in pregnancy [15]. Thus, no upgrades were applied.

#### Overall

We decided to downgrade for some concerns about risk of bias, inconsistency, and imprecision. The final certainty of evidence is **Very Low**.

### (6) Post-traumatic stress (PTS)

We began our evaluation of the certainty of evidence from a high level, as all six effect sizes in the meta-analysis were derived from RCTs.

#### Risk of bias

The five RCTs included in the meta-analysis were assessed as having a moderate risk of bias [16,23,35–37] using the JBI checklist for RCTs [17]. Overall, we downgraded the certainty of evidence by one level, reflecting some serious concerns about internal validity.

#### Indirectness

We assessed the PICO for indirectness across all studies that examined PTS and found no concerns. All studies targeted children and/or adolescents with some trauma, including war-affected trauma, orphanhood-related, HIV-related, and even various types of traumatic experiences. The age range of the participants was from 5 to 24 overall for this outcome, but most of the participants were adolescents aged 10-19 years old. Interventions were predominantly adapted from CBT and mostly delivered by lay-adult CHWs. Comparators were those that received standard of care. Outcomes were mostly measured using child-reported PTSD symptom scale, as we believe this can better reflects children’s and adolescents’ internal states than guardian-reported PTS outcomes. Given such relevancy, we did not downgrade for indirectness for PTS outcome.

#### Inconsistency

We identified some concerns regarding inconsistency and flagged this as a borderline issue. The I^2^ statistic was 97.55% with a Q-test p-value of < 0.001 and τ^2^ ≈ 0.77, indicating very large heterogeneity across study results. Almost all interventions were led by lay-adult CHWs and were primarily group-based, adapted from CBT, with a focus on improving adolescents’ coping skills in response to traumatic experiences.

However, such observed heterogeneity appears to be at least partially explainable by several factors. One key source of variation is the *baseline severity of mental health symptoms*. For instance, in Dorsey et al. (2020) [16] Tanzanian participants exhibited better baseline mental health than their Kenyan counterparts and saw limited improvement. The *nature of the trauma* also likely contributed to variability in outcomes. For example, younger children and adolescents in Murray et al. (2015) were coping with acute trauma and loss [37], which may be more responsive to targeted therapy. In contrast, Dow et al. (2020) examined adolescents experiencing chronic, HIV-related stress [23], conditions that may lead to more entrenched and less modifiable mental health symptoms. Moreover, trauma stemming from HIV may carry more severe societal stigma than that associated with orphanhood, potentially affecting intervention effectiveness. *Other factors, such as age differences across the target populations,* may also have influenced outcomes. We acknowledged that these explanations were speculative and not conclusive. Nevertheless, they appeared to offer plausible reasons for the high level of heterogeneity observed.

While one study had confidence interval that did not overlap with others and showed particularly large positive effects in reducing PTSD symptoms [37], the remaining studies demonstrated similar effect sizes with overlapping confidence intervals. Importantly, no study reported negative effects on adolescent PTS outcomes.

In summary, while we recognised substantial heterogeneity, the consistency in the direction and general magnitude of the effects, alongside plausible explanations for result heterogeneity, led us to judge this as a borderline case. We opted not to downgrade for inconsistency but highlight the potential for variability in effects on PTSD symptoms.

#### Imprecision

We found minimal concerns regarding imprecision. The total sample size across the meta-analysis was 1,767, which is relatively large, although it is not possible to definitively assess whether this exceeds the OIS for a fully powered trial. The pooled effect estimate −0.58 is of moderate magnitude, but the 95% confidence interval, which is −1.54-0.38, includes both the null effect and clinically important effect. Accordingly, we downgraded by one level for imprecision for PTS.

#### Publication bias

Visual inspection of the funnel plot suggests some asymmetry, with a relative paucity of small studies reporting negative or null effects. This may indicate potential publication bias or small-study effects. However, given the limited number of included studies (n=6) and potential heterogeneity in study characteristics, this finding should be interpreted with caution. We do not downgrade but flag this as uncertain.

#### Upgrading considerations

We found no compelling justification to upgrade the quality of evidence. There was no evidence of a dose-response relationship, nor any indication of uncontrolled substantial residual confounding that led to an underestimation of the effect particularly given that all included studies were RCTs. On the contrary, certain sources of bias, such as social desirability bias, might have contributed to overestimation of effects. Overall, the observed effect sizes were mostly modest or small, except for two studies reporting very large effects [16,37]. While we did not give an upgrade to be conservative, we acknowledged that some studies demonstrated meaningful and clinically relevant effects.

#### Overall

Considering all above, we decided to downgrade by one level due to some serious concerns in the risk-of-bias assessment and serious concern regarding imprecision. The final certainty of evidence is **Low**.

### (7) Depression

Seven out of eight effect sizes were derived from RCTs, with the remaining effect size from a NRSI that had a relatively small sample size [39]. We therefore started the evaluation with high certainty of evidence.

#### Risk of bias

Seven out of eight effect sizes were derived from RCTs which were assessed as having a moderate risk of bias using the JBI checklist for RCTs [17]. The remaining effect size came from a NRSI [39] rated as having a moderate risk of bias using JBI checklist for NRSI [24]. Overall, we downgraded the certainty of evidence by one level, reflecting some serious concerns over internal validity.

#### Indirectness

Like PTS, we found no concern over indirectness for adolescent depression. Studies mostly targeted (children and) adolescents with some types and degrees of trauma experience, including war-affected trauma, orphanhood and grief-related, HIV-related, motherhood-associated, disruptive behaviour, and mental health disorders. The age range of the participants was from 7 to 24 overall for this outcome, but most of the participants were adolescents aged 10-19 years old. A large proportion of the interventions took the form of adapted versions of CBT delivered by lay-adult CHWs. Comparators were those that received standard of care. Depression was measured using a wider range of indicators, although the most common one is PHQ-9 and youth CBCL. We included those reported by children or adolescents in our meta-analysis as we believe that their own report may better reflect their true internal states, while vote-counting analysis included all effect sizes reported by either children or their guardian. Given such relevancy, we did not downgrade for indirectness for adolescent depression outcome.

#### Inconsistency

We found no serious concerns over the inconsistency. Although the I^2^ was 85.93% with Q-test p value less than 0.001 and τ^2^ **≈** 0.1, which suggested substantial heterogeneity across study results, the differences in the study results could potentially be explained by various factors.

First, the results could be explained by *types of trauma and problems they face*. It can be the case that orphanhood-aroused trauma and children’s disruptive behaviour more likely elicit empathy while HIV-related are highly stigmatised, leading to greater social isolation, which might affect the intervention effectiveness. Results might also differ based on *how intervention was delivered*. Targeting similar highly stigmatised issues, such as HIV or adolescent motherhood, Dow et al. (2020) found no effects [23] while Tinago et al. (2024) found large reductions in adolescent mothers’ depression [39]. This might suggest the importance of the intervention design and the community engagement component. Tinago et al. (2024) incorporated strong community engagement and trust-building efforts [39], which may have fostered a more supportive environment for adolescent mothers. Dow et al. (2020)’s intervention didn’t appear to address external stigma directly [23], and HIV-related stigma remained deeply ingrained, potentially limiting the effectiveness of CHW-led interventions alone. Moreover, peer-led interventions appear to be quite effective in dealing with trauma with potential high social stigma, such as Simms et al. (2022) [20] and Tinago et al. (2024) [39].

*Baseline conditions* also matter to explain the discrepancies between results. For instance, Dorsey et al. (2020) demonstrated that intervention effectiveness varied by context [16]. Their study found stronger effects in urban Kenya compared to rural Kenya [16]. One possible explanation is that urban participants experienced more severe trauma-related events and reported worse food security outcomes suggesting poverty dynamics at play. In urban Kenya, 67.2% of participants reported going to bed hungry at least some of the time in the past 14 days. In rural Kenya, this figure was even higher at 88.3% [16]. However, in Tanzania, only 10.9% (urban) and 5.5% (rural) reported food insecurity. This suggests that in rural Tanzania, where basic needs were relatively better met, there was less room for mental health improvement through the intervention [16]. Another example is that in Tinago et al. (2024), qualitative evidence suggested that for many participants, the intervention was their first opportunity to openly discuss SRHR issues [39]. This initial exposure to SRHR discussions may have led to large immediate effects, though longer follow-ups are needed to determine whether the effects are sustained.

Although not conclusive, the above discussion might suggest that heterogeneity in results was potentially explainable. Moreover, we found the confidence intervals to be quite overlapping with each other, and nine out of eleven studies in the meta-analysis pointed to a negative effect.

Overall, we did not downgrade for inconsistency, considering that the heterogeneity in results appeared to be explainable by different potentially reasonable factors, and effect sizes were largely similar in direction and magnitude.

#### Imprecision

The 95% confidence interval for the summary effect estimate of CHW intervention’s effects on adolescent depression, (−0.61,0), included the null effect (marginally) and the appreciable beneficial effect. We considered downgrading by one level for imprecision, despite that the OIS criterion should largely be met given the total number of participants of studies included in the meta-analysis was 2,580.

#### Publication bias

Although some variation appeared to exist in the effect estimates from relatively larger studies, the effect estimates still quite consistently pointed to a beneficial effect of the task-shifting strategy to CHWs. The consistency of positive effects in large trials, combined with the symmetry of the funnel plot around the pooled effect, tended to suggest a lower degree of publication bias. We therefore did not downgrade.

#### Upgrading Considerations

We found no compelling reasons to upgrade the quality of evidence. There was neither evidence of a dose-response gradient, nor an indication of substantive residual confounding that potentially led to underestimation of the effects if uncontrolled particularly when most studies were RCTs. On the contrary, some sources of bias (e.g. social desirability bias) might have led to overestimation. The effect sizes observed were mostly small and modest, which ruled out a strong or large effect. Thus, no upgrades were applied.

#### Overall

Given the serious concerns raised over risk of bias and imprecision, we decided to downgrade the certainty of evidence by two levels. So, the final quality of evidence was **Low.**

### Appendix I: Summary of Vote-Counting Results for All Outcomes

**Table I.1.** Number of Positive/Negative/Null Effect Sizes by Outcome ^a^.

| Outcome Category | Number of effect sizes | Number of positive effect sizes | Number of negative effect sizes | Number of no effect |
| --- | --- | --- | --- | --- |
| **Knowledge, Attitudes and Perceptions** |  |  |  |  |
| ***HIV-related*** |  |  |  |  |
| HIV knowledge | 7 | 5 | 0 | 2 |
| Attitude towards HIV | 3 | 2 | 0 | 1 |
| Knowledge of one's HIV status | 1 | 1 | 0 | 0 |
| Attitude towards HIV-related services (e.g., HIV voluntary testing) | 1 | 0 | 0 | 1 |
| Perception of spread of HIV | 1 | 1 | 0 | 0 |
| Perception of likelihood that people change their SRH behaviour to stop the spread of HIV | 1 | 1 | 0 | 0 |
| ***SRHR-related*** |  |  |  |  |
| Knowledge of modern contraceptive methods/condom use | 7 | 3 | 0 | 4 |
| Attitude towards modern contraceptives/condom use | 6 | 2 | 1 | 3 |
| Knowledge of STIs | 4 | 2 | 0 | 2 |
| Knowledge of pregnancy | 1 | 0 | 0 | 1 |
| Knowledge of fertility | 3 | 0 | 0 | 3 |
| Knowledge of SRHR-related services | 2 | 0 | 0 | 2 |
| Attitude towards menstrual health | 1 | 1 | 0 | 0 |
| ***Malaria*** |  |  |  |  |
| Knowledge of malaria | 1 | 1 | 0 | 0 |
| Attitude towards malaria | 1 | 1 | 0 | 0 |
| Perception of malaria | 2 | 1 | 1 | 0 |
| **Health Behaviour and Outcomes** |  |  |  |  |
| ***HIV-related*** |  |  |  |  |
| HIV viral suppression | 8 | 3 | 0 | 5 |
| HIV testing services uptake | 5 | 4 | 0 | 1 |
| ART adherence | 7 | 2 | 0 | 5 |
| Linkage to and retention in care | 6 | 0 | 0 | 6 |
| Patient satisfaction | 5 | 2 | 0 | 3 |
| Other HIV-related services uptake <sup>b</sup> | 5 | 1 | 0 | 4 |
| Participation in community HIV prevention activities | 1 | 1 | 0 | 0 |
| ***SRHR-related*** |  |  |  |  |
| Use and prevalence rate of modern contraceptive methods or condom use | 15 | 8 | 0 | 7 |
| Adolescent pregnancy | 7 | 1 | 1 | 5 |
| Unmet need for contraceptives | 4 | 1 | 0 | 3 |
| Self-efficacy in sexual relations | 4 | 2 | 0 | 2 |
| Coverage of SRH services (a broad concept) | 3 | 3 | 0 | 0 |
| Number of sexual partners | 2 | 0 | 0 | 2 |
| Demand for contraception | 1 | 1 | 0 | 0 |
| Sexual intercourse initiation | 1 | 0 | 0 | 1 |
| SRH practices (general concept) <sup>c</sup> | 1 | 0 | 0 | 0 |
| Partner communication | 1 | 1 | 0 | 0 |
| Abstained in the last 12 months | 1 | 0 | 0 | 1 |
| Use of an appropriate menstrual products at last menstruation or “has enough pads” (self-reported) | 2 | 2 | 0 | 0 |
| Met material needs at last menstruation among AGYM | 4 | 0 | 0 | 4 |
| Reporting leaking menstrual blood at school | 1 | 1 | 0 | 0 |
| ***Mental health*** |  |  |  |  |
| Gender mental disorder and health | 5 | 2 | 0 | 3 |
| Trauma- and stress-related symptoms | 9 | 4 | 0 | 5 |
| Depression | 11 | 6 | 0 | 5 |
| Anxiety | 1 | 1 | 0 | 0 |
| Self-concept | 1 | 1 | 0 | 0 |
| Unresolved grief symptoms/prolonged grief | 2 | 1 | 0 | 1 |
| Functional impairment | 5 | 3 | 0 | 2 |
| Emotional and behavioural symptoms and difficulties | 5 | 0 | 0 | 5 |
| **Maternal health** |  |  |  |  |
| ANC visit | 2 | 2 | 0 | 0 |
| Continuum of care | 2 | 2 | 0 | 0 |
| Skilled birth attendance | 2 | 0 | 0 | 2 |
| Post-natal care within 24 hours of delivery | 2 | 1 | 0 | 1 |
| **Malaria** |  |  |  |  |
| Use of insecticide-treated bed net | 1 | 1 | 0 | 0 |
| **Nutrition** |  |  |  |  |
| Dietary diversity | 1 | 1 | 0 | 0 |
| **Behavioural adjustment** |  |  |  |  |
| Prosocial behaviour | 1 | 1 | 0 | 0 |
| Behavioural disorder | 2 | 1 | 0 | 1 |
| **Alcohol use** |  |  |  |  |
| Alcohol use | 1 | 0 | 0 | 1 |
| **Other outcomes (general health measures)** |  |  |  |  |
| General health status | 2 | 0 | 0 | 2 |
| Health utility | 1 | 1 | 0 | 0 |
| All-cause mortality | 2 | 0 | 0 | 2 |
| Anthropometric outcome | 2 | 0 | 0 | 2 |
| Physical quality of life | 1 | 0 | 0 | 1 |
| Quality of life | 1 | 0 | 0 | 1 |
| **Well-being** |  |  |  |  |
| **Safe and supportive environment** |  |  |  |  |
| Intimate partner violence | 2 | 0 | 0 | 2 |
| Child labour | 2 | 2 | 0 | 0 |
| Child marriage | 1 | 1 | 0 | 0 |
| Positive peer, family and community support | 10 | 6 | 0 | 4 |
| Family functioning | 1 | 1 | 0 | 0 |
| Gender norms in marriage | 1 | 0 | 0 | 1 |
| Equitable adolescent gender norms | 1 | 0 | 0 | 1 |
| Gendered sexual norms | 1 | 1 | 0 | 0 |
| **Learning, education and employment** |  |  |  |  |
| School attendance, completion and retention | 8 | 4 | 0 | 4 |
| Literacy, numeracy and academic performance | 7 | 6 | 0 | 1 |
| Labour/Employment and earnings | 4 | 1 | 0 | 3 |
| **Agency and resilience** |  |  |  |  |
| Self-efficacy | 3 | 2 | 0 | 1 |
| Self-esteem | 1 | 1 | 0 | 0 |
| Hope <sup>d</sup> | 1 | 0 | 0 | 1 |
| **Connectedness** |  |  |  |  |
| Healthy relationship | 7 | 3 | 0 | 4 |

**Note:**

### Appendix J. Sensitivity Analysis of Meta-Analysis

**Table J.1.** HIV Viral Load Suppression.

| Analysis | k | Effect Size | 95% CI | CI Method | I<sup>2</sup> | $\tau^2$ | Q-test p-value | Method |
| --- | --- | --- | --- | --- | --- | --- | --- | --- |
| **Main analysis** | 6 | 1.16 | (0.99, 1.36)<sup>3</sup> | HKSJ | 45.86% | 0.009 | 0.084 | REML |
| **Alternative estimation method** | 6 | 1.16 | (0.99, 1.36) | HKSJ | 48.45% | 0.01 | 0.084 | DL |
| **Alternative CI method – z** | 6 | 1.16 | (1.03, 1.31) | z-based Wald-type | 45.86% | 0.009 | 0.084 | REML |
| **Alternative CI method – t** | 6 | 1.16 | (1.00, 1.36)<sup>4</sup> | t-based Wald-type | 45.86% | 0.009 | 0.084 | REML |
| **Excluding Simms** | 5 | 1.18 | (1.00, 1.39)<sup>6</sup> | HKSJ | 50.65% | 0.009 | 0.086 | REML |
| **Excluding Simms** | 5 | 1.18 | (1.05, 1.33) | z-based Wald-type | 50.65% | 0.009 | 0.086 | REML |

Note:
1. Effect sizes are expressed as RR. Between-study variance was estimated using restricted maximum likelihood (REML), with DerSimonian–Laird (DL) used as an alternative moment-based estimator. Column with header k represents the number of effect sizes pooled in the meta-analysis.
2. Please note that all included studies are RCTs, so there is no separate row for “Including uncontrolled pre-post”.
3. p value = 0.0570
4. p value = 0.0543
5. The “Excluding Simms” rows present results after excluding Simms et al. (2022) [20]. This sensitivity analysis was conducted because the control group in Simms et al. received an active intervention (Zvandiri) that is comparable to the intervention conditions in other included studies. As such, the estimated effect in this study reflects the incremental impact of adding problem-solving therapy (PST), rather than the effect of Zvandiri programme relative to standard care. Although both intervention and control groups in Simms et al. demonstrated substantial improvements in viral load suppression, the additional effect of PST was not statistically significant. We first applied the HKSJ CI and additionally examined z-based Wald-type CI, consistent with guidance from the Cochrane Handbook [41], which notes that HKSJ may yield overly conservative CIs in settings where all studies suggest beneficial effects. Under both approaches, the pooled estimate after excluding Simms et al. was positive and statistically significant.
6. p=0.0475
7. The sensitivity analysis excluding Simms et al. (2022) resulted in a statistically significant pooled effect as in Note 5 above, whereas the summary effect did approximately reach conventional significance under Wald-type CI (row 3 & 4) but not under HKSJ inferential method (row 1 or 2). This suggests that the overall conclusion is somewhat sensitive to study inclusion and the choice of inferential method. However, the direction and magnitude of the effect were consistent across analyses, indicating that the findings are broadly stable but subject to some uncertainty in statistical significance.

**Table J.2.**
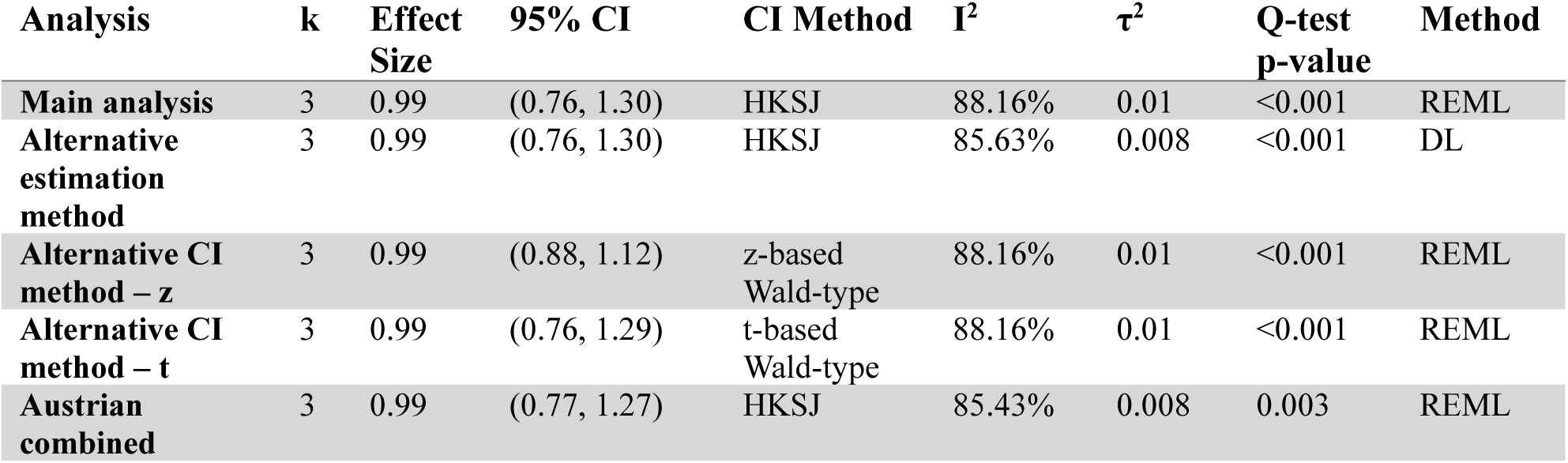
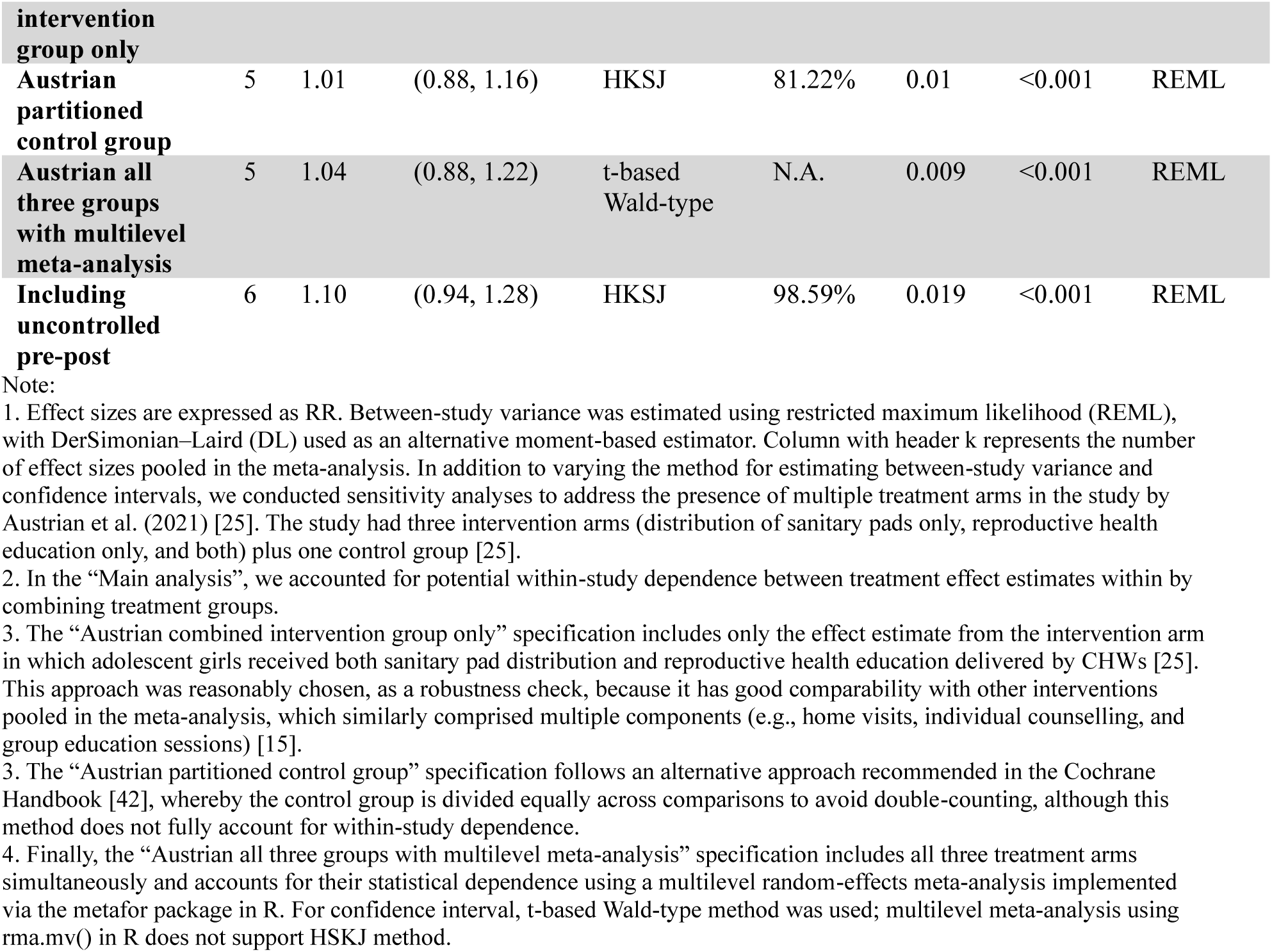
Knowledge of Modern Contraceptives.

**Table J.3.** Use of Modern Contraceptives.

| Analysis | k | Effect Size | 95% CI | CI Method | I<sup>2</sup> | τ<sup>2</sup> | Q-test p-value | Method |
| --- | --- | --- | --- | --- | --- | --- | --- | --- |
| **Main analysis** | 7 | 1.50 | (1.13, 1.99) | HKSJ | 87.83% | 0.075 | <0.001 | REML |
| **Alternative estimation method** | 7 | 1.49 | (1.13, 1.98) | HKSJ | 86.99% | 0.070 | <0.001 | DL |
| **Alternative CI method – z** | 7 | 1.50 | (1.19, 1.88) | z-based Wald-type | 87.83% | 0.075 | <0.001 | REML |
| **Alternative CI method – t** | 7 | 1.50 | (1.12, 2.00) | t-based Wald-type | 87.83% | 0.075 | <0.001 | REML |
| **Including uncontrolled pre-post** | 9 | 1.34 | (1.02, 1.74) | HKSJ | 95.25% | 0.102 | <0.001 | REML |

Note:
1. Effect sizes are expressed as RR. Between-study variance was estimated using restricted maximum likelihood (REML), with DerSimonian–Laird (DL) used as an alternative moment-based estimator. Column with header k represents the number of effect sizes pooled in the meta-analysis.

**Table J.4.** Unmet Need for Modern Contraception.

| Analysis | k | Effect Size | 95% CI | CI Method | I<sup>2</sup> | $\tau^2$ | Q-test p-value | Method |
| --- | --- | --- | --- | --- | --- | --- | --- | --- |
| Main analysis | 4 | 0.94 | (0.68, 1.29) | HKSJ | 0.01% | 0 | 0.091 | REML |
| Alternative estimation method | 4 | 0.88 | (0.50, 1.54) | HKSJ | 53.53% | 0.031 | 0.091 | DL |
| Alternative CI method – z | 4 | 0.94 | (0.82, 1.07) | z-based Wald-type | 0.01% | 0 | 0.091 | REML |
| Alternative CI method – t | 4 | 0.94 | (0.75, 1.17) | t-based Wald-type | 0.01% | 0 | 0.091 | REML |
| Including uncontrolled pre-post | 6 | 0.94 | (0.82, 1.08) | HKSJ | 6.13% | 0.001 | 0.104 | REML |

Note:
1. Effect sizes are expressed as RR. Between-study variance was estimated using restricted maximum likelihood (REML), with DerSimonian–Laird (DL) used as an alternative moment-based estimator. Column with header k represents the number of effect sizes pooled in the meta-analysis.
2. The small number of studies, alongside the absence of large heterogeneity, indicates that HKSJ method may yield overly narrow confidence intervals. Even if it is the case, the result does not appear to change significantly.

**Table J.5.**
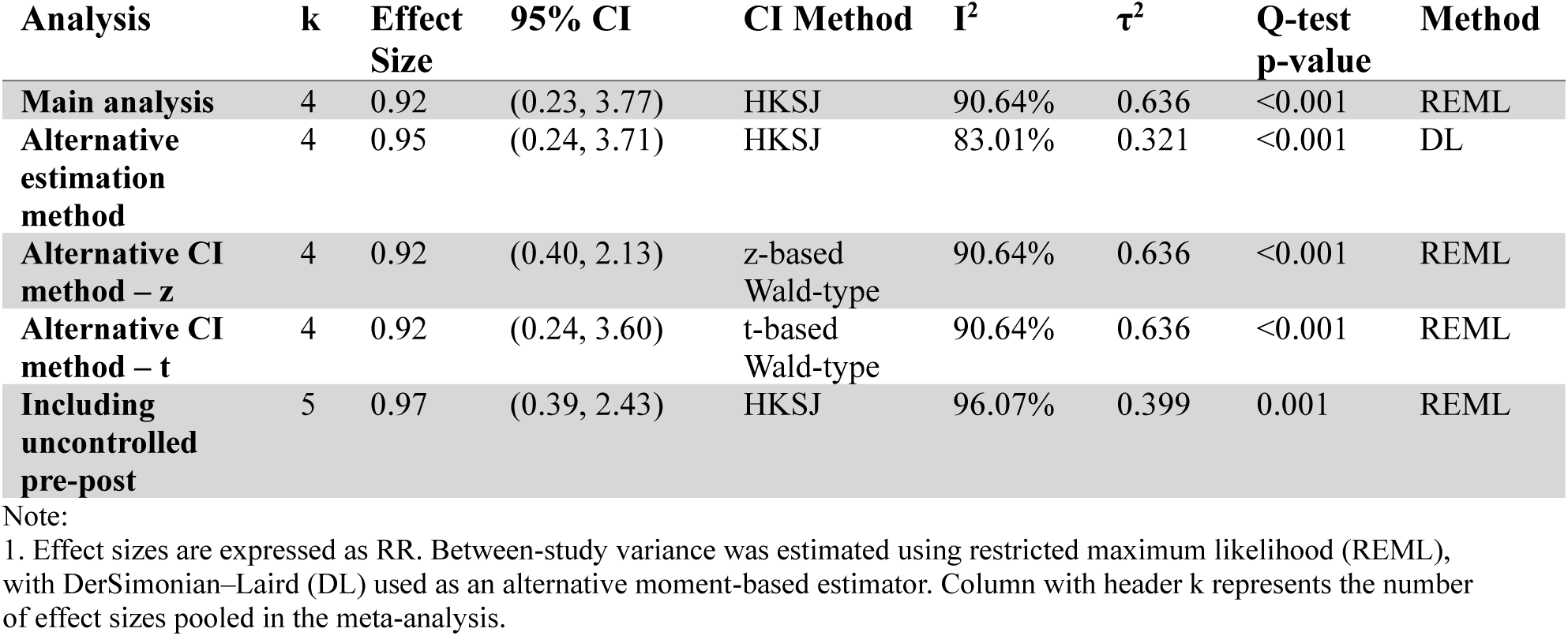
Adolescent Pregnancy.

**Table J.6.** PTS.

| Analysis | k | Effect Size | 95% CI | CI Method | I<sup>2</sup> | $\tau^2$ | Q-test p-value | Method |
| --- | --- | --- | --- | --- | --- | --- | --- | --- |
| Main analysis | 6 | -0.58 | (-1.54, 0.38) | HKSJ | 97.55% | 0.766 | <0.001 | REML |
| Alternative estimation method | 6 | -0.55 | (-1.49, 0.38) | HKSJ | 94.18% | 0.310 | <0.001 | DL |
| Alternative CI method – z | 6 | -0.58 | (-1.30, 0.15) | z-based Wald-type | 97.55% | 0.766 | <0.001 | REML |
| **Alternative CI method – t** | 6 | -0.58 | (-1.52, 0.37) | t-based Wald-type | 97.55% | 0.766 | <0.001 | REML |
| **Dorsey all four effect estimates included with multilevel meta-analysis** | 8 | -0.62 | (-1.85, 0.61) | HKSJ | 97.55% | 0.931 | <0.001 | REML |
| **Including uncontrolled pre-post** | 7 | -0.77 | (-1.67, 0.13) | HKSJ | 98.01% | 0.901 | <0.001 | REML |

Note:
1. Effect sizes are expressed as RR. Between-study variance was estimated using restricted maximum likelihood (REML), with DerSimonian–Laird (DL) used as an alternative moment-based estimator. Column with header k represents the number of effect sizes pooled in the meta-analysis.
2. In the “Main analysis”, we accounted for potential within-study dependence between urban and rural estimates within each country by combining these estimates at the country level. It is, however, reasonable to assume that population and effect estimate across countries are independent.
3. “Dorsey: all four effect estimates included with multilevel meta-analysis” presents results from a robustness check in which all four effect estimates (urban Kenya, rural Kenya, urban Tanzania, and rural Tanzania) were included, and within-study dependence was explicitly modelled using a multilevel meta-analysis.

**Table J.7.**
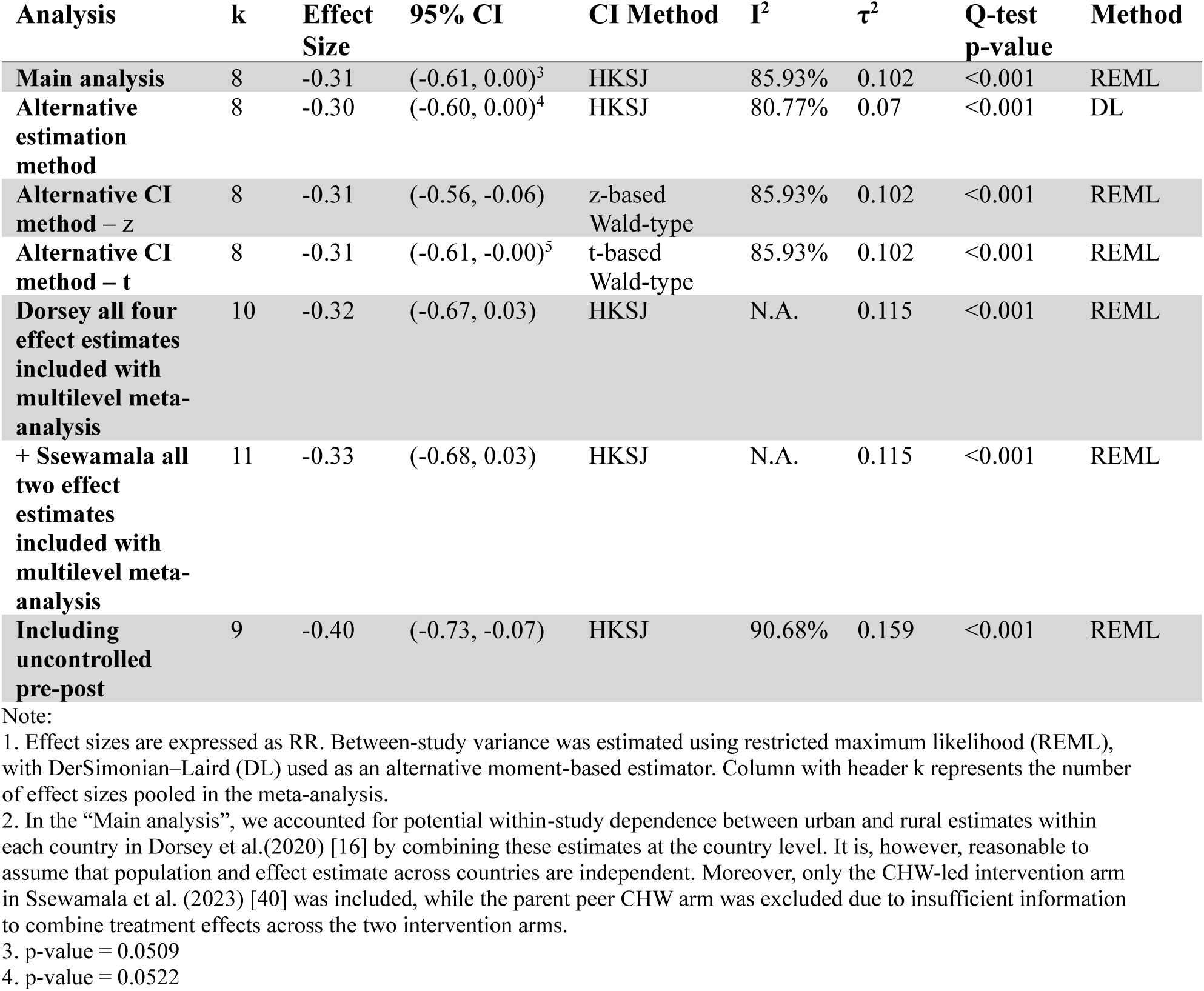
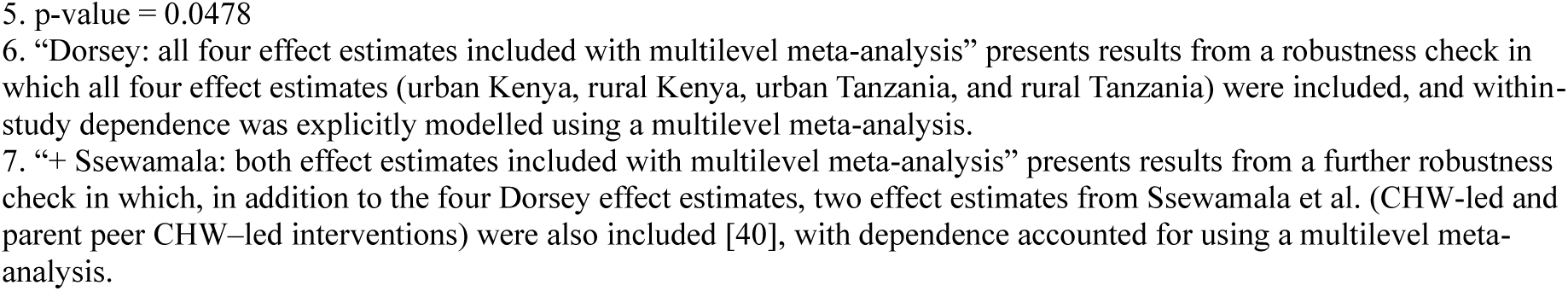
Depression.

| **Analysis** | **k** | **Effect Size** | **95% CI** | **CI Method** | **I<sup>2</sup>** | **τ<sup>2</sup>** | **Q-test p-value** | **Method** |
| --- | --- | --- | --- | --- | --- | --- | --- | --- |
| **Main analysis** | 8 | -0.31 | (-0.61, 0.00)<sup>3</sup> | HKSJ | 85.93% | 0.102 | <0.001 | REML |
| **Alternative estimation method** | 8 | -0.30 | (-0.60, 0.00)<sup>4</sup> | HKSJ | 80.77% | 0.07 | <0.001 | DL |
| **Alternative CI method – z** | 8 | -0.31 | (-0.56, -0.06) | z-based Wald-type | 85.93% | 0.102 | <0.001 | REML |
| **Alternative CI method – t** | 8 | -0.31 | (-0.61, -0.00)<sup>5</sup> | t-based Wald-type | 85.93% | 0.102 | <0.001 | REML |
| **Dorsey all four effect estimates included with multilevel meta-analysis** | 10 | -0.32 | (-0.67, 0.03) | HKSJ | N.A. | 0.115 | <0.001 | REML |
| **+ Ssewamala all two effect estimates included with multilevel meta-analysis** | 11 | -0.33 | (-0.68, 0.03) | HKSJ | N.A. | 0.115 | <0.001 | REML |
| **Including uncontrolled pre-post** | 9 | -0.40 | (-0.73, -0.07) | HKSJ | 90.68% | 0.159 | <0.001 | REML |

Note:
1. Effect sizes are expressed as RR. Between-study variance was estimated using restricted maximum likelihood (REML), with DerSimonian–Laird (DL) used as an alternative moment-based estimator. Column with header k represents the number of effect sizes pooled in the meta-analysis.
2. In the “Main analysis”, we accounted for potential within-study dependence between urban and rural estimates within each country in Dorsey et al.(2020) [16] by combining these estimates at the country level. It is, however, reasonable to assume that population and effect estimate across countries are independent. Moreover, only the CHW-led intervention arm in Ssewamala et al. (2023) [40] was included, while the parent peer CHW arm was excluded due to insufficient information to combine treatment effects across the two intervention arms.
3. p-value = 0.0509
4. p-value = 0.0522
5. p-value = 0.0478
6. “Dorsey: all four effect estimates included with multilevel meta-analysis” presents results from a robustness check in which all four effect estimates (urban Kenya, rural Kenya, urban Tanzania, and rural Tanzania) were included, and within-study dependence was explicitly modelled using a multilevel meta-analysis.
7. “+ Ssewamala: both effect estimates included with multilevel meta-analysis” presents results from a further robustness check in which, in addition to the four Dorsey effect estimates, two effect estimates from Ssewamala et al. (CHW-led and parent peer CHW-led interventions) were also included [40], with dependence accounted for using a multilevel meta-analysis.

**Table J.8.** Summary of Methods Used to Address Dependence.

| **Source of Dependence** | **Study (First Author, Year)** | **Methods in the Main Analysis** | **Methods in the Sensitivity Analysis** |
| --- | --- | --- | --- |
| Multi-arm study within a single population | Austrian, 2021 | Combine treatment groups where participants received sanitary pads and reproductive health education separately | (i) Partition control group<br>(ii) Include effect estimate from the treatment arm where participants received both sanitary pad and reproductive health education<br>(iii) Include all three effect estimates and use multilevel meta-analysis |
| Multi-arm study within a single population | Ssewamala, 2023 | Choose the CHW-led group and drop the parent peer group | (i) Include both effect estimates and use multilevel meta-analysis |
| Multi-settings with plausibly interconnected population | Dorsey, 2020 | Combine urban and rural settings using inverse-variance weighting, and include the weighted effect estimate | (i) Include all four estimates and use multilevel meta-analysis |

### Appendix K. Summary of CHW Effects for Well-Being Outcomes

### Safe and supportive environment

Evidence of the effect of CHW-led interventions on perceived support from peer, family, and community was mixed across different contexts. Across 10 effect sizes, six reported positive effects [35,39,43,44], while four were null [15,43]. Abamecha et al. (2021) found increased malaria-related family support following peer education and SBCC in Ethiopia [44]. However, mixed evidence was particularly salient regarding adolescents’ mental health. Betancourt et al. (2014) reported improved social support for adolescents with mental health needs through a group-based psychosocial intervention in Sierra Leone [35]. In Kenya, Osborn et al. (2020) observed gains in peer support but no family or significant-other support following lay-peer CHW-led education and mentoring [43]. Tinago et al. (2024) documented broad improvements in support from friends, family, and significant others among adolescent mothers participating in peer support sessions on SRHR, mental health, relationships, and livelihoods in Zimbabwe [39]. In contrast, Krug et al. (2023) found no improvements in community support for adolescent SRHR following household visits, counselling, and community engagement in Nigeria [15].

Evidence on intimate partner violence (IPV), examined in two studies, was consistently null (2/2 effect sizes). Austrian et al. (2021; RCT) found no effects of pad distribution, reproductive health education, or their combination on IPV acceptability in Kenya [25]^6^. Similarly, Silverman et al. (2023; RCT) reported no impact of the Reaching Married Adolescents programme in Niger, which included home visits, gender-synchronised group discussions, and community dialogues, on IPV outcomes, despite improvements in contraceptive use, highlighting the difficulty of shifting entrenched gender power dynamics [27].

All remaining outcomes were examined in single studies. For child marriage, Rudgard et al. (2022) found that exposure to HEW increased the likelihood of avoiding child marriage in Ethiopia [34]. For child labour, Posso et al. (2022) reported reductions in both extensive and intensive margins of child labour attributable to HEW exposure in Ethiopia, with differential benefits for boys (greater reductions in intensity) and girls (greater reductions in participation) [45]. For family functioning, Brathwaite et al. (2022) reported improvements in the CHW-led intervention group following an intervention designed to reduce disruptive behaviours through interactive group sessions in Uganda [46]. Regarding gender norms, Austrian et al. (2021; RCT) found that lay-adult CHW-led interventions improved gendered sexual norms, but did not yield significant improvements in equitable adolescent gender norms or marriage-related gender norms. Disaggregated results across treatment arms indicated improvements in equitable adolescent gender norms and gendered sexual norms in Kenya for both the reproductive health-only and combined intervention arms, but no significant effects on marriage-related gender norms [25].

### Learning, education and employment

Across 8 effect sizes on school attendance, completion, and retention, four were positive [34,35], with the remaining four reporting null effects all from Austrian et al. (2021) in Kenya using different school attendance and engagement measures [25]. These findings align with recent quantitative evidence [47] and suggest that structural constraints, such as poverty, school-related costs, gender disparities in educational valuation, and household instability [48], may outweigh menstrual health as primary determinants of school attendance. This supports a shift away from school attendance as a primary outcome in menstrual health programmes in certain contexts.

In contrast, outcomes related to literacy, numeracy, and academic performance showed more consistent improvements, with six of seven effect sizes positive [34,35,43,46]. In Ethiopia, Rudgard et al. (2022) found that HEW exposure improved literacy for both boys and girls and numeracy for girls [34]. Brathwaite et al. (2022) reported academic progress in CHW-led groups in Uganda [46]. Osborn et al. (2020) found improved average grades among Kenyan participants post-intervention [43], while Betancourt et al. (2014) documented sustained gains in academic performance eight months after programme completion in Sierra Leone [35].

Evidence on labour and earnings was limited to a single study. Rudgard et al. (2022) found a reduction in the proportion of boys engaged in more than four income-generating activities per day, but not corresponding effect for girls [34]. No effects were found on time spent in domestic activities (less than three hours per day) for either gender [34].

### Connectedness – Healthy relationships

Among the seven effect sizes assessing healthy relationships, three, reported in Dorsey et al. (2020) [16], showed improvements. Specifically, the study found increased guardian-reported relationship closeness in both Kenya and Tanzania, as well as reduced relationship conflict in Kenya, but no significant effect on relationship conflict in Tanzania [16]. In contrast, Brathwaite et al. (2022) found no evidence of improvement (at the 8 weeks, earliest measure in that study) in impaired relationships with peers, siblings, or parents following a lay-adult CHW-led, school-based group intervention that engaged adolescents and their caregivers to address behavioural problems and mental health issues in Uganda [46].

### Agency and resilience

Three effect sizes assessed general self-efficacy^7^, of which two exhibited improvements [25,44] and one was null [43]. Self-esteem and hope was assessed in a single study respectively: Brathwaite et al. (2022) reported improvements self-esteem in Uganda [46], and Tol found no effect on hope in Burundi [49].

### Appendix L. Figure 1 – 5 in the Main Text

**Figure 1.**
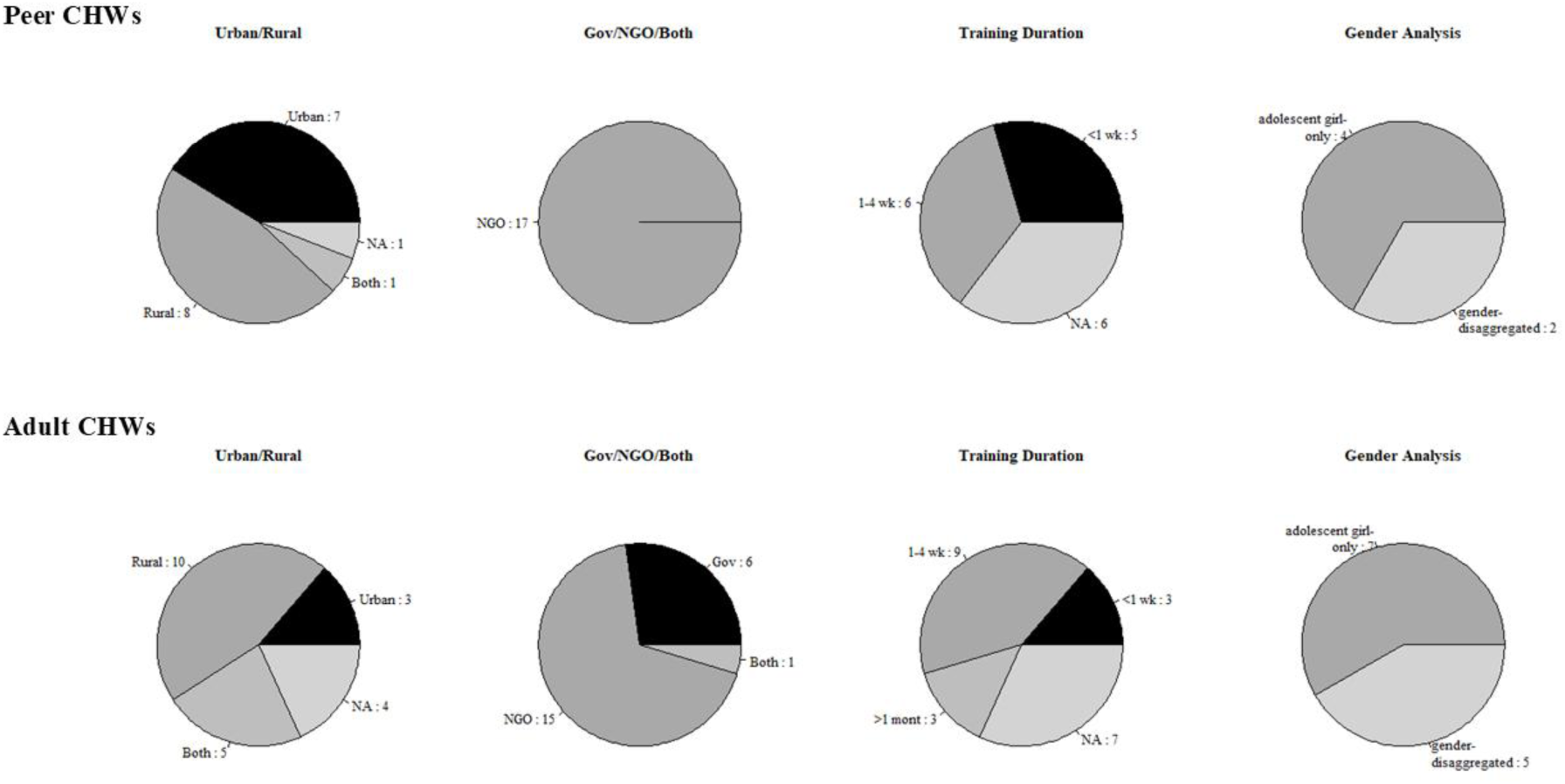
Programmatic design features of the evaluated CHW interventions.

**Figure 2.**
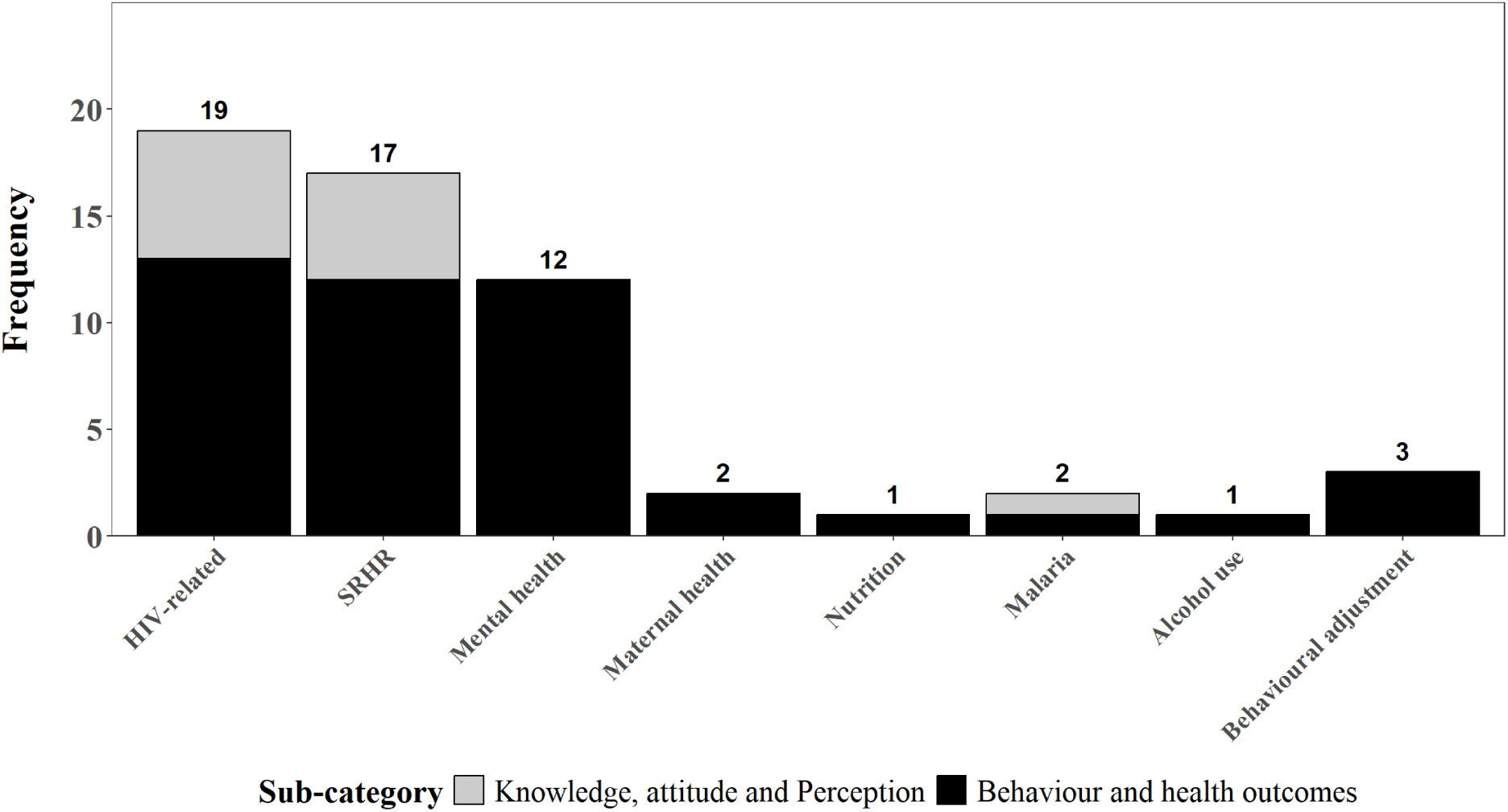
Outcome domains across CHW-led adolescent health interventions, by type.

**Figure 3.**
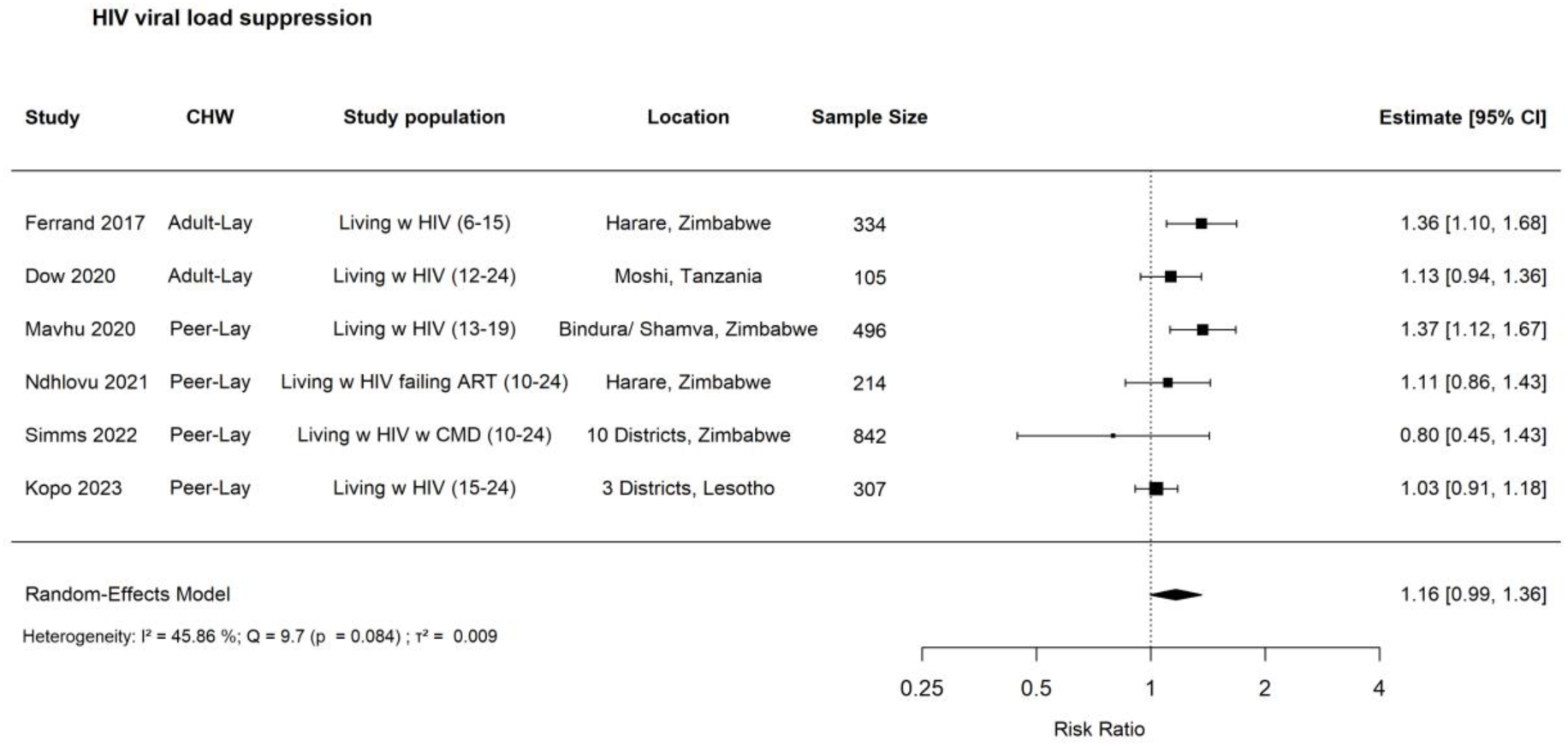
Forest plot: risk ratio for the effects of CHW-led interventions on HIV viral load suppression among adolescents in SSA.

**Figure 4.**
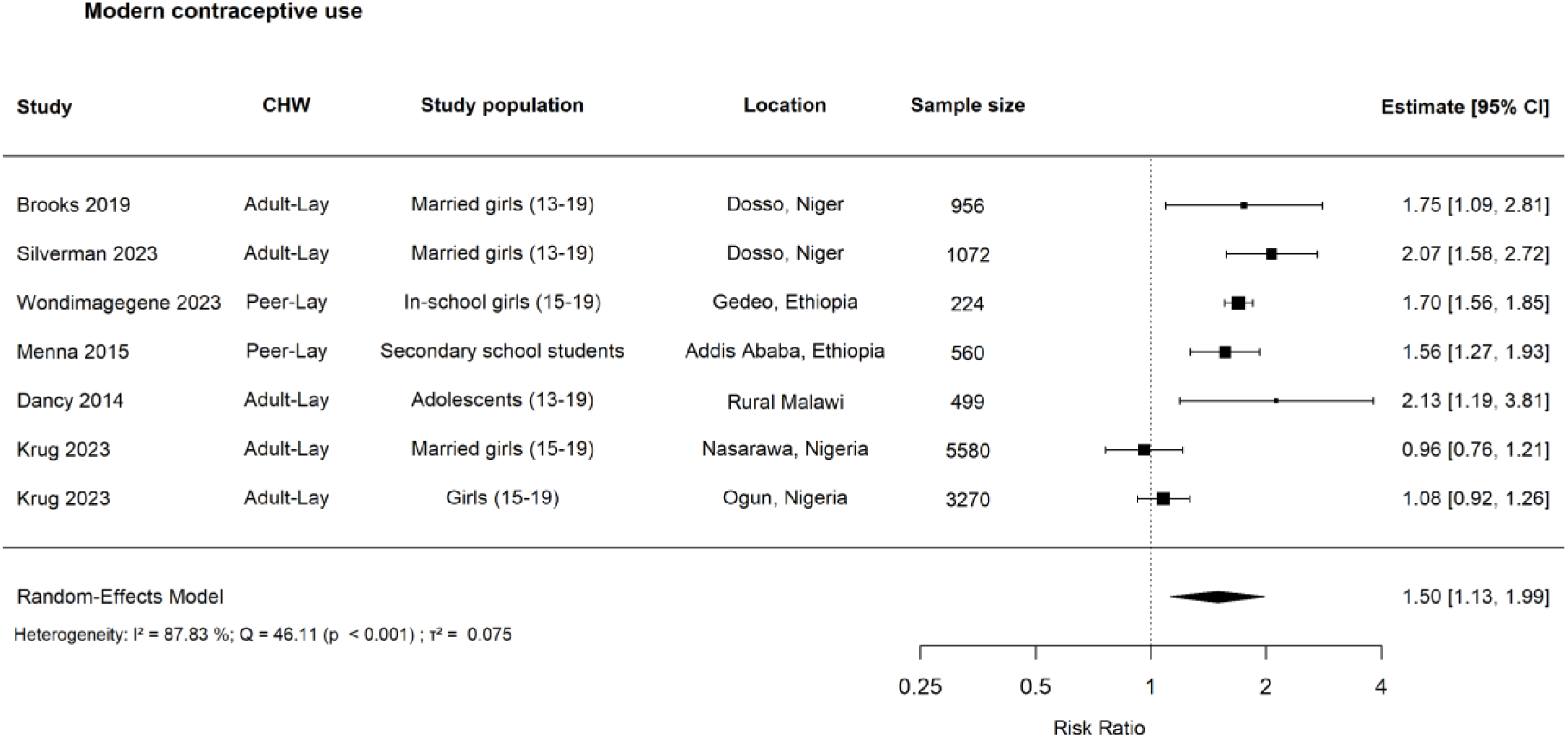
Forest plot: risk ratio for the effects of CHW-led interventions on modern contraceptive use among adolescents in SSA.

**Figure 5.**
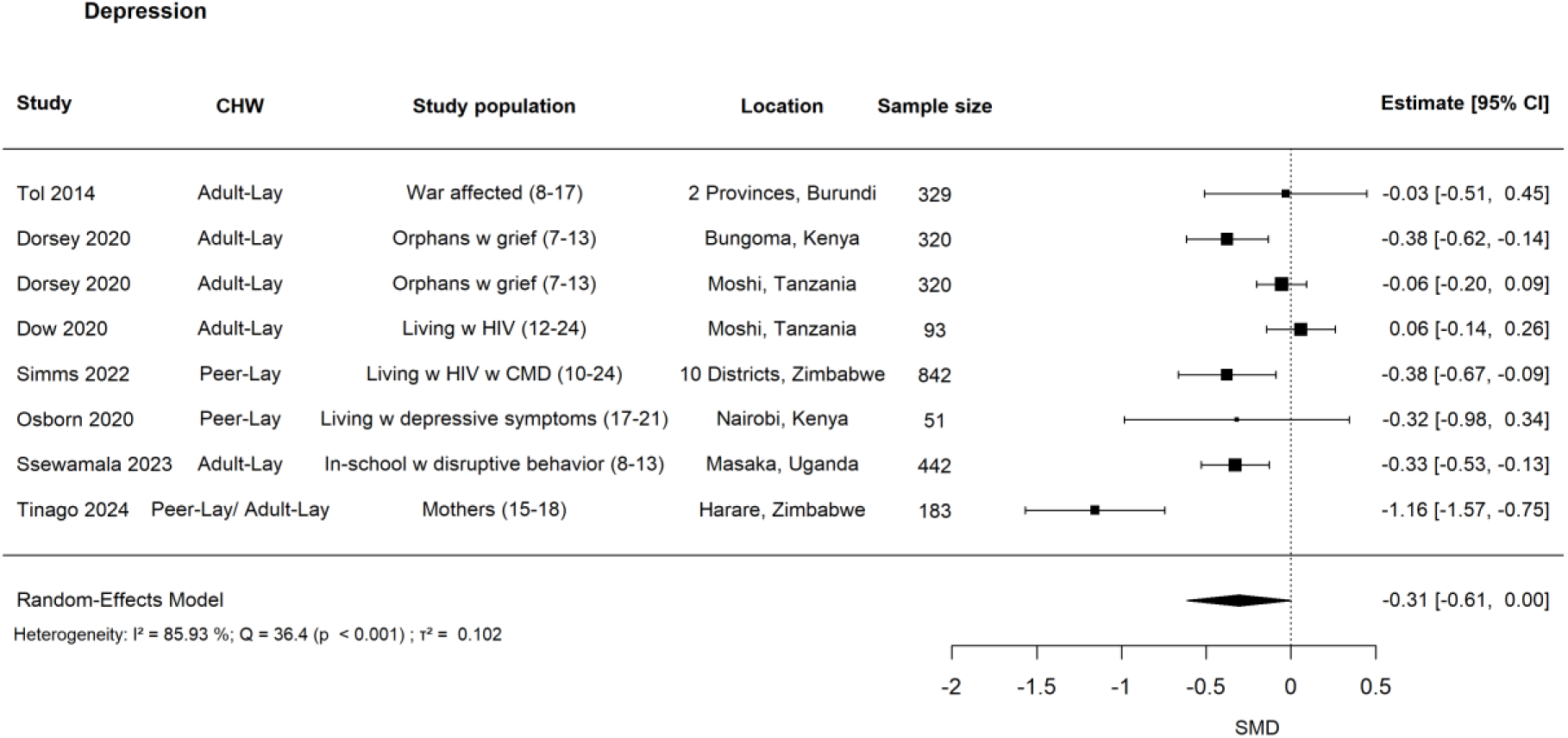
Forest plot: standardised mean difference (SMD) for the effects of CHW-led interventions on depression among adolescents in SSA.

### Appendix M. Additional Meta-Analysis Figures

**Figure M.1.**
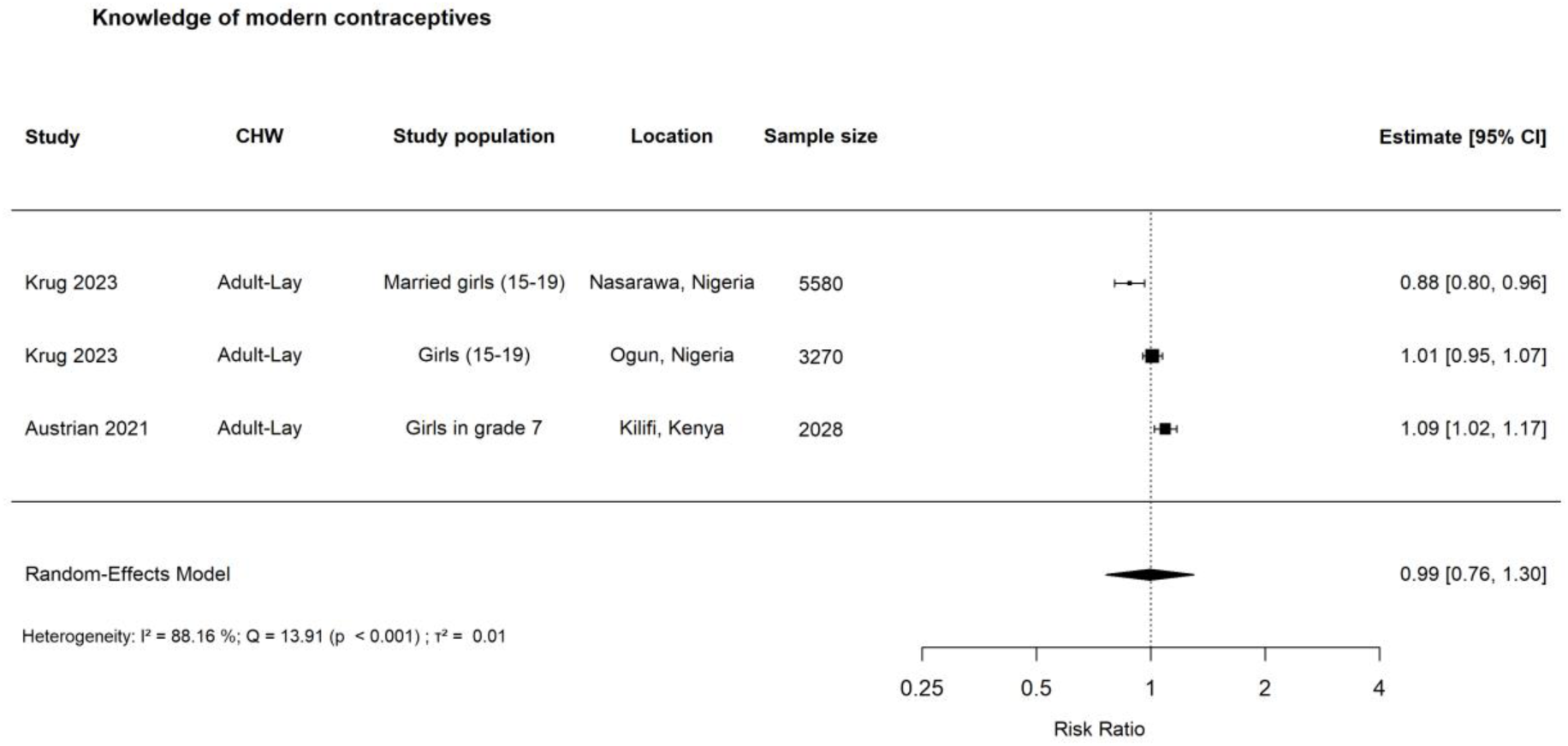
Forest plot: risk ratio for the effects of CHW-led interventions on knowledge of modern contraceptives among adolescents in SSA. **Note:** Three of nine effect sizes were included in the meta-analysis presented above. These measures capture indicators such as awareness of modern contraceptives and the ability to spontaneously name a modern contraceptive method. In the main analysis here, we combined the pad distribution and reproductive health education treatment arms in Austrian et al. (2023) [25], while alternative approaches to handling multiple treatment arms of the study are reported in Table J.2 in Appendix J. We excluded effect measures relating to knowledge of the benefits of modern contraception and intention to use a modern contraceptive method. Visual inspection of the funnel plot (see Appendix H.2) did not suggest clear asymmetry, but the small number of studies (n=3) limits the ability to draw firm conclusions. Details about publication bias, heterogeneity, and imprecision for this outcome can be found in Appendix H.

**Figure M.2.**
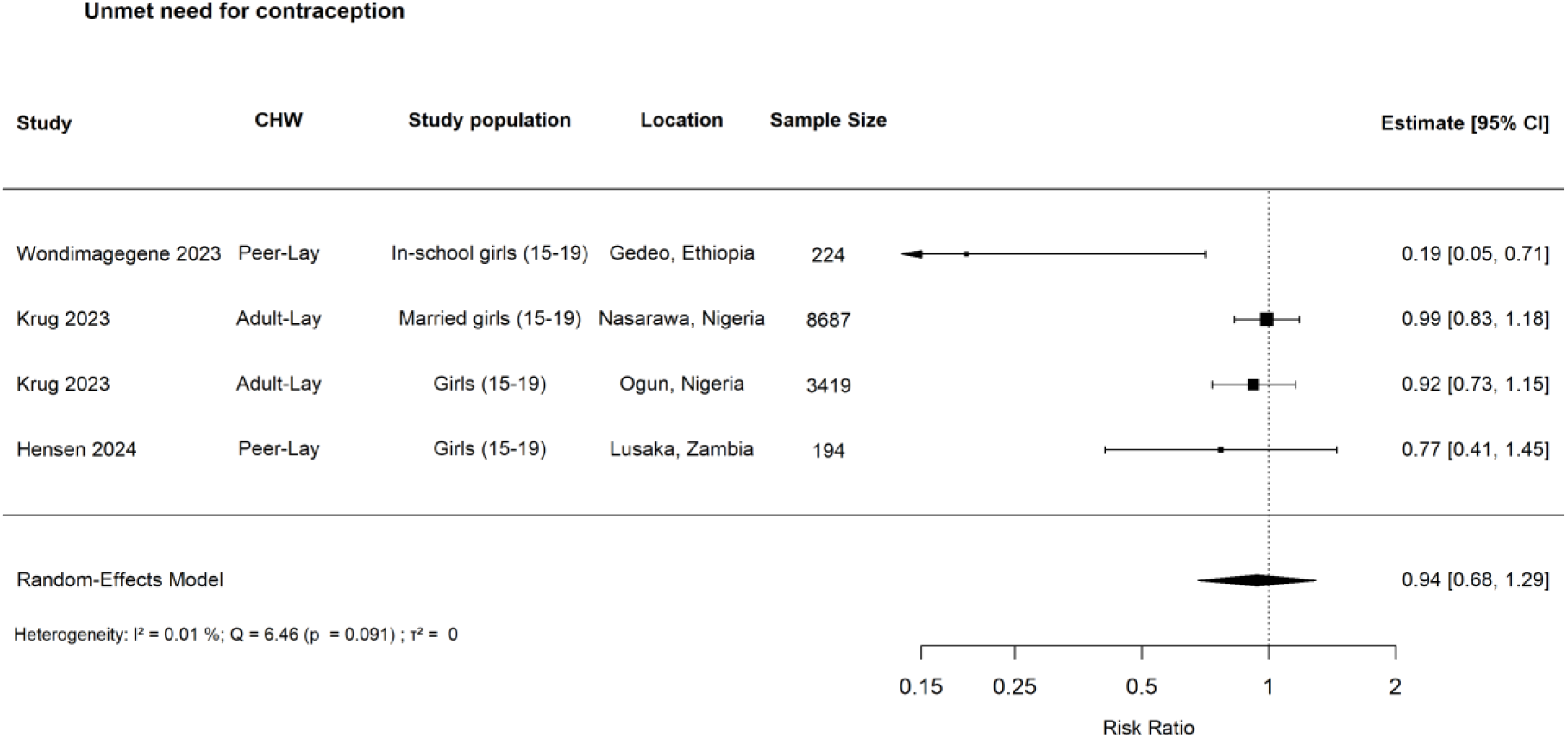
Forest plot: risk ratio for the effects of CHW-led interventions on unmet need for modern contraceptives among adolescents in SSA. **Note:** All the four reported effect sizes were included in the meta-analysis above due to standard measurement. Visual inspection of the funnel plot (see Appendix H.2) suggests possible asymmetry with an uneven distribution of studies and a lack of smaller studies on one side of the funnel. However, the small number of included studies (n=4) limits interpretability. Details about publication bias, heterogeneity, and imprecision for this outcome can be found in Appendix H.

**Figure M.3.**
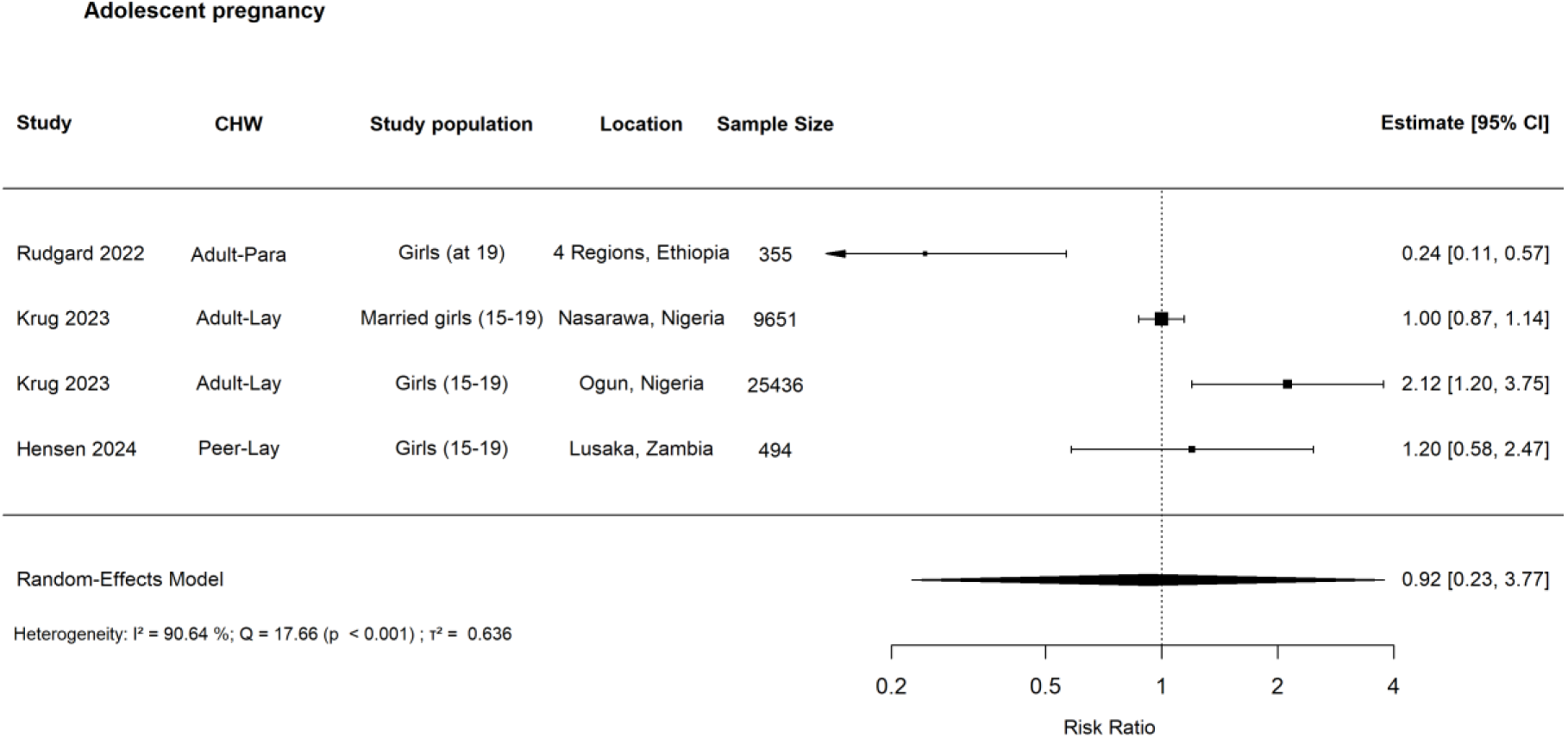
Forest plot: risk ratio for the effects of CHW-led interventions on adolescent pregnancy among adolescents in SSA. **Notes:** Four of seven reported effect sizes were included in the meta-analysis. Two of the excluded effect sizes measured age at first birth [15]. In Hensen et al. (2023), one of the two effect sizes was excluded because it focused only on sexually active girls aged 15-19, while we included the one that considered all girls in this age group [33]. Given the presence of small studies reporting both positive and negative results shown in the funnel plot (see Appendix H.2), and the limited number of effect estimates (n=4), we did not downgrade for publication bias but note a moderate level of uncertainty. Details about publication bias, heterogeneity, and imprecision for this outcome can be found in Appendix H.

**Figure M.4.**
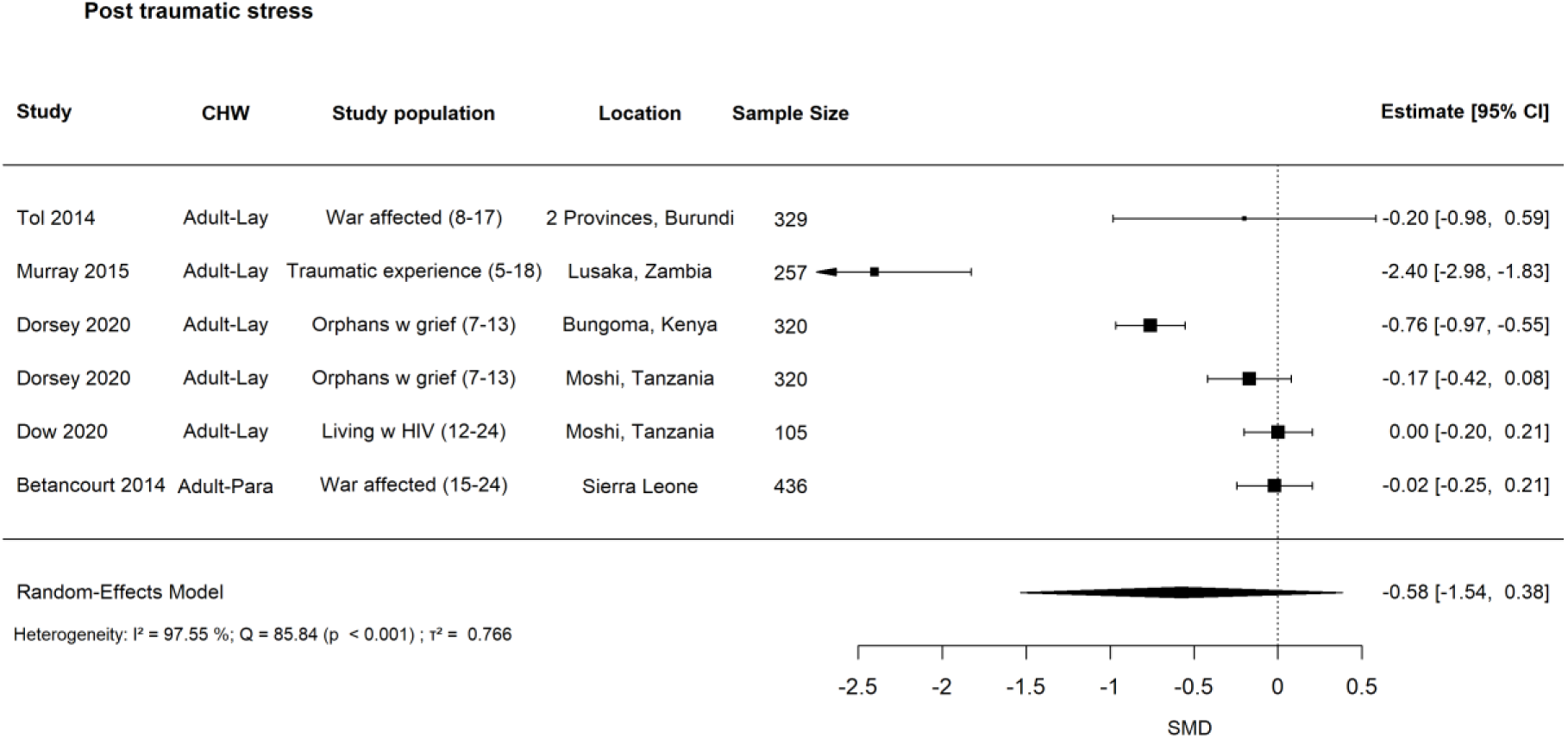
Forest plot: standardised mean difference (SMD) for the effects of CHW-led interventions on PTS among adolescents in SSA. **Note:** Six of 13 effect sizes were included in the meta-analysis above. In the main analysis here, we combined urban and rural estimates within Kenya and Tanzania respectively in Dorsey et al. (2020) [16] using fixed-effect inverse-variance weighting, and included only country-specific estimates in the meta-analysis. It is reasonable to assume that populations across country settings are independent within the study. In addition, we prioritised child self-reports over parent reports, leading to the exclusion of four effect measures from Dorsey et al. (2020) [16]. This decision reflects the view that internal states, such as stress and anxiety, are more accurately captured through self-report [16]. We also excluded one of the effect sizes reported in Betancourt et al. (2014) that measured psychological distress, while retaining the estimate measuring PTS, which is more closely aligned with our outcome of interest [35]. Visual inspection of the funnel plot (see Appendix H.2) suggests some asymmetry, with a relative paucity of small studies reporting negative or null effects. This may indicate potential publication bias or small-study effects. However, given the limited number of included studies and potential heterogeneity in study characteristics, this finding should be interpreted with caution. Details about publication bias, heterogeneity, and imprecision for this outcome can be found in Appendix H.

## Footnotes

1 Details about which specific words were used and their reasons were provided in Appendix A.3.

2 Many of the studies we reviewed evaluated lay workers, particularly peers, who delivered education, counselling and support to adolescents in primary healthcare facilities or hospitals. However, we did not classify these as community health worker interventions unless they included outreach activities, such as home visits, service provision (e.g., ART distribution), or community-based event organisation. Examples of studies that did not meet these criteria include the VUKA Family Programme [26], the Integrated Youth Centre Rewards Programme [27], in-clinic adolescent peer group support for engagement [28], the Family Clinic Day Intervention [29], and the Teen Club Intervention [30].

3 Similarly, within a given study, if some results were derived using an appropriate comparative design but others relied on uncontrolled pre-post analyses (e.g., examining changes within the intervention group only), we excluded those *outcomes*. Such results were also omitted from the vote-counting analysis, as they did not meet our comparator-related inclusion criteria.

4 While the HKSJ method performs well in the presence of observed heterogeneity, it may produce overly wide confidence intervals when the number of studies is small [56]. Therefore, we additionally reported conventional Wald-type confidence intervals as a sensitivity analysis to assess the robustness of results under alternative inferential assumptions, particularly given the limited number of studies in several meta-analyses.

5 The total number of programmes/interventions across community and school settings, as well as the total number of programmes led by peer and adult CHWs, are both 37, which exceeds the 35 included studies, because they are counted at the intervention level rather than the study level. There are two included studies that spanned across two country settings.

6 This refers to knowledge of service location in Krug et al. (2023) [63].

7 A seemingly counterintuitive finding that could be potentially explained by increased preventive knowledge, positive attitude changes and enhanced family support perceptions [90].

8 Gender-matched group interventions refer to a modality in which female CHWs engage with adolescent girls, and male CHWs engage with adolescent boys. The count of gender-matched group interventions is distinct from the count of female CHW-led interventions, as the former includes both female and male CHWs.

9 For Rudgard et al. (2022) [68] and Posso et al. (2021) [61], they studied the HEWs in Ethiopia where more than 95% of the CHW workforce are female.

10 In accordance with Cochrane guidelines [95], we present summary tables of robustness test results for each outcome without accompanying meta-analysis plots, as including multiple forest plots would substantially increase the length of the document without adding meaningful value.

11 Which create social expectations for early childbearing after marriage.

12 Notably, the only included study in our review showing reduced pregnancy pointed to indirect benefits through school retention and delayed marriage [68], although more robust research is needed.

13 Two out of four effect sizes were positive [72], and the remaining two were null [63].

14 There are several counterintuitive inconsistencies in the findings, which raised concerns about the validity of the results. For example, Austrian et al. (2021), they found improvements in contraceptive knowledge among participants who received reproductive health education only, but no positive effect in the treatment group that received both reproductive health education and sanitary pad distribution [58].

15 To accurately isolate the effect of CHWs, the control group would ideally receive the same products, services, and education materials as the intervention group, differing only in delivery by CHWs. However, few included studies met this criterion. Accordingly, our findings pertain to CHW-led interventions rather than the independent effect of CHWs and should be interpreted with this in mind.

16 Based on the intervention descriptions, CHWs appear to constitute the primary delivery component. However, this assumption should be interpreted with caution.

17 Namely, the crude proportion of participants with the outcome in question.

3 We did not rely on p-values. For details about our assessment criteria of baseline comparability, please refer to Note a under Table H.1.1.

4 Studies included in the meta-analysis are the primary basis for GRADE assessment, but other studies examining the outcome, though not included in the meta-analysis, should also be considered when evaluating indirectness. If these excluded studies differ substantially in terms of PICO from those in the meta-analysis, this may raise concerns about the applicability of the pooled effect estimate across settings.

5 Including all effect estimates, regardless of being in meta-analysis or not.

6 When counting the effect estimate in Austrian et al. (2021) [25] for IPV, we combined the two treatment arms (pads only and reproductive health education only) following the Cochrane guideline [42], and the combined effect suggests no evidence.

7 Self-efficacy in sexual relations was categorised under SRHR.

